# The relationship of early-life diseases with lifetime childlessness: Evidence from Finland and Sweden

**DOI:** 10.1101/2022.03.25.22272822

**Authors:** Aoxing Liu, Evelina T Akimova, Xuejie Ding, Sakari Jukarainen, Pekka Vartiainen, Tuomo Kiiskinen, Sara Kuitunen, Aki S Havulinna, Mika Gissler, Stefano Lombardi, Tove Fall, Melinda C Mills, Andrea Ganna

**Affiliations:** Institute for Molecular Medicine Finland, FIMM, University of Helsinki, Helsinki, Finland; Program in Medical and Population Genetics, Broad Institute of MIT and Harvard, Cambridge, MA, USA; Leverhulme Centre for Demographic Science and Nuffield College, University of Oxford, Oxford, United Kingdom; Finnish Institute for Health and Welfare, THL, Helsinki, Finland; Research Centre for Child Psychiatry and Invest Research Flagship, University of Turku, Turku, Finland; Academic Primary Health Care Centre, Stockholm, Sweden; Department of Molecular Medicine and Surgery, Karolinska Institutet, Stockholm, Sweden; Institute for Economic Research, VATT, Finland; Molecular Epidemiology, Department of Medical Sciences, and Science for Life Laboratory, Uppsala University, Uppsala, Sweden; Department of Economics, Econometrics & Finance, Faculty of Economics and Business, University of Groningen, the Netherlands; Department of Genetics, University Medical Centre Groningen, the Netherlands; Analytic and Translational Genetics Unit, ATGU, Massachusetts General Hospital, Boston, MA, USA

**Author notes:** Corresponding authors: Melinda C Mills and Andrea Ganna.

## Abstract

The percentage of women born 1965-1975 remaining childless is ∼20% in many Western European and ∼30% in some East Asian countries. Around a quarter of childless women do that voluntary, suggesting a remaining role for disease. Single diseases have been linked to childlessness, mostly in women, yet we lack a comprehensive picture of the effect of early-life diseases on lifetime childlessness. We examined all individuals born 1956-1968 (men) and 1956-1973 (women) in Finland (n=1,035,928) and Sweden (n=1,509,092) to completion of reproduction in 2018 (age 45 women; 50 men). Leveraging nationwide registers, we associated sociodemographic and reproductive information with 414 diseases across 16 categories, using a population and matched pair case-control design of siblings discordant for childlessness (71,524 full-sisters, 77,622 full-brothers). The strongest associations were mental-behavioural, particularly amongst men (schizophrenia, acute alcohol intoxication), congenital anomalies and endocrine-nutritional-metabolic disorders (diabetes), strongest amongst women. We identified novel associations for inflammatory (e.g., myocarditis) and autoimmune diseases (e.g., juvenile idiopathic arthritis). Associations were dependent on age at onset, earlier in women (21-25 years) than men (26-30 years). Disease association was mediated by singlehood, especially in men and by educational level. Evidence can be used to understand how disease contributes to involuntary childlessness.

Text box:
**Defining Childlessness**
We use the term childlessness to describe individuals that have had no live-born children by the end of their reproductive lifespan (age 45 for women; 50 for men). Childlessness is defined in the literature as being both involuntary, related to biology and fecundity (e.g., infertility, inability to find a partner) and voluntary or ‘childfree’^1^ (e.g., active choice, preference^2^). It has been estimated that 4-5% of the current 15-20% women who are childless in Europe are voluntary childless^3^. Childless individuals are subjected to discrimination and marginalization in many societies^4^, with infertile women globally experiencing multiple types of violence and coercion^5^. A parallel line of work, which is not the position of this paper or authors, is to problematize and stigmatize childless individuals as egoistic and place blame on this group for producing a so-called ‘demographic disaster’ of shrinking and ageing populations and collapse of social security systems^6^. The approach of this paper is to provide a neutral, data-driven, and factual examination of early-life diseases related to childlessness, with the aim to design a better understanding of health to prevent childlessness among those who want to have children.

## Main

In Western European countries, around 20% of women born around 1965 remained childless, with the highest levels of permanent childlessness now in East Asia, ranging from 28% (Japan) to 35% (Hong Kong) for women born in 1975^7,8^. There has also been a sharp increase in childlessness over the last decades in countries such as in Finland, increasing since the 1970s from 14% to 22% (women aged 40 years) and 22% to 32% (men)^9,10^. Demographic research has isolated core factors linked to the rise in contemporary levels of childlessness. Access to effective contraception at the end of the 1960s in many countries and more recently, emergency post-coital contraception in the late 1990s, provided couples with more abilities to control their reproduction^7^. Women’s opportunities also vastly changed in many industrialised nations since the 1980s, with gains in higher levels of education and entry into the labour market accompanied with shifts in gender norms and equity^11,12^. At the same time, couples faced work-life reconciliation constraints when planning to have children including lack of access to childcare, challenging housing conditions, economic uncertainty, inability to find a partner, and the general absence of supportive family policies^12^. Given that women entered the labour market while still taking on the bulk of household labour, this ‘incomplete gender revolution’ resulted in women often being forced to decide between a career and parenthood^13,14^. Higher levels of job strain and lack of work-life reconciliation has also been found to reduce fertility intentions^15^. Another shift has been the growth of precarious, temporary, and uncertain employment, leading many to postpone or forgo entering partnerships and parenthood^16,17^. Many of these changes resulted in a general postponement of children to later ages^12^. This in turn meant that individuals were having children at ages when they had lower fecundity (reproductive capacity), leading to infertility-related issues and having fewer children^18^. This also resulted in an increase in involuntary childlessness (see **Text Box**).

Although economic, societal, and cultural freedom has been linked to individuals who remained ‘childfree’ or voluntarily childless^19^, the individuals who report that they never intended to have children, however, has remained low and is estimated to be around 5% in Europe^3^. Rather than being planned at an early age, becoming voluntarily childfree appears to be a mix of unforeseen circumstances, often related to postponement of childbearing and adaptation to a childfree life^20,21^. This makes the decomposition of measuring voluntary versus involuntary childlessness challenging. A longitudinal study measuring fertility intentions of childless women (questioned from age 14 to their late 40s) found that they engaged in repeated postponement of childbearing and the subsequent adoption of a childless expectation at older ages or had indecision about parenthood evidenced by changing childless expectations across various ages.

The majority of childless individuals are thus involuntarily childless and wanted to have children. From the standpoint of public health, involuntary childlessness may impact other health domains, with childless women more likely to suffer from relationship dissolution, lower levels of self-esteem and isolation, and higher risks of clinical depression^12^. A recent systematic review found that infertile women globally are also more likely to experience psychological, physical, and sexual violence as well as economic coercion^5^.

While the aforementioned demographic and socioeconomic literature has shown social, economic, and structural factors as having a major effect on childlessness, the fact that a large proportion of individuals are still involuntarily childless suggests that biology and diseases likely have a considerable influence on individuals’ chances of being childless or having a particular number of children over their lifetime (**Figure S1**). First, diseases may directly influence childlessness through medical conditions that affect the fecundity, co-morbidities and mortality of the individual, as well as the risk of stillbirth^22^. Second, diseases can impact selection into a partnership, which in turn lowers the chance of being partnered and thus having children^23^. Also, some may choose to be childfree due to knowledge or concerns of intergenerational transmission of certain known genetic conditions^24^. Finally, many diseases are associated with lower socioeconomic status, unemployment, and economic uncertainty, which may amplify the psychosocial impact of diseases on childlessness^25^.

A review of the literature linking disease to childlessness can be found in **Table S1**. Previous medical studies largely only examined a limited selection of diseases directly associated with reproductive biology such as recurrent miscarriage^26^, polycystic ovary syndrome^27^, and endometriosis^28^. Another large area of research has been the study of infertility and inflammatory bowel disease (IBD). A systematic review found that up to one-third of IBD patients opted to be voluntarily childlessness, often related to lack of knowledge related to pregnancy-related IBD issues^29^, with clinical studies finding that women with moderate to severe IBD had higher incidences of adverse pregnancy outcomes^30^. Multiple sclerosis has also been associated with higher levels of childlessness, linked to fewer being in a stable relationship, fear of genetically transmitting multiple sclerosis, or discontinuation of treatments^31^. Another line of study has examined fecundity in patients with major psychiatric disorders^32^, and those with a genetic liability for schizophrenia, with the later finding no clear association with childlessness^33^.

Current research has examined singular diseases in parallel and thus lacks a systematic assessment of the role of multiple diseases on childlessness and their relative importance. Moreover, the association of diseases can be sex-specific, given that men and women differ regarding both reproductive patterns (men have a higher chance of being childless, higher parity, and a longer reproductive lifespan than women^34^) and the prevalence and severity of diseases. For men, comparatively limited information has been collected on the causes and consequences of infertility and having children^35,36^, despite infertility being a major public health concern. Finally, previous studies have had a limited time horizon to follow participants and the methodological tools to control for unmeasured familial confounding, either genetic or environmental.

We address these limitations of previous research by studying the entire reproductive and health history of a cohort of Finnish and Swedish men and women until the end of their reproductive period using high-quality nationwide population registers. The current study determines to what extent diseases are associated with childlessness, whether such associations vary between sexes, differ by the age at disease onset, or stratify by parity, and the role that partnership formation and educational level plays in mediating the results. To answer these questions, we used a disease-agnostic family-based approach which allows us to robustly evaluate the relative importance of 414 disease diagnoses in relation to childlessness while providing public-health-relevant metrics to interpret the relative importance and contextualize the results of our study.

## Results

### Study population

Our study population included 1,425,640 women and 1,119,380 men, born after 1956 and alive by age 16, did not emigrate, and largely completed their reproductive period at the end of 2018 (**Fig. 1A**). Among them, 230,198 women and 279,454 men were childless. The proportion of childless individuals – those with no live-born children – was higher in men (25.4%) than women (16.6%), higher amongst Finns (23.3%) than Swedes (18.7%), and slightly higher in the general population than in individuals who had same-sex full-siblings (0.9% and 0.5% higher in women and men, respectively). Among individual with children, a two-child parity was most prevalent in men (47.1%) and women (48.9%), amongst both Finns (44.0%) and Swedes (50.8%) (**Table S2)**. Both women and men with the lowest educational attainment were more likely to be childless (e.g., in Finland, 24.2% were childless in women and 37.4% in men) than the general population (e.g., in Finland, 18.8% were childless in women and 27.7% in men) (**Table S3-S4**). When looking across generations, the education level of parents was correlated with an individual’s childless status (**Table S3-S4**). For example, among index persons whose parents had completed the first stage of tertiary education, men were less likely to be childless (e.g., in Finland, 25.3%) compared to the general population (e.g., in Finland, 27.7%) while women behaved in the opposite way (e.g., in Finland, 21.3%, compared to 18.8% in the general population).

**Fig. 1:**
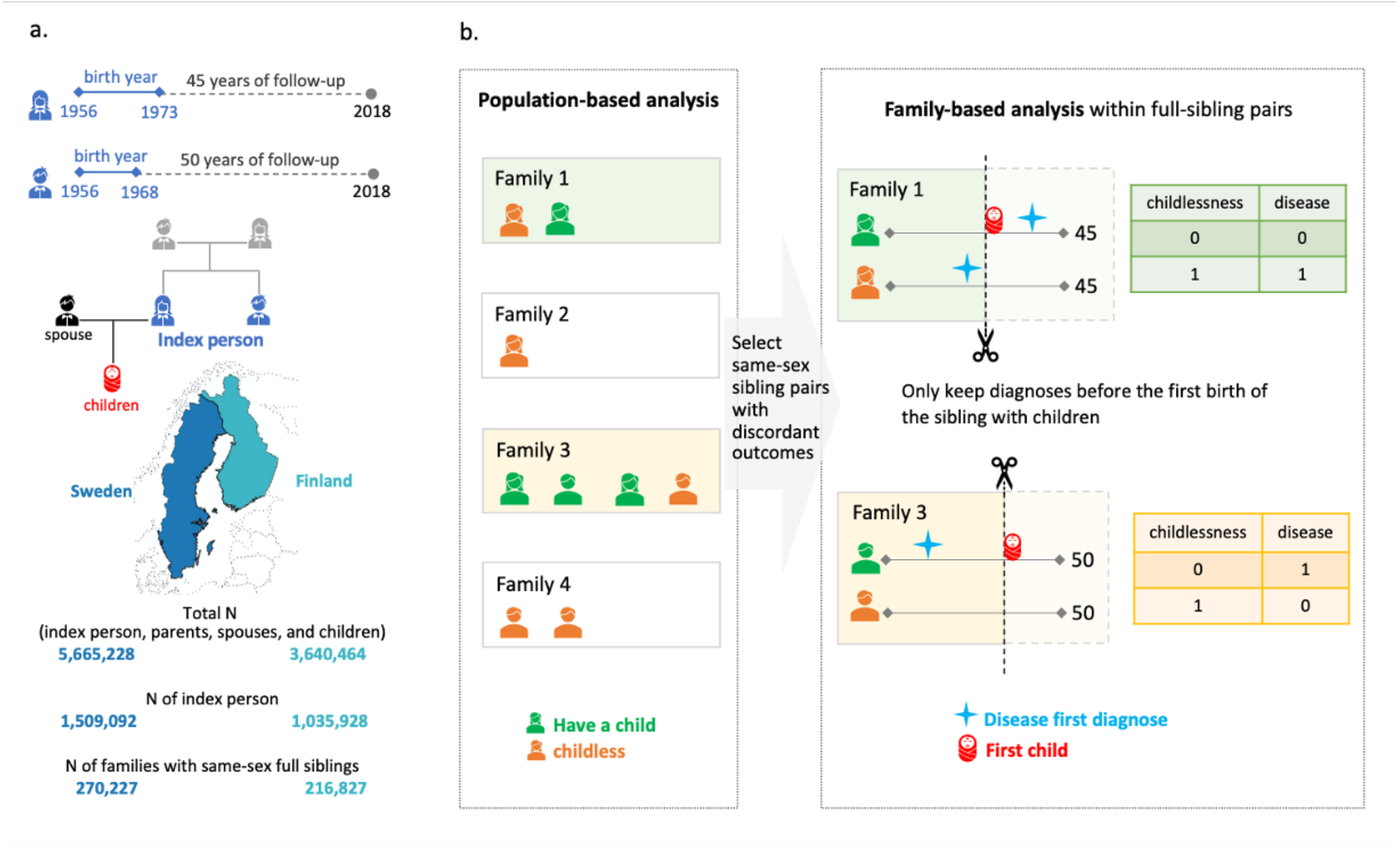
Schematic overview of the study. Panel A shows the birth cohorts, the follow-up period, and the number of Finns and Swedes included in the analyses. A total of three generations of familial relationships were considered for the index persons, with the parental information used to match sibling pairs, the children’s information to define the main outcome of childlessness, and the registered spouse’s information to determine the secondary outcome of singlehood. Panel B shows the statistical approach used in the main analyses, where only families with same-sex siblings discordant for childlessness are included. Within each family, we randomly selected one childless sibling as a case, and, as control, the sibling with children that was the closest for birth order with the case. Within a sibling pair, disease diagnoses are only considered if they occurred at least one year before the birth of the first child or within the corresponding age at which the sibling was childless, since sibling ages will differ.

For the entire study population, we have information from nationwide registers covering 414 disease diagnoses across 16 main categories (**Figure S2; Table S5**). Disease prevalence across the entire follow-up was highly comparable between Finland and Sweden (**Figure S3**) and we presented meta-analysis results between the two countries. Our main analysis focused on 71,524 full-sister and 77,622 full-brother pairs who were discordant on childlessness (**Fig. 1B**).

### Disease diagnoses associated with childlessness

Eleven rare diseases were associated with an almost complete lack of children (e.g., severe intellectual disability, childhood leukemia, and muscular dystrophy) (**Table S6**) and were therefore not included in all the remaining analyses. Out of the remaining 403 diseases (328 in women; 325 in men), 74 were significantly associated with childlessness in at least one sex (P<1.5× 10^−4^, after multiple-testing correction), with 33 disease diagnoses shared among women and men (**Fig. 2**). A full list of the results can be found in **Table S7** or in an interactive dashboard: https://dsgelrs.shinyapps.io/DiseaseSpecificLRS/. More than half of the significant associations were mental-behavioral disorders (26/53 (49%) in women and 30/54 (55.5%) in men), which was, together with congenital anomalies, the disease categories with the strongest associations with childlessness (OR=3.1 [95% CI: 2.6-3.7] in women compared to men (OR=3.2 [CI: 2.6-4.0]), averaging over all mental-behavioral disorders (**Fig. 2** and **Table S8**). There was substantial heterogeneity among different mental-behavioral disorders (P=4.5× 10^−71^ in women and 7.9× 10^−128^ in men in a heterogeneity test). For example, mild intellectual disability was the condition with the strongest association with childlessness (e.g., OR=21.7 [10.8-43.5] in men), while smaller effects were observed for mood disorders (e.g., 2.1 [1.8-2.3] in men). Severe diseases of the brain such as cerebral palsy (OR=13.4 [7.9-22.7] in women and 12.0 [6.5-22.2] in men) and malignant neoplasm of the brain (5.5 [2.8-10.8] in women and 5.0 [2.8-8.9] in men), which impair health and functioning in several ways but can also result in behavioral and personality disorders^37,38^, were also strongly associated with childlessness. For individuals with severe mental disorders or physical disabilities (e.g., intellectual disability), especially amongst women, involuntary sterilization was historically carried out in Finland (from 1935 to 1970) and Sweden (1934 to 1976)^39^.

**Fig. 2:**
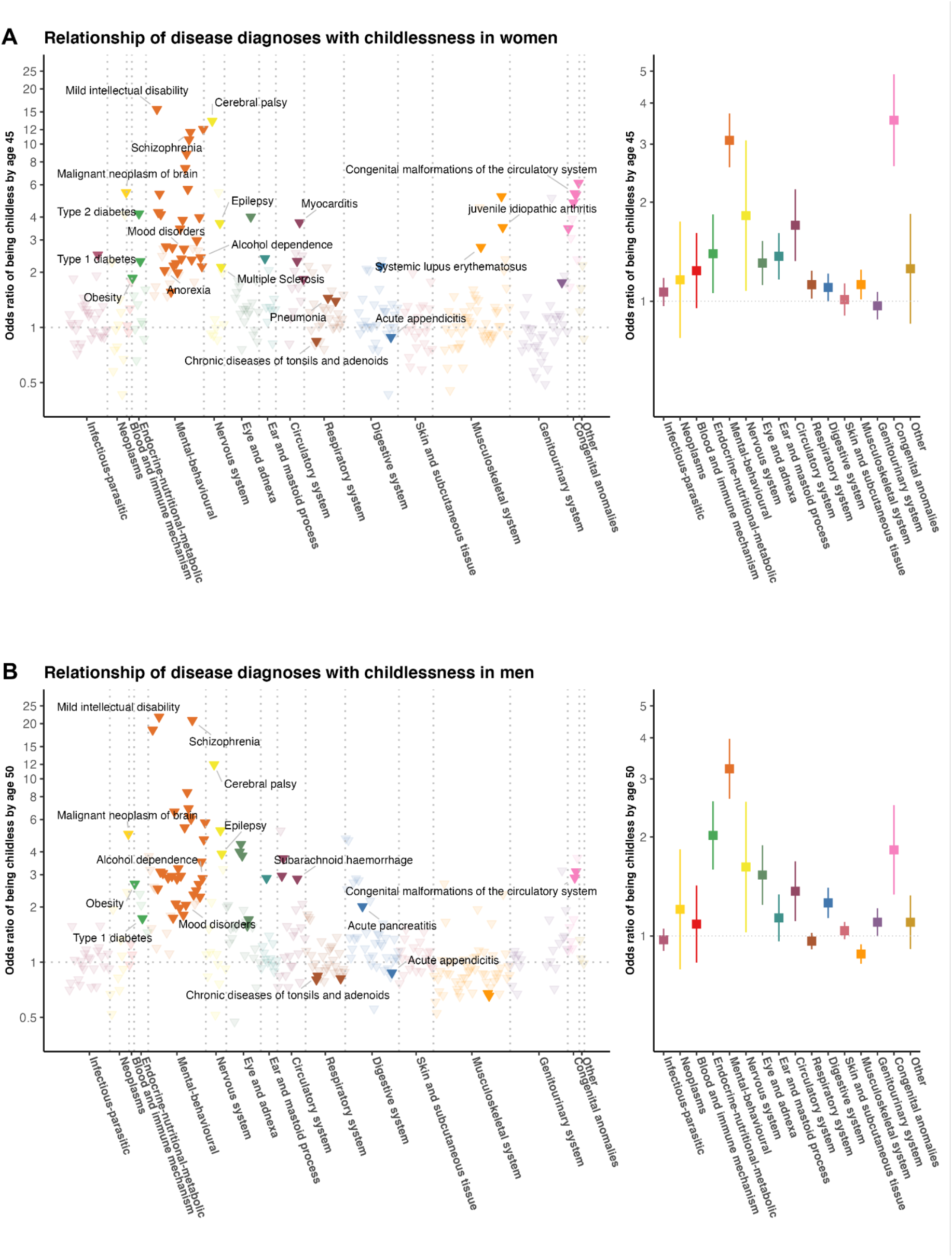
**Relationship of 403 disease diagnoses with childlessness by age 45 in women (panel A) and 50 in men (panel B) in 71,524 full-sister and 77,622 full-brother pairs who were discordant on childlessness, using a matched case-control design** Odds ratios are computed for each disease diagnosis (left figures of each panel) and averaged over disease categories (right figures of each panel). Only disease diagnoses that are significantly associated with childlessness after multiple-testing correction (P <1.5× 10^−4^) are colored. Labels are assigned only for certain disease diagnoses that are described in the text.

Another disease category strongly associated with childlessness, in both sexes, encompassed the endocrine-nutritional-metabolic disorders (OR=1.4 [1.1-1.8] in women and 2.0 [1.6-2.6] in men, averaging over all endocrine-nutritional-metabolic disorders). For example, a diagnosis of obesity (OR=1.8 [1.5-2.4] in women and OR=2.7 [2.0-3.6] in men), which is normally recorded in the secondary healthcare registers only in severe cases, and type 1 and type 2 diabetes (e.g., in women, OR=2.3 [2.0-2.6] for type 1 diabetes and 4.2 [2.2-8.1] for type 2 diabetes) were among the diseases with the strongest associations with childlessness.

We found several inflammatory diseases to be significantly associated with childlessness across multiple organ systems including respiratory, circulatory, genitourinary, digestive, nervous, and musculoskeletal systems. For example, a diagnosis of pneumonia (OR=1.4 [1.3-1.6]), myocarditis (3.7 [2.3-6.1]), chronic tubulo-interstitial nephritis (1.8 [1.3-2.3]), multiple sclerosis (2.1 [1.5-3.1]), systemic lupus erythematosus (2.7 [1.6-4.6]), and juvenile idiopathic arthritis (3.5 [2.6-4.9]) significantly increased subsequent childlessness in women.

Puzzlingly, but consistently with previous findings^40^, we observed that chronic diseases of tonsils and adenoids and acute appendicitis, were associated with reduced, rather than an increased odds of childlessness for women and men (e.g., for chronic diseases of tonsils and adenoids, OR=0.84 [0.80-0.88] in women and 0.84 [0.80-0.89] in men), with similar effect sizes in Finns and Swedes (P for heterogeneity=0.60 between two countries in both sexes).

### Sex-specific effects

The disease category encompassing congenital anomalies showed markedly different effects between sexes (OR=3.5 [95% CI: 2.6-4.9] in women and men [OR=1.8 [1.3-2.5], P for sex difference=3.7× 10^−3^). Overall, malformations of the digestive system (4.8 [2.2-10.3] in women and 1.7 [1.0-3.0] in men, P=0.03) and musculoskeletal system (3.5 [2.0-6.1] in women and 1.2 [0.7-1.9] in men, P=3.7× 10^−3^) were more strongly associated with childlessness in women than men.

Significant sex-dependent effects were also observed for several mental-behavioral and endocrine-nutritional-metabolic disorders (**Fig. 3A**). For example, diagnoses of schizophrenia (OR=11.6 [9.1-14.9] in women and 20.8 [16.1-27.0] in men, P for sex difference=1.5× 10^−3^) and acute alcohol intoxication (2.2 [1.6-2.9] in women and 4.7 [3.8-5.7] in men, P=4.0× 10^−5^) had stronger associations with childlessness in men compared to women. Diagnosis of type 1 diabetes showed stronger association in women than in men (2.3 [2.0-2.6] in women and 1.7 [1.5-2.0] in men, P=3.9× 10^−3^). The stronger association in women might be due to the fact that, in the observational period, women with type 1 diabetes were recommended against pregnancies if under poor glycemic control^41^.

**Fig. 3:**
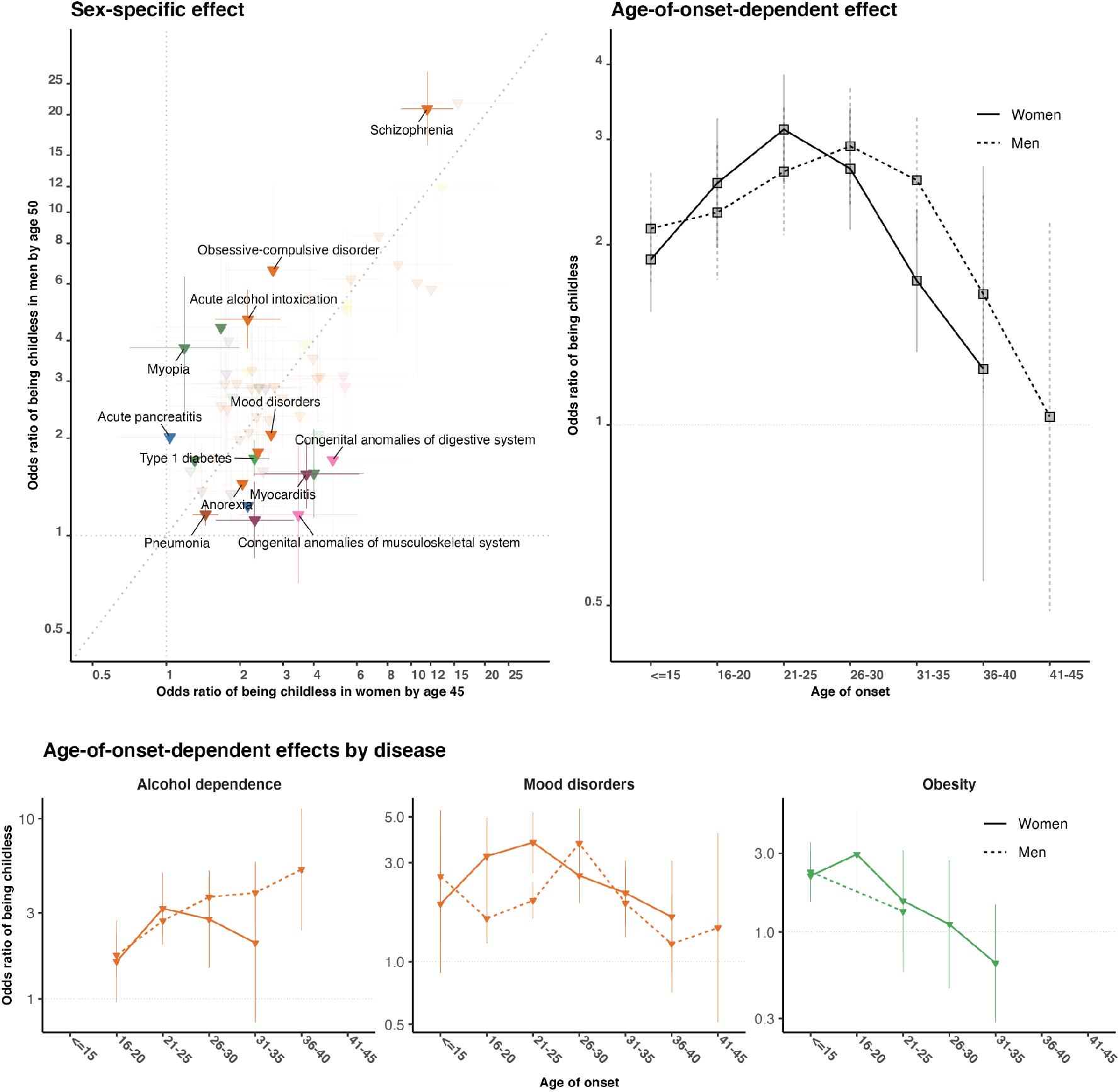
**Sex-specific (panel A) and age-of-onset-dependent effects (panel B and C) for the association between disease diagnoses and childlessness in 71,524 full-sister and 77,622 full-brother pairs who were discordant on childlessness** In panel A, we compare the odds ratio for 60 disease diagnoses that significantly increased the odds of childlessness either in men or women. Only disease diagnoses that are significantly different between sexes at a nominal P value are colored. In Panel B we report, for each age category, the average effect associated with childlessness across 30 disease diagnoses in women and 31 in men as obtained from an age-of-onset-stratified analysis. In Panel C, we report age-of-onset-stratified odds ratios associated with childlessness for three major diseases/disorders for which we observe a significant trend. Estimates for each age category are computed only if the number of individuals with this disease within the age group is more than five.

### Age-of-onset of disease effects

We hypothesized that the age when the disease was first diagnosed, a proxy for the age of onset, would influence the chances of being childless by either capturing disease severity or directly impacting factors underlying individuals’ reproductive trajectory. We first evaluated if there was a general trend across disease diagnoses (30 in women and 31 in men) that were associated with childlessness and for which we had enough individuals to estimate age-of-onset-stratified effects (**Fig. 3B** and **Table S9-S10**). We observed a non-linear effect, with the strongest association with childlessness occurring when the disease was first diagnosed between 21-25 years old in women (OR=3.1 [2.5-3.8]) and later, between 26-30 years old, in men (3.1 [2.4-3.9]). Smaller effects were observed at younger ages (e.g., in women, OR=1.9 [1.6-2.3] for onset before 16 years old, P=7.1× 10^−4^) and, especially, older ages of onset (e.g., in women, 1.2 [0.5-2.8] for onset between 36-40 years old, P=0.04). Despite this broader trend, there was substantial heterogeneity between disease diagnoses (**Fig. 3C**). For example, among women diagnosed with obesity, the group that received their first diagnoses between 16-20 years old had higher levels of childlessness than those diagnosed at later ages (OR=3.0 [1.6-5.6] for diagnosed between 16-20 years old and 1.1 [0.5-2.7] between 26-30 years old, P differences=0.08).

### Stratification by parity for individuals with children

In the main analyses we compared childless individuals to their siblings with children, regardless of parity. We reasoned that childless siblings might be more phenotypically similar to their siblings with fewer children (lower parity) compared to siblings who had multiple children (higher parity). When comparing childless individuals to their siblings with just one child, 14 disease diagnoses in women and six in men had significantly reduced ORs (**Figure S4A-B**) compared to the main analysis. For example, in women, the OR of schizophrenia on childlessness dropped from 11.6 [9.1-14.9] to 5.1 [3.7-7.0] (P difference=6.9× 10^−5^) when we compared childless individuals with their siblings with just one child. For individuals with higher parities, such as having exactly two children (**Figure SC-D**) or those with at least three or more children (**Figure S4E-F**), limited differences in ORs were observed compared to the main analyses. Jointly, these results indicated that childless individuals were more similar to their siblings with one child with regard to risk for childlessness-associated early-life diseases compared to siblings with higher parities.

### Number of unborn children and completed fertility rate reduction related to disease diagnoses

Having identified a set of disease diagnoses that conferred a larger relative odds of childlessness, we estimated the number of ‘missing children’ not born in the observational period due to the disease diagnoses by combining relative risk, disease prevalence, and parity. We estimated that 2,728 [2,586-2,871] children per 100,000 men and 1,489 [1,376-1,605] per 100,000 women were missing or not born due to their parents having at least one mental-behavioral disorder (**Table S13-S14**). Accordingly, this disease category was associated with 68% of total unborn children related to disease diagnoses among males and 59% among female participants. Disease-specific analyses revealed that schizophrenia was linked to the largest number of children that were not born between both sexes (893 [848-930] children per 100,000 men, 612 [549-669] per 100,000 women) followed by anxiety disorders among men (569 [475-665]) and mood disorders among women (389 [313-470]). We note that the estimates of the number of children not born or ‘missing’ due to disease diagnoses could be slightly biased because of the use of only individuals with same-sex full siblings in our main analyses.

The completed total fertility rate (TFR), partitioned as the sum of disease and non-disease age-specific fertility rates for each sex, was markedly lower among individuals who carry any of the disease diagnoses significantly associated with childlessness (TFR=1.5 for women and 1.1 for men) compared to the unaffected individuals (2.0 for women and 1.8 for men), resulting in an overall TFR reduction of 38% in women and 21% in men (**Table S15-S16)**. Overall, affected women and men accounted for 5.4% and 4.6% of overall observed population-level TFR, respectively.

### Mediation effect by singlehood

There were 83.0% women and 77.0% men having registered partners before age 45 and 50, respectively. Restricting to childless individuals, the proportion dropped to 36.3% in women and 29.4% in men. We therefore examined to what degree the associations between disease diagnoses and childlessness were mediated by singlehood. First, we estimated the effect of disease diagnoses on the chance of being without a partner by age 45 in women and 50 in men **(Figure S5-S6**). We then compared log ORs on singlehood with log ORs on childlessness and observed a high correlation between these two estimates in both women (R^2^=0.71, P=1.0× 10^−15^) and men (R^2^=0.85, P=3.5× 10^−23^) (**Figure S7**). A causal mediation analysis performed for each disease also indicated the mediation role of partnership formation, with 29.3% (median) of the disease effects on childlessness in women and 37.9% in men mediated by singlehood (**Table S17**). Different patterns were observed across diseases. For example, singlehood was a significant mediator for women diagnosed with schizophrenia (OR=2.1 [2.0-2.2] for indirect and 6.8 [5.0-9.2] for direct effect) but not for women with hypertension (1.2 [1.1-1.4] for indirect and 2.4 [1.6-3.5] for direct effect).

Next, we investigated if some of the associations between disease diagnoses and childlessness remained significant among partnered individuals. We considered 133 disease diagnoses in women and 123 in men, which had more than 30 affected individuals with registered partners in the sibling-based analysis. Some diseases that were strongly associated with singlehood (e.g., mild intellectual disability) could not be included in the analyses because they were too rare among partnered individuals. Nonetheless, we observed six diseases in women and eleven in men that remained associated with childlessness among partnered individuals (**Figure S8**). Compared to the estimates obtained from both partnered and unpartnered individuals, the effects on childlessness were largely reduced for most disease diagnoses **(Figure S9)**, such as epilepsy (e.g., from OR=3.7 [3.2-4.3] for all women to 1.8 [1.4-2.4] for partnered women) and acute alcohol intoxication in men (from 4.7 [3.8-5.7] for all men to 2.4 [1.5-3.8] for partnered men). Several endocrine-nutritional-metabolic diseases retained a strong association with childlessness among partnered individuals, such as obesity in men (OR=2.7 [2.0-3.6] for all men and 3.3 [1.9-5.7] for partnered men) and type 1 diabetes in women (2.3 [2.0-2.6] for all women and 2.5 [2.0-3.2] for partnered women).

### Population-based and sensitivity analyses

We performed several sensitivity analyses to determine the robustness of our estimation. First, our main results were based on within-sibling analysis, which can account for unmeasured familial factors but also made several assumptions, including siblings being generalizable to the population. Thus, we also obtained population-based estimates from a matched case-control design controlling for several potential confounders (**Figure S10**). Overall, the population-based design resulted in larger sample sizes, similar ORs, and smaller CIs compared to the sibling-based design (**Figure S11**). However, stronger associations were observed from the population-based design in men for psychoactive substance abuse (e.g., for alcohol dependence, OR=2.9 [2.5-3.2] from sibling-based design and 3.8 [3.5-4.0] from population-based design, P differences=1.4× 10^−4^) and mood disorders (2.1 [1.8-2.3] from sibling-based design and 2.7 [2.5-2.9] from population-based design, P=2.0× 10^−4^).

Second, in the main analysis, we included individuals who died before the end of follow-up (affecting 7.4% full-sisters and 15.0% full-brothers) thus including reproductive-age mortality as one of the possible mechanisms explaining childlessness. To understand the overall effect of reproductive-age mortality, we also conducted an analysis only considering individuals alive by the end of follow-up (**Figure S12**). Overall, the results were consistent (**Figure S13**), but we observed two disease diagnoses in men for which the main effects on childlessness were partially explained by reproductive age mortality: acute alcohol intoxication (OR=4.7 [3.8-5.8] for all men and 3.9 [2.2-3.8] for men alive by age 50, P differences=0.006) and subarachnoid hemorrhage (2.9 [1.8-4.5] for all men and 0.9 [0.4-1.8] for men alive by age 50, P=0.005). Third, in the main analysis, we opted for conditional logistic regression because the length of follow-up would be identical to the sibling pairs if ignoring reproductive-age mortality (**Fig. 1** and **2**). When using a Cox proportional hazards model that captured both time-varying and death (**Figure S14**), we observed similar results with reductions in some disease diagnoses **(Figure S15**).

Fourth, research has found that individuals who remain childless in Finland, particularly men, have lower levels of education, suggesting it may be important to adjust for educational level^1,42^. In the sibling-based analysis, we adjusted for differences in individuals’ education level between siblings but did not find substantial differences in our results compared to the unadjusted main analysis (**Figure S16**). In the population-based analysis, we adjusted for the individuals’ highest education level in the population-based analysis and observed significant changes in ORs for many diseases, in both women and men (**Figure S17**). For example, in men, after adjusting for the highest education level, many disease diagnoses had significantly reduced ORs (16 out of 45 (35.5%) disease diagnoses with increased risk of childlessness), while obesity (OR increased from 3.2 [2.8-3.7] to 4.4 [3.7-5.2], P differences=3.5× 10^−3^) and acute pancreatitis (OR increased from 1.9 [1.6-2.3] to 2.5 [2.1-3.0], P differences=0.04) were the only disease diagnoses exhibiting stronger associations.

## Discussion

The rich longitudinal Finnish and Swedish nationwide population registers provided a unique resource to comprehensively assess the role and relative importance of multiple diseases on childlessness. In addition to the results summarized here, all findings can be explored on the interactive online dashboard available: https://dsgelrs.shinyapps.io/DiseaseSpecificLRS/.

Our study uses a hypothesis-free approach to estimate the associations between early-life diseases and childlessness across the entire reproductive lifespan. Importantly, we identified several novel disease-childlessness associations such as autoimmune (e.g., juvenile idiopathic arthritis, multiple sclerosis, systemic lupus erythematosus) and inflammatory diseases (e.g., myocarditis). For certain diseases, treatments such as methotrexate (immunosuppressant, widely used for rheumatoid arthritis and multiple sclerosis, and occasionally used for systemic lupus erythematosus) were reported to have side effects on fertility in women^43^.

Amongst all disease categories, mental-behavioral disorders had the strongest associations with childlessness and were associated with 68% of the total ‘missing’ or unborn children among males and 59% among females attributable to these diseases. Previous literature has given much attention to the role that diseases play in childlessness among women, while men have been understudied^44^. We observed substantial sex differences for disease-childlessness associations. For example, mental-behavioral disorders such as schizophrenia and acute alcohol intoxication had stronger associations with childlessness in men than women, while diabetes-related diseases and congenital anomalies had stronger associations amongst women. These sex-specific effects may potentially be explained by differences in disease severity, bias in diagnostic practices, partner choices, and direct biological effects on fertility. Sex differences were also observed in the impact that age-of-onset of disease has on childlessness. Overall, diseases diagnosed between 21-25 years old in women and, later, between 26-30 years old in men, tend to have the strongest association with childlessness. This is also consistent with the sex difference in age at union formation, which occurs later in men^45^.

Our analyses highlight the important effects that many diseases have on partnership formation. Childless individuals are twice as likely to be single, in line with previous research showing the importance of selection into partnership on fertility^23^. Singlehood represents a major mediator for the odds of being childless, especially in men, and we estimated that 29.3% (median) of the disease effect on childlessness in women and 37.9% in men was mediated by partnership formation. The effects of disease diagnoses on partnership formation are mediated by complex socio-cultural factors, social norms, and sex-specific behavioral preferences. For example, mental-behavioral disorders such as schizophrenia and acute alcohol intoxication were strongly associated with singlehood in men, but the effects were substantially smaller in women likely reflecting how different social and individual preferences for behavioral phenotypes correlated to mental-behavioral disorders impact partner choice in the two sexes. Nonetheless, we also identify several diseases that impact childlessness among partnered individuals. Some of these diseases are more likely to exert a direct biological effect on fertility or pregnancy complications as in the case of diabetes and obesity, whereas others such as some mental-health disorders, might impact family stability or delay the age at family formation.

Our study has several strengths. First, for the majority of the study participants, we have virtually complete coverage of health and reproductive information until the end of their reproductive period. This allows us to estimate public-health-relevant metrics such as the total number of unborn or ‘missing’ children related to diseases, which cannot be estimated with incomplete follow-up.

Second, we considered more than 400 diseases across both men and women, allowing us to compare relative effects on childlessness both between diseases and between sexes. The use of nationwide data from two countries provided a large sample of 2.5 million, and most importantly, allowed us to assess how robust our findings were in different healthcare systems and diagnostic practices. Finland has had a marked increase in childlessness since the 1970s of up to 22% for women and 32% for men^9,10^, with our analyses revealing potential underlying mechanisms such as the wider detrimental effects of alcohol dependence in men and endocrine-nutritional-metabolic disorders.

Third, by leveraging the information about family formation and singlehood, we could explore to what extent this information, which has been described as an important mediator of childlessness^46^, mediated the association between disease and childlessness. Fourth, we used a matched-sibling design to limit confounding from a shared familial environment. In our study, individuals with siblings had only a slightly lower proportion of childlessness compared to the general population. Sibling designs have been extensively used in analyses of Nordic registers and have several advantages compared to population-based estimates^47,48^.

Our study also reveals the legacy of a troubled and unjust past of reproductive rights, often stemming from eugenic thinking and discrimination related disability and gender^38,49,50^. We found that intellectual disability was the condition with the strongest association in men, but also high levels of childlessness associated with cerebral palsy and behavioural and personality disorders. Until 1970 in Finland and up to 1976 in Sweden, individuals categorized as having severe mental disorders or physical disabilities – particularly women – were subjected to involuntary sterilization^38^. We also acknowledge that women with existing diseases, such as women with heart disease, can experience serious complications from pregnancy, thereby noting that strategies for high-risk individuals need further development^51^.

Our study also has several limitations. First, disease diagnoses are largely obtained from registers covering secondary and tertiary healthcare in hospitals that capture more severe disease cases (e.g., mental health and behavioral conditions), and thus, lack of the onset and occurrence of major diseases included in primary healthcare data (e.g., low-grade metabolic conditions). Moreover, disease diagnoses have changed over time; we have defined disease outcomes by harmonizing three versions of International Classification of Diseases (ICD) codes that might capture diseases with different accuracy. Our findings are based on individuals born between 1956 and 1973 and results may thus not be entirely generalizable to more recent cohorts, because reproductive and partnering practices have changed and because better treatments might alleviate the effect of some diseases on childlessness.

Second, as with previous studies to date, we were unable to partition the impacts that diseases have into voluntary and involuntary childlessness due to a lack of data on reproductive preferences and intentions and difficulties in differentiating the two^22,52,53^. Given that estimates of the voluntary childfree group are around a quarter of total childless, we note that public health interventions should take this group of voluntary childfree individuals into account. Among factors that impact involuntary childlessness, the effect of long-term institutionalization and disease-related treatments are two aspects that future registry-based studies might be able to explore. For example, medications for epilepsy are known to be associated with pregnancy complications^54^.

Third, assortative mating may lead to an overestimation of the disease impact of childlessness among partnered couples.

Fourth, despite our attempt to use a study design that minimizes confounding and reverse causality, these biases might challenge the interpretation of the results. For example, although full siblings share roughly half of their genetic materials and similar family of origin characteristics and only siblings with the closest birth order were considered, siblings can still experience different childhood conditions and even be influenced by each other’s conditions^55^. Also, the use of only individuals with same-sex full siblings may introduce selection bias, although for most disease diagnoses their effects obtained from sibling-based analyses were not significantly different from those obtained from population-based analyses.

Finally, there is the question of the generalisability of our results to other nations. The Nordic countries have been the forerunners of demographic change in the realm of partnerships and fertility^56,57^, making these results relevant for other industrialised nations. There are also important socioeconomic fertility gradients in Sweden and Finland that mirror changes in many industrialised nations. In the Nordic countries, there has been a positive association between men’s higher education and fertility^58,59^. For Nordic women, there was initially the reverse gradient (i.e., higher levels of childlessness amongst those with higher education), but this has shifted over time with women increasingly similar to men^58^. This ‘new Nordic’ fertility regime links higher socioeconomic status to higher fertility^60^ but also multi-partner fertility^61^, a trend that has emerged in other nations as well.

In conclusion, we comprehensively described the effects different diseases, particularly those with onset prior to the peak reproductive age have on the chance of being childless over a lifetime. This evidence can be used as a basis for future studies focusing on prioritizing health interventions to counter involuntary childlessness.

## Data Availability

All results (aggregated data) can be explored on the interactive online dashboard available at https://dsgelrs.shinyapps.io/DiseaseSpecificLRS/. Due to data protection regulations, we are not allowed to share individual-level data directly. However, all Finnish and Swedish register data used in this study can be applied from national data agencies listed in appendix p 2. 

## Online Methods

### Study population

We considered all individuals born in Finland and Sweden between 1956 and 1968 (men) or between 1956 and 1973 (women) as index persons, for whom the vast majority completed their reproductive lifespan (45 years old for women, 50 for men) by 31 December 2018 (**Fig. 1A**). Individuals who emigrated during the study period were excluded to avoid incomplete follow-up for disease diagnoses and reproductive information. We further excluded individuals who died before the age of 16 to eliminate the impact of diseases on pre-reproductive survival. In total, we considered 1,425,640 index women (572,518 Finns and 853,122 Swedes) and 1,119,380 men (463,410 Finns and 655,970 Swedes). For these index individuals, we also obtained information for parents, spouses, siblings, and children for a total of 9,305,692 individuals (3,640,464 Finns and 5,665,228 Swedes).

### Childlessness (main outcome)

The primary reproductive outcome of this study is childlessness, defined as having no live-born children by the age 45 for women and 50 for men. For every index person, we extracted demographic information of all biological children born before 31 December 2018 in Finland from the Population Information System and in Sweden from the Multi-Generation Register. Medical birth information including stillbirth and the use of assisted reproductive techniques was obtained from the Medical Birth Register available since 1987 in Finland and 1973 in Sweden. We excluded children conceived by assisted reproductive techniques (0.3% Finland, 0.8% Sweden) to control for potential confounding from social inequalities in medical help-seeking for infertility, especially during the observational period.

### Marriage and partnership (secondary outcome)

For index individuals in Finland, we obtained their longitudinal marriage and partnership information between 11 April 1971 and 31 December 2018 from the Population Information System. In Sweden, we collected information of married couples and cohabiting unions with biological children between 1 January 1977 and 31 December 2017 from Statistics Sweden. We defined individuals as partnerless if they did not have any abovementioned marriage or partnership registered by age 45 (women) or 50 (men).

### Disease diagnosis (exposure)

A wide variety of disease endpoints were defined by clinical expert groups^62^, through International Classification of Diseases (ICD) codes of version 8 (1969-1986), 9 (1987-1995 in Finland, 1987-1996 in Sweden), and 10 (1996-2018 in Finland, 1997-2018 in Sweden). We collected main diagnoses (ICD codes) and admission dates for secondary healthcare inpatient hospital treatments (since 1969, but in Sweden, nation-wide coverage began in 1973 for psychiatric diagnoses and 1987 for others^63^) and specialist outpatient visits in Finland from the Care Register for Health Care (since 1998) and in Sweden from the National Patient Register (since 2001). In Finland, we obtained cancer diagnoses (International Classification of Diseases for Oncology, 3rd Edition) and corresponding dates of diagnoses from the Finnish Cancer Registry, available since 1953. Additionally, for all individuals who died before the end of follow-up, we collected the date of death and basic, immediate, and contributing causes of death from Statistics Finland and Statistics Sweden to capture additional disease diagnoses that were missing from hospital inpatient and outpatient registers. The age of onset was defined as the first record of a disease diagnosis.

In total, we considered 16 disease categories, with infectious-parasitic diseases and congenital anomalies defined only in Finland and all remaining categories defined in both countries. To remove highly correlated disease diagnoses, we estimated tetrachoric correlations between every two disease endpoints using individual-level data and only kept the one with the highest prevalence if a group of disease diagnoses had tetrachoric correlations higher than 0.7. After removing highly correlated disease diagnoses, we considered 414 diseases for which we had more than 30 affected individuals in the sibling-based analysis for each sex, in Finland or Sweden.

### Sibling-based analysis (main analysis)

There were 274,205 pairs of full-sisters (118,978 in Finland and 155,227 in Sweden) and 212,849 full-brothers (97,849 in Finland and 115,000 in Sweden) among index persons (**Fig. 1A**), which allowed us to control for potential confounding from genetic and environmental factors by performing a matched pair case-control study design (**Fig. 1B**). First, we kept only families having at least two same-sex full-siblings discordant for childlessness (at least one sibling with children and one childless). Then, within each family, we randomly selected one childless sibling as a case, and, as control, the sibling with children that was the closest for birth order with the case. Childless siblings were of similar age as siblings with children (absolute mean age difference=0.3 (SD=4.0) and 0.5 (4.4) for full-sisters and full-brothers, respectively). Finally, within each sibling pair, for both the case and the control, we only considered disease diagnoses that had their onset at least one year before the age at first birth for the sibling with children, to eliminate the impact of diseases triggered by pregnancy. With the matched design, we excluded diseases diagnosed after the reproductive lifespan in order to avoid reverse causation. We used a conditional logistic regression model to investigate the association between a disease and childlessness. Full siblings were likely to share environmental factors, especially during the pre-reproductive period. Therefore, we only adjusted for birth year effects. A similar model was used to calculate the association between diseases and singlehood. We applied Bonferroni correction (P=0.05/328=1.5× 10^−4^ for women, 0.05/325=1.5× 10^−4^ for men, where 328 and 325 are the numbers of unique diseases considered in women and men, respectively) to control for the familywise error rate.

Additionally, for disease diagnoses that were significantly associated with childlessness, we assessed whether there were any age-of-onset-dependent effects by considering the age of onset (8 groups <=15, 16-20, 21-25, 26-30, 31-35, 36-40, 41-45, or unaffected for both sexes and an additional group (46-50) for men) as fixed effects. For individuals with children, we further assessed whether the effects of disease diagnoses were consistent across parities by comparing childless individuals to their siblings with one child, two children, or more than two children.

### Population-based analysis (secondary analysis)

To estimate the population-level effect of diseases on childlessness we performed a nested incident-matched case-control design which matched each childless case to one control based on sex, birth year, municipality of birth (545 from Finland and 1,091 from Sweden), and the highest parental education level (International Standard Classification of Education 1997) (**Supplementary Materials p 52**). Parental education level was used instead of the index person education level to avoid confounding between an individual’s educational attainment and medical conditions^32^. We used conditional logistic regression analysis with no additional covariates in the model. In total, we used 226,860 matched pairs of women (107,375 in Finland and 119,485 in Sweden) and 274,941 of men (127,689 in Finland and 147,252 in Sweden) for the population-based analysis.

### Number of unborn children and completed fertility rate reduction related to disease diagnoses

To quantify the total fertility deficit related to diseases, for the studied cohort, we calculated the total number of unborn children per 100,000 people and partitioned the total fertility rate (TFR) as the sum of disease and non-disease age-specific fertility rates for each sex (**Supplementary Materials p 46**).

### Mediation effect by singlehood

To assess the role that partnership formation plays in mediating the observed associations, we performed a causal mediation analysis^64^ to partition the association between disease diagnosis and childlessness into indirect (mediated by singlehood) and direct components (**Supplementary Materials p 46**).

### Sensitivity analysis

First, to eliminate the impact of diseases on reproductive age mortality, we used the sibling design framework described above but restricted the analysis to 66,255 full-sister pairs (33,556 in Finland and 32,699 in Sweden) alive by age 45 and 66,009 full-brother pairs (32,996 in Finland and 33,013 in Sweden) alive by age 50. Second, we used the sibling design framework described above but instead of conditional logistic regression we performed a stratified Cox proportional hazards regression model considering chronological age as the time scale and disease status as a time (age) varying exposure (unaffected until disease onset and affected afterward). Follow-up started at age 16, the estimated start of the reproductive lifespan, and was censored at death or one year before the age at first birth of the control, whichever occurs first. Third, to quantify to what extent the identified associations between diseases and childlessness were mediated by social characteristics such as educational attainment, we further adjusted for individual’s highest education level for both sibling- and population-based analyses.

## Acknowledgments

This work was supported by the European Research Council (835079, 945733), The Leverhulme Trust, Leverhulme Centre for Demographic Science and ESRC/UKRI (ES/W002116/1). The computations for the Swedish data were enabled by resources in project sens2019018 provided by the Swedish National Infrastructure for Computing (SNIC) at UPPMAX, partially funded by the Swedish Research Council through grant agreement no. 2018-05973. We thank Therese Andersson (Karolinska Institutet), Amir Sariaslan (University of Helsinki), and Guosheng Su (Aarhus University) for valuable advice on statistical modeling, Gianmarco Mignogna (Massachusetts General Hospital and University of Helsinki) and David Brazel (University of Oxford) for useful discussions during project planning, and Vincent Llorens (University of Helsinki) for suggestions on disease coding and applying Cox proportional hazards regression.

## Author contributions

AL, MCM, and AG designed the study and wrote the manuscript. AL conducted the statistical analysis and generated the figures and tables. ETA calculated the number of unborn children related to diseases and total fertility rates and wrote the relevant paragraph. XD helped in performing sensitivity analysis and drafted Figure S1. SJ, PV, and TK assisted in interpreting the results from a clinical standpoint. TK and ASH defined disease endpoints.

SK, AG, and AL developed the R shiny application. TF and MG coordinated data application and extraction for Swedish and Finnish nationwide registers. SL contributed to the editing of socio-economic data and statistical modeling. All authors discussed the results, revised the manuscript, and had final responsibility for the decision to submit for publication.

## Data availability

All results (aggregated data) can be explored on the interactive online dashboard available at https://dsgelrs.shinyapps.io/DiseaseSpecificLRS/. Due to data protection regulations, we are not allowed to share individual-level data directly. However, all Finnish and Swedish register data used in this study can be applied from national data agencies listed in **Supplementary Materials p 3**.

## References

1. Kreyenfeld, M. and Konietzka, D., 2017. Analyzing childlessness (pp. 3–15). Springer International Publishing.

2. Tanturri, M.L. and Mencarini, L., 2008. Childless or childfree? Paths to voluntary childlessness in Italy. Population and development review, 34(1), pp.51–77.

3. Miettinen, A. and Szalma, I., 2014. Childlessness intentions and ideals in Europe. Finnish Yearbook of Population Research, 49, pp.31–55.

4. Dykstra, P.A. and Hagestad, G.O., 2007. Roads less taken: Developing a nuanced view of older adults without children. Journal of family issues, 28(10), pp.1275–1310.

5. Wang, Y., Fu, Y., Ghazi, P., Gao, Q., Tian, T., Kong, F., Zhan, S., Liu, C., Bloom, D.E. and Qiao, J., 2022. Prevalence of intimate partner violence against infertile women in lowincome and middle-income countries: a systematic review and meta-analysis. The Lancet Global Health, 10(6), pp.e820–e830.

6. Last, J.V., 2013. What to expect when no one’s expecting: America’s coming demographic disaster. Encounter Books.

7. Sobotka T. Childlessness in Europe: Reconstructing long-term trends among women born in 1900–1972. In: Kreyenfeld M, Konietzka D, eds. Childlessness in Europe: Contexts, causes, and consequences. Cham: Springer, 2017: 17–53.

8. Sobotka, T., 2021. World’s highest childlessness levels in East Asia. Population Societies, 595(11), pp.1–4.

9. Rotkirch A, Miettinen A. Childlessness in Finland. In: Kreyenfeld M, Konietzka D, eds. Childlessness in Europe: Contexts, causes, and consequences. Cham: Springer, 2017: 139–158.

10. Saarela J, Skirbekk V. Childlessness and union histories: evidence from Finnish population register data. J Biosoc Sci 2020; 52: 78–96.

11. Balbo N, Billari FC, Mills MC. Fertility in advanced societies: A review of research. Eur J Popul 2013; 29: 1–38.

12. Mills M, Rindfuss RR, McDonald P, Te Velde E. Why do people postpone parenthood? Reasons and social policy incentives. Hum Reprod Update 2011; 17: 848–860.

13. Esping-Andersen, G., 2009. Incomplete revolution: Adapting welfare states to women’s new roles. Polity.

14. Thévenon, O., 2009. Increased women’s labour force participation in Europe: Progress in the work-life balance or polarization of behaviours?. Population, 64(2), pp.235–272.

15. Begall, K. and Mills, M., 2011. The impact of subjective work control, job strain and work–family conflict on fertility intentions: A European comparison. European journal of population= Revue europeenne de demographie, 27(4), p.433.

16. Mills, M. and Blossfeld, H.P., 2006. Globalization, Uncertainty and the Early Life Course: A Theoretical Framework1. In Globalization, uncertainty and youth in society (pp. 1–23). Routledge.

17. Hofmann, B. and Hohmeyer, K., 2013. Perceived economic uncertainty and fertility: Evidence from a labor market reform. Journal of Marriage and Family, 75(2), pp.503–521.

18. Te Velde, E., Habbema, D., Leridon, H. and Eijkemans, M., 2012. The effect of postponement of first motherhood on permanent involuntary childlessness and total fertility rate in six European countries since the 1970s. Human reproduction, 27(4), pp.1179–1183.

19. Merz, E.M. and Liefbroer, A.C., 2012. The attitude toward voluntary childlessness in Europe: Cultural and institutional explanations. Journal of Marriage and Family, 74(3), pp.587–600.

20. Berrington, A., 2004. Perpetual postponers? Women’s, men’s and couple’s fertility intentions and subsequent fertility behaviour. Population trends, 117, pp.9–19.

21. Gray, E., Evans, A. and Reimondos, A., 2013. Childbearing desires of childless men and women: When are goals adjusted?. Advances in life course research, 18(2), pp.141–149.

22. Stephansson O, Dickman PW, Johansson AL, Cnattingius S. The influence of socioeconomic status on stillbirth risk in Sweden. Int J Epidemiol 2001; 30: 1296–1301.

23. Briley DA, Tropf FC, Mills MC. What explains the heritability of completed fertility? Evidence from two large twin studies. Behav Genet 2017; 47: 36–51.

24. Alwan S, Yee IM, Dybalski M, et al. Reproductive decision making after the diagnosis of multiple sclerosis (MS). Mult Scler J 2013; 19: 351–358.

25. Currie J, Schwandt H. Short-and long-term effects of unemployment on fertility. PNAS 2014; 111: 14734–14739.

26. Coomarasamy A, Dhillon-Smith RK, Papadopoulou A, et al. Recurrent miscarriage: evidence to accelerate action. Lancet 2021; 397: 1675–1682.

27. Hull MGR. Epidemiology of infertility and polycystic ovarian disease: endocrinological and demographic studies. Gynecol Endocrinol 1987; 1: 235–245.

28. Bulletti C, Coccia ME, Battistoni S, Borini A. Endometriosis and infertility. J Assist Reprod Genet 2010; 27: 441–447.

29. Purewal, S., Chapman, S., Czuber-Dochan, W., Selinger, C., Steed, H. and Brookes, M.J., 2018. Systematic review: the consequences of psychosocial effects of inflammatory bowel disease on patients′ reproductive health. Alimentary Pharmacology & Therapeutics, 48(11-12), pp.1202–1212.

30. Lee, H.H., Bae, J.M., Lee, B.I., Lee, K.M., Wie, J.H., Kim, J.S., Cho, Y.S., Jung, S.A., Kim, S.W., Choi, H. and Choi, M.G., 2020. Pregnancy outcomes in women with inflammatory bowel disease: a 10-year nationwide population-based cohort study. Alimentary pharmacology & therapeutics, 51(9), pp.861–869.

31. Ferraro, D., Simone, A.M., Adani, G., Vitetta, F., Mauri, C., Strumia, S., Senesi, C., Curti, E., Baldi, E., Santangelo, M. and Montepietra, S., 2017. Definitive childlessness in women with multiple sclerosis: a multicenter study. Neurological Sciences, 38, pp.1453–1459.

32. Power RA, Kyaga S, Uher R, et al. Fecundity of patients with schizophrenia, autism, bipolar disorder, depression, anorexia nervosa, or substance abuse vs their unaffected siblings. JAMA Psychiatry 2013; 70: 22–30.

33. Lawn, R.B., Sallis, H.M., Taylor, A.E., Wootton, R.E., Smith, G.D., Davies, N.M., Hemani, G., Fraser, A., Penton-Voak, I.S. and Munafo, M.R., 2019. Schizophrenia risk and reproductive success: a Mendelian randomization study. Royal Society Open Science, 6(3), p.181049.

34. Mills MC, Tropf FC. The biodemography of fertility: a review and future research frontiers. KZfSS Kölner Zeitschrift für Soziologie und Sozialpsychologie 2015; 67: 397–424.

35. Tanturri ML, Mills MC, Rotkirch A, et al. State-of-the-art report: Childlessness in Europe. Fam Soc 2015; 32: 1–53.

36. Goldscheider, F.K. and Kaufman, G., 1996. Fertility and commitment: Bringing men back in. Population and Development Review, 22, pp.87–99.

37. Pruitt DW, Tsai T. Common medical comorbidities associated with cerebral palsy. Phys Med Rehabil Clin N Am 2009; 20: 453–467.

38. Alias H, Morthy SK, Zakaria SZS, Muda Z, Tamil AM. Behavioral outcome among survivors of childhood brain tumor: a case control study. BMC Pediatr 2020; 20: 1–10.

39. Broberg G, Roll-Hansen N, eds. Eugenics and the welfare state: Norway, Sweden, Denmark, and Finland. East Lansing: Michigan State University Press, 2005.

40. Wei L, MacDonald T, Shimi S. Association between prior appendectomy and/or tonsillectomy in women and subsequent pregnancy rate: a cohort study. Fertil Steril 2016; 106: 1150–1156.

41. Jonasson, JM, Brismar K, Sparen P, et al. Fertility in women with type 1 diabetes: a population-based cohort study in Sweden. Diabetes care 2007. 30: 2271–2276.

42. Miettinen, A., 2010. Voluntary or involuntary childlessness? Socio-demographic factors and childlessness intentions among childless Finnish men and women aged 25-44. Finnish Yearbook of Population Research, pp.5–24.

43. Lloyd ME, Carr M, McElhatton P, Hall GM, Hughes RA. The effects of methotrexate on pregnancy, fertility and lactation. QJM 1999; 92: 551–563.

44. Agarwal, A., Mulgund, A., Hamada, A. and Chyatte, M.R., 2015. A unique view on male infertility around the globe. Reproductive biology and endocrinology, 13(1), pp.1–9.

45. Esteve, A., Cortina, C. and Cabré, A., 2009. Long term trends in marital age homogamy patterns: Spain, 1922-2006. Population, 64(1), pp.173–202.

46. Keizer R, Dykstra PA, Jansen MD. Pathways into childlessness: Evidence of gendered life course dynamics. J Biosoc Sci 2008; 40: 863–878.

47. Furu K, Kieler H, Haglund B, et al. Selective serotonin reuptake inhibitors and venlafaxine in early pregnancy and risk of birth defects: population based cohort study and sibling design. BMJ 2015; 350: 1798.

48. Obel C, Zhu JL, Olsen J, et al. The risk of attention deficit hyperactivity disorder in children exposed to maternal smoking during pregnancy a re-examination using a sibling design. J Child Psychol Psychiatry 2016; 57: 532–537.

49. Spektorowski, A. and Mizrachi, E., 2004. Eugenics and the welfare state in Sweden: The politics of social margins and the idea of a productive society. Journal of Contemporary History, 39(3), pp.333–352.

50. Stern, A.M., 2016. Eugenic nation: Faults and frontiers of better breeding in modern America (Vol. 17). Univ of California Press.

51. Pfaller, B., Sathananthan, G., Grewal, J., Mason, J., D’Souza, R., Spears, D., Kiess, M., Siu, S.C. and Silversides, C.K., 2020. Preventing complications in pregnant women with cardiac disease. Journal of the American College of Cardiology, 75(12), pp.1443–1452.

52. Verweij, R.M., Stulp, G., Snieder, H. and Mills, M.C., 2021. Explaining the Associations of Education and Occupation with Childlessness: The Role of Desires and Expectations to Remain Childless. Population Review, 60(2), pp.166–194.

53. Rybińska, A. and Morgan, S.P., 2019. Childless expectations and childlessness over the life course. Social Forces, 97(4), pp.1571–1602.

54. Meador K, Reynolds MW, Crean S, Fahrbach K, Probst C. Pregnancy outcomes in women with epilepsy: a systematic review and meta-analysis of published pregnancy registries and cohorts. Epilepsy Res 2008; 81: 1–13.

55. Morosow, K. and Kolk, M., 2020. How does birth order and number of siblings affect fertility? A within-family comparison using Swedish register data. European Journal of Population, 36(2), pp.197–233.

56. Surkyn, J. and Lesthaeghe, R., 2004. Value orientations and the second demographic transition (SDT) in Northern, Western and Southern Europe: An update. Demographic research, 3, pp.45–86.

57. Sobotka, T., and L. Toulemon. 2008. Overview Chapter 4: Changing family and partnership behaviour: Common trends and persistent diversity across Europe, Demographic R

58. Jalovaara, M., G. Neyer, G. Andersson, J. Dahlberg, L. Dommermuth, P. Fallesen, and T. Lappegård. 2018. Education, gender, and cohort fertility in the Nordic countries, European Journal of Population 35(3): 563–586. doi:https://doi.org/10.1007/s10680-018-9492-2.

59. Lappegård, T., and M. Rønsen. 2013. Socioeconomic differences in multipartner fertility among Norwegian men, Demography 50(3): 1135–1153. doi:https://doi.org/10.1007/s13524-012-0165-1.

60. Kolk, M. 2019. The relationship between lifecourse accumulated income and childbearing of Swedish men and women born 1940–1970, Stockholm Research Reports in Demography 19. doi:https://doi.org/10.17045/sthlmuni.8283368.v1.

61. Jalovaara, M., L. Andersson and A. Miettinen. (2022). Parity disparity: Educational differences in Nordic fertility across parities and number of reproductive partners, Population Studies, 76(1): 119–136. https://doi.org/10.1080/00324728.2021.1887506.

## References for Methods

62. Kurki et al. FinnGen: Unique genetic insights from combining isolated population and national health register data. medRxiv 2022–03.

63. Ludvigsson JF, Andersson E, Ekbom A. External review and validation of the Swedish national inpatient register. BMC Public Health 2011; 11: 1–16.

64. Kim YM, Cologne JB, Jang E, et al. Causal mediation analysis in nested case-control studies using conditional logistic regression. Biom J 2020; 62: 1939–1959.

