## appendix for "The relationship of early-life diseases with lifetime childlessness: Evidence from Finland and Sweden"

### Table of Contents

1. Theoretical framework for the relationship of diseases with childlessness … 2
2. Review of the literature linking disease to childlessness … 3
3. Nationwide registers from Finland and Sweden … 6
4. Descriptive statistics of fertility and social characteristics in Finland and Sweden … 6
    4.1. Background of childlessness and feritlity in Finland and Sweden … 6
    4.2. Descriptive statistics of childlessness and feritlity in Finland and Sweden … 6
    4.3. Descriptive statistics of social characteristics in Finland and Sweden … 7
5. Disease diagnosis … 10
    5.1. Definition of disease endpoints through ICD-8, ICD-9, and ICD-10 … 10
    5.2. Diseases not included due to virtually complete lack of children … 33
    5.3. Disease prevalence from Finland and Sweden … 34
6. Relationship of disease diagnoses with childlessness … 35
    6.1. Effects of each disease diagnosis … 35
    6.2. Effects by disease category … 36
    6.3. Age-of-onset-dependent effects … 37
    6.4 Stratification by parity for individuals with children … 38
7. Number of unborn children and total fertility rate related to disease diagnoses … 40
8. Mediation effect by singlehood … 46
    8.1. Effects on singlehood … 47
    8.2. Mediation analysis … 49
    8.3. Effects among individuals with registered partner … 50
9. Population-based and sensitivity analyses … 52

Reference … 61

### 1. Theoretical framework for the relationship of diseases with childlessness

In this study childlessness is used to describe individuals that have had no live-born children by the end of their entire reproductive lifespan (age 45 for women; 50 for men). Childlessness is defined in the literature as being both involuntary (e.g., infertility, inability to find a partner) and voluntary or "childfree"(e.g., active choice, preference) (see Text Box Main article).

Diseases can influence individuals' chances of being childless over their lifetime (**Figure S1**). First, diseases may directly influence childlessness through medical conditions that affect the fecundity and mortality of the individual as well as the risk of stillbirth. Second, diseases can impact selection into a partnership which in turn lowers the chance of being partnered and thus having children. Also, some may choose to be childfree due to the concern of genetic transmission of heritable diseases. There could also be concerns about intergenerational transmission of disease and the intergenerational transmission of health and fertility preferences. Finally, diseases are associated with lower socio-economic status, unemployment, and economic uncertainty, which may amplify the psychosocial impact of diseases on childlessness. There are of course multiple non-disease related reasons that individuals postpone or not having children related to factors such as individual choice, work-life reconciliation, housing and economic uncertainty, or inability to find a partner. While individuals' health conditions can also be influenced by their childless status, our study focused on the path from early-life diseases to lifetime chances of being childless.

**Figure S1**: Theoretical framework for the relationship of diseases with childlessness.

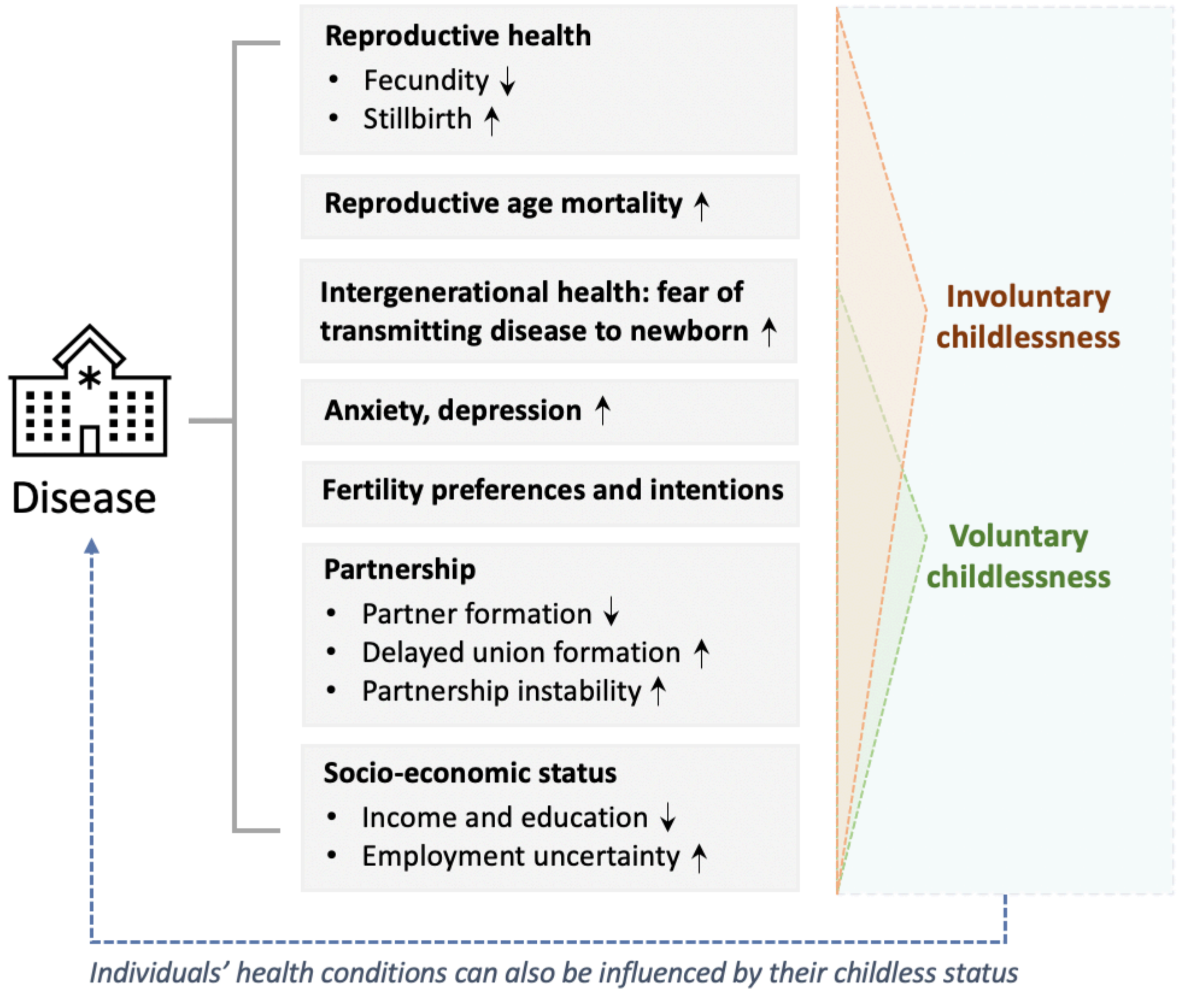

### 2. Review of the literature linking disease to childlessness

**Table S1**: Summary of previous studies linking the relationship of diseases with childlessness.

| **Study** | **DOI** | **Country** | **Data** | **Disease** | **Outcome** | **Review** |
|---|---|---|---|---|---|---|
| Fear and Fertility in Inflammatory Bowel Disease: A Mismatch of Perception and Reality Affects Family Planning Decisions | 10.1002/ibd.20839 | Australia | IBD-database (hospital-based) | Crohn's disease, IBD | Childlessness | No |
| Voluntary childlessness is increased in women with inflammatory bowel disease | 10.1002/ibd.20082 | US | Survey among members of Crohn's and Colitis Foundation of America | Crohn's disease, IBD | Voluntary childlessness, temporary childlessness | No |
| Tubal factor infertility, with special regard to chlamydial salpingitis | -- | NA | NA | tubal factor infertility | Involuntary childlessness | Yes |
| Systematic review: fertility in non-surgically treated inflammatory bowel disease | 10.1111/apt.12478 | NA | NA | Crohn's disease, IBD | Fertility, childlessness | Yes |
| What Factors Might Drive Voluntary Childlessness (VC) in Women with IBD? Does IBD-specific Pregnancy-related Knowledge Matter? | 10.1093/ecco-jcc/jjw078 | UK | Survey among members of Crohn's and Colitits UK | Crohn's disease, IBD | Voluntary childlessness | No |
| Idiopathic impaired spermatogenesis: genetic epidemiology is unlikely to provide a short-cut to better understanding | 10.1093/humupd/dmh045 | NA | NA | impaired spermatogenesis | -- | No |
| Does the level of reproductive knowledge specific to inflammatory bowel disease predict childlessness among women with inflammatory bowel disease? | 10.1155/2015/715354 | Canada | Crohn’s and Colitis Pregnancy Knowledge questionnaire | Crohn's disease, IBD | Childlessness | No |
| Sexual and reproductive issues and inflammatory bowel disease: a neglected topic in men | 10.1097/MEG.0000000000001074 | NA | NA | Crohn's disease, IBD | Childlessness | Yes |
| Pregnancy outcomes in women with inflammatory bowel disease: A 10-year nationwide population-based cohort study | 10.1111/apt.15654 | South Korea | Korean National Health Insurance claims database | Crohn's disease, IBD | Pregnancy outcomes | No |
| Definitive childlessness in women with multiple sclerosis: a multicenter study | 10.1007/s10072-017-2999-1 | Italy | Hospital patients | Multiple Sclerosis | Childlessness | No |
| Sexual health and fertility for individuals with inflammatory bowel disease | 10.3748/wjg.v25.i36.5423 | NA | NA | Crohn's disease, IBD | Childlessness | Yes |
| Inflammatory bowel disease-specific pregnancy knowledge of gastroenterologists against general practitioners and obstetricians | 10.1177/2050640615580893 | Australia | Crohn’s and Colitis Pregnancy Knowledge questionnaire | Crohn's disease / IBD | Childlessness | No |

| | | | | | | |
|---|---|---|---|---|---|---|
| Systematic review: the consequences of psychosocial effects of inflammatory bowel disease on patients' reproductive health | 10.1111/apt.15019 | NA | NA | Crohn's disease / IBD | Childlessness | Yes |
| Endometriosis | 10.1016/S0140-6736(04)17403-5 | NA | NA | Endometriosis | Infertility | Yes |
| ESHRE guideline: management of women with endometriosis | 10.1093/humrep/det457 | NA | NA | Endometriosis | Infertility | Yes |
| Polycystic ovary syndrome | 10.1016/S0140-6736(07)61345-2 | NA | NA | Polycystic ovary syndrome | Infertility | Yes |
| Diagnosis and Treatment of Polycystic Ovary Syndrome: An Endocrine Society Clinical Practice Guideline | /10.1210/jc.2013-2350 | NA | NA | Polycystic ovary syndrome | Infertility | Yes |
| Endometriosis: pathogenesis and treatment | 10.1038/nrendo.2013.255 | NA | NA | Endometriosis | Infertility | Yes |
| Polycystic ovary syndrome: etiology, pathogenesis and diagnosis | 10.1038/nrendo.2010.217 | NA | NA | Polycystic ovary syndrome | Infertility | Yes |
| Coeliac disease | 10.1016/S0140-6736(03)14027-5 | NA | NA | Coeliac disease | Infertility | Yes |
| Polycystic ovary syndrome | 10.1016/S0140-6736(07)61345-2 | NA | NA | Polycystic ovary syndrome | infertility | Yes |
| Diagnosis and Treatment of Polycystic Ovary Syndrome: An Endocrine Society Clinical Practice Guideline | /10.1210/jc.2013-2350 | NA | NA | Polycystic ovary syndrome | infertility | Yes |
| Endometriosis: pathogenesis and treatment | 10.1038/nrendo.2013.255 | NA | NA | Endometriosis | infertility | Yes |
| Polycystic ovary syndrome: etiology, pathogenesis and diagnosis | 10.1038/nrendo.2010.217 | NA | NA | Polycystic ovary syndrome | infertility | Yes |
| Coeliac disease | 10.1016/S0140-6736(03)14027-5 | NA | NA | Coeliac disease | infertility | Yes |
| Family planning in inflammatory bowel disease: childlessness and disease-related concerns among female patients | 10.1097/MEG.0000000000001037 | Germany, Austria, Switzerland | university-hospital based survey | Crohn's disease / IBD | childlessness | No |

| | | | | | | |
|---|---|---|---|---|---|---|
| Risk Factors for Voluntary Childlessness in Men and Women With Inflammatory Bowel Disease | 10.1093/ibd/izac104 | US | survey among patients | Crohn's disease / IBD | voluntary childlessness | No |
| Systematic review: fertility in non-surgically treatedinflammatory bowel disease | 10.1111/apt.12478 | NA | NA | Crohn's disease / IBD | fertility | Yes |
| Systematic review: the consequences of psychosocial effects of inflammatory bowel disease on patients' reproductive health | 10.1111/apt.15019 | NA | NA | Crohn's disease / IBD | voluntary childlessness | Yes |
| Fecundity, pregnancy outcomes, and breastfeeding in patients with inflammatory bowel disease: a large cohort survey | 10.3109/00365521.2013.772229 | Spain | survey among patients | Crohn's disease / IBD | fertility, pregnancy outcomes, breastfeeding | No |
| Increased risk of preterm birth for women with inflammatory bowel disease | 10.1016/0016-5085(90)90617-A | US, North Carolina | self-administered questionnaire | Crohn's disease / IBD, ulcerative colitis | voluntary and involuntary childlessness, infertility, fecund-ability, methods of birth control | No |
| Perception of Reproductive Health in Women with Inflammatory Bowel Disease | 10.1093/ecco-jcc/jjy221 | Malta, Greece, Israel, Italy, Portugal, and Spain | survey in hospitals | Crohn's disease / IBD, ulcerative colitis | -- | No |
| Fertility and Assisted Reproductive Technologies Outcomes of Women with Non-surgically Managed Inflammatory Bowel Diseases: A Systematic Review | 10.1093/ecco-jcc/jjac170 | NA | NA | Crohn's disease / IBD, ulcerative colitis | voluntary childlessness, number of children | Yes |
| The Young Adult with Diabetes: Impact of the Disease on Marriage and Having Children | 10.2337/diacare.8.1.52 | US | small survey (N=50) | diabetis | marriage, having children | No |
| The Relationship Between Common Variant Schizophrenia Liability and Number of Offspring in the UK Biobank | 10.1176/appi.ajp.2018.18020140 | UK | Biobank | schizophrenia | childlessness | No |
| Schizophrenia risk and reproductive success: a Mendelian randomization study | 10.1098/rsos.181049 | UK | Biobank | schizophrenia liability | childlessness | No |
| Early-onset major depressive disorder in men is associated with childlessness | 10.1016/j.jad.2009.10.024 | US | National Epidemiologic Survey on Alcohol and Relationd Conditions | major depressive disorder, bipolar disorder | childlessness, number of children | No |

### 3. Nationwide registers from Finland and Sweden

All Finnish and Swedish register data used in this study can be applied from national data agencies including Statistics Finland (https://www.stat.fi/index_en.html), Population Information System (DVV, https://dvv.fi/en/individuals), Finnish Institute for Health and Welfare (THL, https://thl.fi/en/web/thlfi-en/statistics-and-data/data-and-services/register-descriptions/care-register-for-health-care) and Finnish Cancer Registry (https://cancerregistry.fi/) from Finland, and Statistics Sweden (https://www.scb.se/en/) and National Board of Health and Welfare (Socialstyrelsen, https://www.socialstyrelsen.se/en/) from Sweden.

### 4. Descriptive statistics of fertility and social characteristics in Finland and Sweden

#### 4.1 Background of childlessness and feritlity in Finland and Sweden

Childlessness is interesting to study in the Finnish and Swedish context. Despite declines in period fertility since around 2010, cohort fertility in Finland and Sweden has remained relatively stable and close to replacement levels[1,2,3]. They have had higher levels of gender equity, rates of female labour force participation, and experienced a strong shift in women's educational expansion[4]. Although fertility has remained relatively high, Finland has had the highest levels of childlessness in Europe[5] and a 'parity polarisation' of high levels of childlessness coupled with higher parities in certain groups[4,6]. There is also a parallel 'partner polarisation', or bifurcation of those who never partner with an increase in multi-partner fertility, given that a new union encourages childbearing[3,7].

Finland differs from Sweden in that it has a parity polarisation amongst particularly the lower, but also the medium-educated groups[4]. These differences have been attributed to Finland's skewed national and local sex ratios related to deaths during WWII, mass emigration, low population density, and a larger proportion of women compared to men with tertiary education[5]. The 'two-child family norm' only seems to hold for the higher educated, and childlessness is higher amongst lower educated men and higher educated women[4]. Nordic gender equality and work-family reconciliation therefore seems to disproportionately aid the highly educated groups to reach the two-child norm. Conversely, the fertility patterns of lower educated men and women in Sweden and Finland are more heterogeneous and polarized between childless and higher parity[4].

#### 4.2 Descriptive statistics of childlessness and feritlity in Finland and Sweden

For index individuals in both Finland and Sweden, we considered the numbers of biological children they had by age 45 in women and 50 in men (**Table S2**). Among 1,425,640 index women and 1,119,380 men, 230,198 women and 279,454 men were childless. The proportion of childless individuals – those with no live-born children – was higher in men (25.4%) than women (16.6%), and higher amongst Finns (23.3%) than Swedes (18.7%). Among index persons with children, a two-child parity was most prevalent in men (47.1%) and women (48.9%), amongst both Finns (44.0%) and Swedes (50.8%).

**Table S2**: Descriptive statistics of childlessness and parity for index persons in Finland and Sweden.

| | Finland | | Sweden | |
|---|---|---|---|---|
| | Women | Men | Women | Men |
| Childless | 107,913 (18.8 %) | 128,213 (27.7 %) | 122,285 (14.3%) | 151,241 (23.1 %) |
| One child | 91,114 (15.9 %) | 69,047 (14.9 %) | 115,262 (13.5%) | 91,426 (13.9 %) |
| Two children | 206,436 (36.1 %) | 145,292 (31.4 %) | 377,564 (44.3%) | 250,122 (38.1 %) |
| Three children | 114,966 (20.1 %) | 81,764 (17.6 %) | 177,225 (20.8%) | 118,518 (18.1 %) |
| At least four children | 52,089 (9.1 %) | 39,094 (8.4 %) | 60,786 (7.1%) | 44,663 (6.8 %) |
| Total | 572,518 (100%) | 463,410 (100%) | 853,122 (100%) | 655,970 (100%) |

#### 4.3 Descriptive statistics of social characteristics in Finland and Sweden

To understand the extent to which the social characteristics varied amongst childless individuals and individuals with children in the general population, we considered the highest education level and parental education level in both Finns and Swedes and partnership patterns in Finns only (**Table S3-S4**), depending on data availability.

Overall, in both countries, women were more educated than men (e.g., in Finland, 49.8% of index women completed the first stage of tertiary education while the proportion dropped to 32.5% for index men). Both women and men with the lowest educational attainment (level 0-2; pre-primary education, primary education or first stage of basic education, or lower secondary or second stage of basic education) were more likely to be childless (e.g., in Finland, 24.2% were childless in women and 37.4% in men) than the general population (e.g., in Finland, 18.8% were childless in women and 27.7% in men). Amongst individuals who completed the first stage of tertiary education (levels 5 and 6), men were less likely to be childless (e.g., in Finland, 20.8%) compared to the general population (e.g., in Finland, 27.7%), however, for women, a slightly higher probability of childlessness was observed (e.g., in Finland, 19.2%, compared to 18.8% in the general population).

Across generations, individuals included in these analyses were more educated than their parents (e.g., in Finland, 42.1% of index persons completed the first stage of tertiary education while the proportion dropped to 20.5% in their parents). The education level of parents was also correlated with an individual's childless status. For example, men who had parents with the lowest educational attainment (level 0-2) were more likely to be childless (e.g., in Finland, 28.9%) than the general population (e.g., in Finland, 27.7%). Among index persons whose parents had completed the first stage of tertiary education (levels 5-6), men were less likely to be childless (e.g., in Finland, 25.3%) compared to the general population (e.g., in Finland, 27.7%) while women behaved in the opposite way (e.g., in Finland, 21.3%, compared to 18.8% in the general population).

Regarding the partnership, we only obtained detailed information on partnership types for Finns, for which most of the registered relationships were marriages.

**Table S3**: Descriptive statistics of social characteristics for index persons in Finland.

| Variable | Women | | Men | |
|---|---|---|---|---|
| | With children | Childless | With children | Childless |
| **Parental education level (ISCED 1997)** | | | | |
| Pre-primary education, primary education or first stage of basic education, or lower secondary or second stage of basic education (Level 0-2) | 224,817 (48.4%) | 48,904 (45.3%) | 177,274 (52.9%) | 72,107 (56.2%) |
| Upper secondary education (Level 3) | 141,466 (30.4%) | 32,438 (30.1%) | 92,034 (27.5%) | 33,818 (26.4%) |
| Post-secondary non-tertiary education (Level 4) | 399 (0.1%) | 90 (0.1%) | 95 (0%) | 24 (0%) |
| First stage of tertiary education (Level 5) | 95,139 (20.5%) | 25,710 (23.8%) | 64,076 (19.1%) | 21,735 (17.0%) |
| Second stage of tertiary education (Level 6) | 2,784 (0.6%) | 771 (0.7%) | 1,718 (0.5%) | 529 (0.4%) |
| **Index person's education level (ISCED 1997)** | | | | |
| Pre-primary education, primary education or first stage of basic education, or lower secondary or second stage of basic education (Level 0-2) | 41,466 (8.9%) | 13,232 (12.3%) | 52,867 (15.8%) | 31,625 (24.7%) |
| Upper secondary education (Level 3) | 184,774 (39.8%) | 38,776 (35.9%) | 157,338 (46.9%) | 64,247 (50.1%) |
| Post-secondary non-tertiary education (Level 4) | 7,992 (1.7%) | 1,170 (1.1%) | 5,800 (1.7%) | 965 (0.8%) |
| First stage of tertiary education (Level 5) | 224,983 (48.4%) | 53,117 (49.2%) | 115,052 (34.3%) | 30,419 (23.7%) |
| Second stage of tertiary education (Level 6) | 5,390 (1.2%) | 1,618 (1.5%) | 4,140 (1.2%) | 957 (0.7%) |
| **Partnership pattern** | | | | |
| No partner | 69,263 (14.9%) | 70,300 (65.1%) | 45,308 (13.5%) | 93,067 (72.6%) |
| Ever-married | 395,127 (85.0%) | 37,169 (34.4) | 289,834 (86.5%) | 34,719 (27.1%) |
| Registered partnership | 215 (0.05%) | 444 (0.41%) | 55 (0.02%) | 427 (0.33%) |

**Table S4**: Descriptive statistics of social characteristics for index persons in Sweden.

| Variable | Women | | Men | |
|---|---|---|---|---|
| | With children | Childless | With children | Childless |
| **Parental education level (ISCED 1997)** | | | | |
| Pre-primary education, primary education or first stage of basic education, or lower secondary or second stage of basic education (Level 0-2) | 227,829 (31.2%) | 38,260 (31.3%) | 179,183 (35.5%) | 58,593 (38.7%) |
| Upper secondary education (Level 3) | 331,431 (45.3%) | 52,960 (43.3%) | 217,967 (43.2%) | 62,713 (41.5%) |
| Post-secondary non-tertiary education (Level 4) | 13,745 (1.9%) | 2,460 (2.0%) | 7,705 (1.5%) | 2,131 (1.4%) |
| First stage of tertiary education (Level 5) | 149,146 (20.4%) | 26,934 (22.0%) | 94,453 (18.7%) | 26,311 (17.4%) |
| Second stage of tertiary education (Level 6) | 8,686 (1.2%) | 1,671 (1.4%) | 5,421 (1.1%) | 1,493 (1.0%) |
| **Index person's education level (ISCED 1997)** | | | | |
| Pre-primary education, primary education or first stage of basic education, or lower secondary or second stage of basic education (Level 0-2) | 53,253 (7.3%) | 14,833 (12.1%) | 71,808 (14.2%) | 31,105 (20.6%) |
| Upper secondary education (Level 3) | 362,598 (49.6%) | 55,670 (45.5%) | 274,402 (54.4%) | 80,326 (53.1%) |
| Post-secondary non-tertiary education (Level 4) | 39,626 (5.4%) | 7,938 (6.5%) | 49,607 (9.8%) | 13,314 (8.8%) |
| First stage of tertiary education (Level 5) | 267,718 (36.6%) | 42,304 (34.6%) | 101,521 (20.1%) | 24,897 (16.5%) |
| Second stage of tertiary education (Level 6) | 7,642 (1.0%) | 1,540 (1.3%) | 7,391 (1.5%) | 1,599 (1.1%) |

### 5. Disease diagnosis

#### 5.1. Definition of disease endpoints through ICD-8, ICD-9, and ICD-10

Disease endpoints were defined by clinical expert groups, through International Classification of Diseases (ICD) codes of version 8 (1969-1986), 9 (1987-1995 in Finland, 1987-1996 in Sweden), and 10 (1996-2018 in Finland, 1997-2018 in Sweden).

To remove highly correlated disease diagnoses, we estimated tetrachoric correlations between every two disease endpoints using individual-level data and only kept the one with the highest prevalence if a group of disease diagnoses had tetrachoric correlations higher than 0.7. After removing highly correlated disease diagnoses, we considered 414 disease diagnoses for which we had more than 30 affected individuals in the sibling-based analysis for each sex, in Finland or Sweden.

Disease categories were defined based on the ICD coding system. Among 16 disease categories, infectious-parasitic diseases and congenital anomalies were defined only in Finland while all remaining categories were defined in both countries. Number of diseases endpoints defined for each category is presented in **Figure S2**.

**Figure S2**. Number of disease endpoints by disease category.

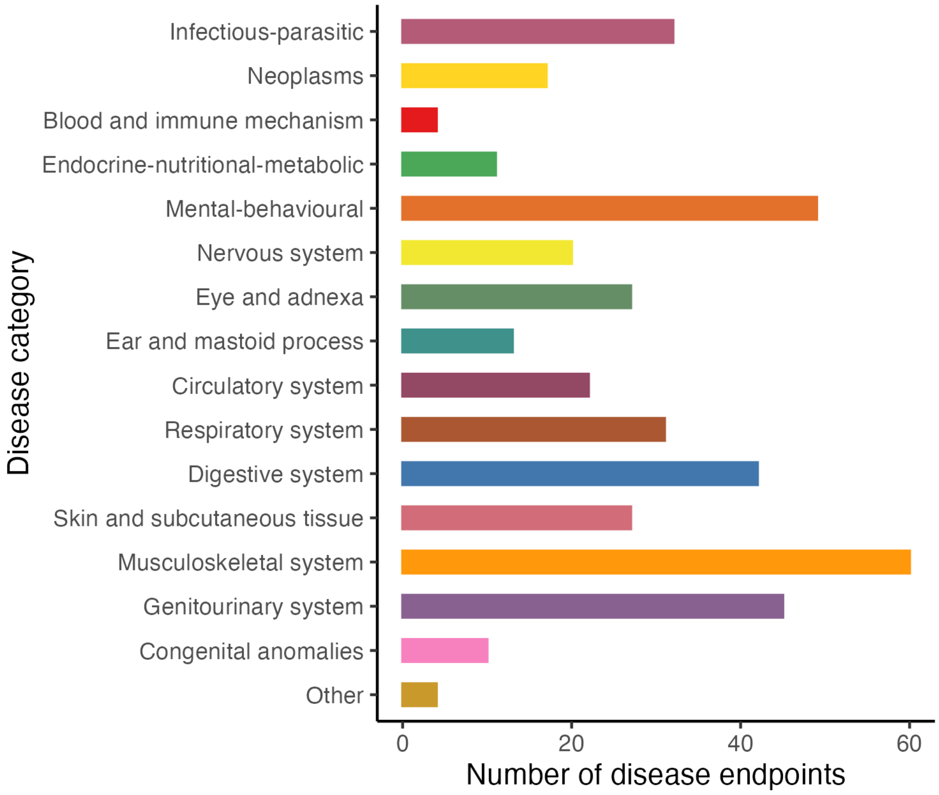

**Table S5**: Definition of 414 disease endpoints (16 categories) through ICD-8, ICD-9, and ICD-10.

| Disease | ICD-8 | ICD-9 | ICD-10 | Category | Population |
|---|---|---|---|---|---|
| Respiratory TBC, ICD-8 codes not specifying whether confirmed bacteriologically or histologically | 011, 012 | -- | -- | Infectious-parasitic | Finland |
| Other salmonella infections | 003 | 003 | A02 | Infectious-parasitic | Finland |
| Viral and other specified intestinal infections | 0088, 0089 | 008 | A08 | Infectious-parasitic | Finland |
| Diarrhoea and gastroenteritis of presumed infectious origin | 009 | 009 | A09 | Infectious-parasitic | Finland |
| Tuberculosis | 010, 011, 012, 013, 014, 015, 016, 017, 018 | 010, 011, 012, 013, 014, 015, 016, 017, 018 | A15, A16, A17, A18, A19 | Infectious-parasitic | Finland |
| Scarlet fever | 0341 | 0341 | A38 | Infectious-parasitic | Finland |
| Meningococcal infection | 036 | 036 | A39 | Infectious-parasitic | Finland |
| Other septicaemia | 03810, 03820, 03880, 03888, 03897, 03899 | 0381, 0382, 0383, 0384, 0388 | A41 | Infectious-parasitic | Finland |
| Erysipelas | 035 | 035 | A46 | Infectious-parasitic | Finland |
| Bacterial infection, other or unspecified | 039 | 040, 041 | A48, A49 | Infectious-parasitic | Finland |
| Gonococcal infection | 098 | 098 | A54 | Infectious-parasitic | Finland |
| Anogenital herpesviral [herpes simplex] infection | 05402 | 0541A | A60 | Infectious-parasitic | Finland |
| Other predominantly sexually transmitted diseases, not elsewhere classified | 0990, 0999 | 0990A, 0993A, 0994A, 0998X, 0999X | A63, A64 | Infectious-parasitic | Finland |
| Other diseases caused by chlamydiae | -- | 0769A, 0769B, 0769C, 0769D, 0769E, 0769F, 0769X | A74 | Infectious-parasitic | Finland |
| Acute poliomyelitis | 040, 041, 042, 043, 044, 045 | 045 | A80 | Infectious-parasitic | Finland |
| Viral meningitis | 045 | 047 | A87 | Infectious-parasitic | Finland |
| Arthropod-borne viral fevers and viral haemorrhagic fevers | 061, 067, 068 | 065, 066, 0786, 0787 | A90, A91, A92, A93, A94, A95, A96, A98, A99 | Infectious-parasitic | Finland |
| Herpesviral [herpes simplex] infections | 05400, 05401, 05403, 05404, 05405, 05406, 05407, 05409 | 0540A, 0541B, 0542A, 0542B, 0543A, 0543B, 0544A, 0544B, 0544C, 0544D, 0544X, 0545A, 0546A, 0547A, 0547X, 0548X, 0549X | B00 | Infectious-parasitic | Finland |
| Viral infections characterized by skin and mucous membrane lesions | 0790, 0791 | 0780A, 0781A, 0781B, 0781D, 0783A | B00, B01, B02, B03, B04, B05, B06, B07, B08, B09 | Infectious-parasitic | Finland |
| Varicella [chickenpox] | 052 | 052 | B01 | Infectious-parasitic | Finland |
| Zoster [herpes zoster] | 053 | 053 | B02 | Infectious-parasitic | Finland |
| Measles | 055 | 055 | B05 | Infectious-parasitic | Finland |

| | | | | | |
|---|---|---|---|---|---|
| Viral hepatitis | 070 | 070 | B15, B16, B17, B18, B19 | Infectious-parasitic | Finland |
| Other viral diseases | 075, 0792, 0799 | 0785, 0787, 0788 | B25, B26, B27, B30, B33, B34 | Infectious-parasitic | Finland |
| Mumps | 072 | 072 | B26 | Infectious-parasitic | Finland |
| Infectious mononucleosis | 075 | 075 | B27 | Infectious-parasitic | Finland |
| Mycoses | 110, 111, 112, 113, 114, 115, 116, 117 | 110, 111, 112, 114, 115, 116, 117, 118 | B35, B36, B37, B38, B39, B40, B41, B42, B43, B44, B45, B46, B47, B48, B49 | Infectious-parasitic | Finland |
| Candidiasis | 112 | 112 | B37 | Infectious-parasitic | Finland |
| Pediculosis, acariasis and other infestations | 132, 133, 134 | 132, 133, 134 | B85, B86, B87, B88, B89 | Infectious-parasitic | Finland |
| Sequelae of other and unspecified infectious and parasitic diseases | 066, 079, 074 | 139 | B94 | Infectious-parasitic | Finland |
| Bacterial, viral, and other infectious agents | 0340, 074, 078 | 079, 0340 | B95, B96, B97 | Infectious-parasitic | Finland |
| Other infectious diseases | 13608, 13609 | 1368, 1369 | B99 | Infectious-parasitic | Finland |
| Malignant neoplasm of testis | 186 | 186 | C62 | Neoplasms | Finland |
| Malignant neoplasm of brain | 191 | 191 | C71 | Neoplasms | Finland |
| Secondary and unspecified malignant neoplasm of lymph nodes | 196 | -- | C77 | Neoplasms | Finland |
| Lymphoid leukaemia | 204 | 204 | C91 | Neoplasms | Finland, Sweden |
| Myeloid leukaemia | 205 | 205 | C92 | Neoplasms | Finland, Sweden |
| Carcinoma in situ of uteri, other/unspecified | -- | 233B | D067, D069 | Neoplasms | Sweden |
| Benign neoplasm: other/unspecified salivary gland | 21020 | 210C | D117, D119 | Neoplasms | Sweden |
| Benign neoplasm: Long bones of lower limb | 21306 | 2137 | D162 | Neoplasms | Finland |
| Benign neoplasm: Long bones of lower limb | 21306 | 213H | D162 | Neoplasms | Sweden |
| Benign lipomatous neoplasm of other sites/unspecified | 214 | 214 | D177, D179 | Neoplasms | Finland |
| Haemangioma, any site | 22700 | 2280 | D180 | Neoplasms | Finland |
| Haemangioma, any site | 22700 | 228A | D180 | Neoplasms | Sweden |
| Melanocytic naevi, other sites/unspecified | -- | 216W, 216X | D229 | Neoplasms | Sweden |
| Benign neoplasm: Skin of other and unspecified parts of face | 21604 | 2163B, 2163C, 2163D, 2163E, 2163F, 2163G, 2163X | D233 | Neoplasms | Finland |
| Benign neoplasm: Skin of trunk | 21606 | 2165B, 2165C, 2165D, 2165E, 2165F, 2165G, 2165X | D235 | Neoplasms | Finland |

| | | | | | |
|---|---|---|---|---|---|
| Benign neoplasm: Skin, unspecified | 21600, 21609, 2161, 2162, 2168, 2169 | 2168B, 2168C, 2168D, 2168E, 2168F, 2168G, 2168X, 2169B, 2169C, 2169D, 2169E, 2169F, 2169G, 2169X | D239 | Neoplasms | Finland |
| Leiomyoma of uterus | 21899 | 218 | D25 | Neoplasms | Finland |
| Benign neoplasm: Vulva | 2212 | -- | D280 | Neoplasms | Finland |
| Benign neoplasm: Pituitary gland, craniopharyngeal duct | 22620 | 227D | D352, D353 | Neoplasms | Sweden |
| Iron deficiency anaemia secondary to blood loss (chronic) | 28000 | 2800 | D500 | Blood and immune mechanism | Finland |
| Iron deficiency anaemia secondary to blood loss (chronic) | 28000 | 280 | D500 | Blood and immune mechanism | Sweden |
| Other and unspecified iron deficiency | 28008, 28009 | 2801, 2808, 2809 | D501, D508, D509 | Blood and immune mechanism | Finland |
| Other and unspecified iron deficiency | 28008, 28009 | 280 | D501, D508, D509 | Blood and immune mechanism | Sweden |
| Allergic purpura | 2870 | 2870 | D690 | Blood and immune mechanism | Finland |
| Allergic purpura | 2870 | 287A | D690 | Blood and immune mechanism | Sweden |
| Sarcoidosis | 135 | 135 | D86 | Blood and immune mechanism | Finland, Sweden |
| Hypothyroidism, other/unspecified | 244 | 2448X, 2449 | E038, E039 | Endocrine-nutritional-metabolic | Finland |
| Hypothyroidism, other/unspecified | 244 | 244W, 244X | E038, E039 | Endocrine-nutritional-metabolic | Sweden |
| Nontoxic multinodular goitre | 241 | 2411 | E042 | Endocrine-nutritional-metabolic | Finland |
| Nontoxic multinodular goitre | 241 | 241B | E042 | Endocrine-nutritional-metabolic | Sweden |
| Thyrotoxicosis with diffuse goitre | 2420 | 2420 | E050 | Endocrine-nutritional-metabolic | Finland |
| Thyrotoxicosis with diffuse goitre | 2420 | 242A | E050 | Endocrine-nutritional-metabolic | Sweden |
| Thyroiditis, ILD-related definition | 245 | 2448A | E06, E0380 | Endocrine-nutritional-metabolic | Finland |
| Type 1 diabetes | 250 (onset before age 40) | 2500B, 2501B, 2502B, 2503B, 2504B, 2505B, 2506B, 2507B, 2508B | E10 | Endocrine-nutritional-metabolic | Finland |
| Type 1 diabetes | 250 (onset before age 40) | 250 (onset before age 40) | E10 | Endocrine-nutritional-metabolic | Sweden |
| Type 2 diabetes | -- | 2500A, 2501A, 2502A, 2503A, 2504A, 2505A, 2506A, 2507A, 2508A | E11 | Endocrine-nutritional-metabolic | Finland |
| Hypoglycaemia, other or unspecified | 25101 | 251C | E161, E162 | Endocrine-nutritional-metabolic | Sweden |
| Hyperprolactinaemia | -- | 2531A | E221 | Endocrine-nutritional-metabolic | Finland |
| Polycystic ovarian syndrome | 25690 | 2564 | E282 | Endocrine-nutritional-metabolic | Finland |

| | | | | | |
|---|---|---|---|---|---|
| Polycystic ovarian syndrome | 25690 | 256E | E282 | Endocrine-nutritional-metabolic | Sweden |
| Other specified/unspecified nutritional deficiencies | 26998, 26999 | 2693, 2698, 2699 | E638, E639 | Endocrine-nutritional-metabolic | Finland |
| Other specified/unspecified nutritional deficiencies | 26998, 26999 | 269D, 269W, 269X | E638, E639 | Endocrine-nutritional-metabolic | Sweden |
| Obesity, other/unspecified | 27799 | 2780 | E668, E669 | Endocrine-nutritional-metabolic | Finland |
| Obesity, other/unspecified | 27799 | 278A | E668, E669 | Endocrine-nutritional-metabolic | Sweden |
| Other mental disorders due to brain damage and dysfunction and to physical disease | 2940, 2941, 2942, 2948, 2949, 309 | 2938 | F06 | Mental-behavioural | Finland |
| Other mental disorders due to brain damage and dysfunction and to physical disease | 2940, 2941, 2942, 2948, 2949, 309 | 293W | F06 | Mental-behavioural | Sweden |
| Acute alcohol intoxication | -- | 3050 | F100 | Mental-behavioural | Finland |
| Acute alcohol intoxication | -- | 305A | F100 | Mental-behavioural | Sweden |
| Alcohol dependence | 303 | 303 | F102 | Mental-behavioural | Finland, Sweden |
| Mental and behavioural disorders due to opioids | 3040, 3041 | 304A | F11 | Mental-behavioural | Sweden |
| Mental and behavioural disorders due to cannabinoids | 3045 | 3052, 3043 | F12 | Mental-behavioural | Finland |
| Mental and behavioural disorders due to cannabinoids | 3045 | 304D | F12 | Mental-behavioural | Sweden |
| Mental and behavioural disorders due to sedatives or hypnotics | 3042, 3043 | 3041, 3054 | F13 | Mental-behavioural | Finland |
| Mental and behavioural disorders due to sedatives or hypnotics | 3042, 3043 | 304B | F13 | Mental-behavioural | Sweden |
| Mental and behavioural disorders due to cocaine | 3044 | 304C | F14 | Mental-behavioural | Sweden |
| Mental and behavioural disorders due to use of other stimulants, including caffeine | 3046 | 3044, 3057 | F15 | Mental-behavioural | Finland |
| Mental and behavioural disorders due to use of other stimulants, including caffeine | 3046 | 304E | F15 | Mental-behavioural | Sweden |
| Mental and behavioural disorders due to hallucinogens | 3047 | 304F | F16 | Mental-behavioural | Sweden |
| Mental and behavioural disorders due to multiple drug | 3048, 3049 | 3049, 3059 | F19 | Mental-behavioural | Finland |

| | | | | | |
|---|---|---|---|---|---|
| use and use of other psychoactive substances | | | | | |
| Mental and behavioural disorders due to multiple drug use and use of other psychoactive substances | 3048, 3049 | 304X, 305X | F19 | Mental-behavioural | Sweden |
| Schizophrenia | 2950, 2951, 2952, 2953, 2954, 2955, 2956, 2958, 2959 | 2951, 2952, 2953, 2956, 2959 | F20 | Mental-behavioural | Finland |
| Schizophrenia | 2950, 2951, 2952, 2953, 2954, 2955, 2956, 2958, 2959 | 295B, 295C, 295D, 295G, 295X | F20 | Mental-behavioural | Sweden |
| Persistent delusional disorders | 297 | 2971 | F22 | Mental-behavioural | Finland |
| Acute and transient psychotic disorders | 2980, 2981, 2982, 2983 | 298 | F23 | Mental-behavioural | Finland |
| Schizoaffective disorder | 2957 | 2957 | F25 | Mental-behavioural | Finland |
| Schizoaffective disorder | 2957 | 295H | F25 | Mental-behavioural | Sweden |
| Other and unspecified nonorganic psychotic disorders | 29999 | 2989 | F28, F29 | Mental-behavioural | Finland |
| Other and unspecified nonorganic psychotic disorders | 29999 | 298X | F28, F29 | Mental-behavioural | Sweden |
| Mood disorders | 296, 29800, 3004, 30110 | 296, 3004A, 3011D | F30, F31, F32, F33, F34, F38, F39 | Mental-behavioural | Finland |
| Bipolar affective disorders | 2961, 2962, 2963, 2968 | 2962, 2963, 2964, 2967 | F31 | Mental-behavioural | Finland |
| Bipolar affective disorders | 2961, 2962, 2963, 2968 | 296C, 296D, 296E | F31 | Mental-behavioural | Sweden |
| Persistent mood disorders | 3004 | 3004 | F34 | Mental-behavioural | Finland |
| Persistent mood disorders | 3004 | 300E | F34 | Mental-behavioural | Sweden |
| Other and unspecified mood [affective] disorders | 29699 | -- | F38, F39 | Mental-behavioural | Sweden |
| Phobic anxiety disorders | 3002 | 3002C, 3002D, 3002X | F40 | Mental-behavioural | Finland |
| Phobic anxiety disorders | 3002 | 300C | F40 | Mental-behavioural | Sweden |
| Anxiety disorders | 3000, 3001, 3002, 30030, 3005, 3006, 3007, 3008, 3009, 305, 30680, 30799 | 3000, 3001, 3002, 3003, 3006, 3007, 3008, 3009, 3078A, 309 | F40, F41, F42, F43, F44, F45, F48 | Mental-behavioural | Finland |
| Panic disorder | -- | 3000B, 3002B | F410 | Mental-behavioural | Finland |
| Panic disorder | -- | 300A, 300C | F410 | Mental-behavioural | Sweden |
| Generalized anxiety disorder | -- | 3000C | F411 | Mental-behavioural | Finland |
| Generalized anxiety disorder | -- | 300A | F411 | Mental-behavioural | Sweden |
| Other anxiety disorders | 3000 | 3000A | F412, F413, F418, F419 | Mental-behavioural | Finland |
| Other anxiety disorders | 3000 | 300A | F412, F413, F418, F419 | Mental-behavioural | Sweden |
| Obsessive-compulsive disorder | 3003 | 3003 | F42 | Mental-behavioural | Finland |

| | | | | | |
|---|---|---|---|---|---|
| Obsessive-compulsive disorder | 3003 | 300D | F42 | Mental-behavioural | Sweden |
| Other reaction to severe stress, and adjustment disorders | -- | 309 | F430, F432, F438, F439 | Mental-behavioural | Finland, Sweden |
| Post-traumatic stress disorder | -- | 309X | F431 | Mental-behavioural | Sweden |
| Dissociative [conversion] disorders | 3001 | 3001 | F44 | Mental-behavioural | Finland |
| Dissociative [conversion] disorders | 3001 | 300B | F44 | Mental-behavioural | Sweden |
| Somatoform disorder | 305 | 3007, 3008 | F45 | Mental-behavioural | Finland |
| Somatoform disorder | 305 | 300H, 300W | F45 | Mental-behavioural | Sweden |
| Other neurotic disorders | 3005, 3006, 3007, 3008, 3009 | 3006, 3009 | F48 | Mental-behavioural | Finland |
| Other neurotic disorders | 3005, 3006, 3007, 3008, 3009 | 300G, 300X | F48 | Mental-behavioural | Sweden |
| Anorexia (incl.atypical) | 784 | 3071, 7830 | F500, F501 | Mental-behavioural | Finland |
| Anorexia (incl.atypical) | 784 | 307B, 783A | F500, F501 | Mental-behavioural | Sweden |
| Bulimia nervosa (incl. atypical) | -- | 307F | F502, F503 | Mental-behavioural | Sweden |
| Other eating disorders | -- | 307F | F504, F505, F508, F509 | Mental-behavioural | Sweden |
| Nonorganic sleeping disorders | 30640 | 3074 | F51 | Mental-behavioural | Finland |
| Paranoid personality disorder | 3010 | 3010 | F600 | Mental-behavioural | Finland |
| Schizoid personality disorder | 3012 | 3012 | F601 | Mental-behavioural | Finland |
| Dissocial personality disorder | 3017 | 3017 | F602 | Mental-behavioural | Finland |
| Emotionally unstable personality disorder | 3013 | 3018D | F603 | Mental-behavioural | Finland |
| Emotionally unstable personality disorder | 3013 | 301W | F603 | Mental-behavioural | Sweden |
| Anxious personality disorder | -- | 301W | F606 | Mental-behavioural | Sweden |
| Dependent personality disorder | 3016 | 3016 | F607 | Mental-behavioural | Finland |
| Other specified and unspecified personality disorders | 30188, 3019 | 3018B, 3018E, 3018X | F608, F609 | Mental-behavioural | Finland |
| Other specified and unspecified personality disorders | 30188, 3019 | 301W | F608, F609 | Mental-behavioural | Sweden |
| Mild intellectual disability | 311 | 3170 | F70 | Mental-behavioural | Finland |
| Mild intellectual disability | 311 | 317 | F70 | Mental-behavioural | Sweden |
| Moderate intellectual disability | 312 | 3180 | F71 | Mental-behavioural | Finland |
| Moderate intellectual disability | 312 | 318A | F71 | Mental-behavioural | Sweden |
| Severe intellectual disability | 313 | 3181 | F72 | Mental-behavioural | Finland |

| | | | | | |
|---|---|---|---|---|---|
| Severe intellectual disability | 313 | 318B | F72 | Mental-behavioural | Sweden |
| Profound intellectual disability | 314 | 3182 | F73 | Mental-behavioural | Finland |
| Profound intellectual disability | 314 | 318C | F73 | Mental-behavioural | Sweden |
| Other and unspecified intellectual disability | 315 | 3199 | F78, F79 | Mental-behavioural | Finland |
| Other and unspecified intellectual disability | 315 | 319 | F78, F79 | Mental-behavioural | Sweden |
| Specific development disorders of scholastic skills | 3061 | 3150, 3151, 3152 | F81 | Mental-behavioural | Finland |
| Behavioural disorders | 30899 | 3120A, 3123D, 3138A | F91 | Mental-behavioural | Finland |
| Other behavioural and emotional disorders with onset usually occurring in childhood and adolescence | 3066, 3067 | 3076, 3077, 3075D, 3075C | F98 | Mental-behavioural | Finland |
| Other behavioural and emotional disorders with onset usually occurring in childhood and adolescence | 3066, 3067 | 307F, 307G, 307H | F98 | Mental-behavioural | Sweden |
| Bacterial meningitis | 320 | 320 | G00, G01 | Nervous system | Finland, Sweden |
| Other encephalitis | 32308, 32309 | 3238B, 3238X | G041, G048, G049, G051, G052, G058 | Nervous system | Finland |
| Multiple Sclerosis | 34099 | 340 | G35 | Nervous system | Finland, Sweden |
| Epilepsy | 3450, 3451, 3453, 3459 | 3450, 3451, 3454, 3455, 3456, 3457, 3458, 3459 | G40 | Nervous system | Finland |
| Epilepsy | 3450, 3451, 3453, 3459 | 345J, 345K, 345L, 345M, 345N, 345W, 345X | G40 | Nervous system | Sweden |
| Status epilepticus | 34520 | 3452, 3453 | G41 | Nervous system | Finland |
| Status epilepticus | 34520 | 345P, 345Q | G41 | Nervous system | Sweden |
| Migraine | 34609 | 346 | G43 | Nervous system | Finland, Sweden |
| Other headache syndromes | 34601 | -- | G44 | Nervous system | Finland, Sweden |
| Other sleep disorders | -- | 3478, 3074 | G470, G471, G472, G478, G479 | Nervous system | Finland |
| Sleep apnoea | -- | 3472 | G473 | Nervous system | Finland |
| Sleep apnoea | -- | 347 | G473 | Nervous system | Sweden |
| Bell palsy | 350 | 3510 | G510 | Nervous system | Finland |
| Bell palsy | 350 | 351A | G510 | Nervous system | Sweden |
| Carpal tunnel syndrome | -- | 3540 | G560 | Nervous system | Finland |
| Carpal tunnel syndrome | -- | 354A | G560 | Nervous system | Sweden |
| Lesion of radial nerve | 35708 | 3543 | G563 | Nervous system | Finland |
| Lesion of sciatic nerve | 35399 | 3550 | G570 | Nervous system | Finland |
| Other and unspecified mononeuropathies, also in other diseases | 35708 | 3559 | G588, G589, G59 | Nervous system | Finland |
| Muscular dystrophy | 33030,33090 | 3591 | G710 | Nervous system | Finland |
| Muscular dystrophy | 33030,33090 | 359B | G710 | Nervous system | Sweden |

| | | | | | |
|---|---|---|---|---|---|
| Cerebral palsy | 343 | 343 | G80 | Nervous system | Finland |
| Hemiplegia | 34400 | 342 | G81 | Nervous system | Finland, Sweden |
| Other and unspecified paralytic syndromes | 34408,34409 | 344 | G838, G839 | Nervous system | Finland, Sweden |
| Disorders of brain, other and unspecified | 34798,34799 | 3488,3489 | G938, G939, G948 | Nervous system | Finland |
| Disorders of brain, other and unspecified | 34798,34799 | 348W,348X | G938, G939, G948 | Nervous system | Sweden |
| Postprocedural disorders of nervous system | -- | 3490, 997 | G97 | Nervous system | Finland |
| Unilateral visual impairment (ICD-8) | 3792, 3793 | -- | -- | Eye and adnexa | Finland |
| Chalazion | 3780 | 3732 | H001 | Eye and adnexa | Finland |
| Disorders of lacrimal system | -- | -- | H04 | Eye and adnexa | Finland |
| Stenosis and insufficiency of lacrimal passages | 36802 | 3755 | H045 | Eye and adnexa | Finland |
| Conjunctivitis (acute, non-atopic) | 36000,36008,36009 | 3720 | H100, H102, H103, H105, H108, H109, H131, H132 | Eye and adnexa | Finland |
| Conjunctivitis (acute, non-atopic) | 36000,36008,36009 | 372A | H100, H102, H103, H105, H108, H109, H131, H132 | Eye and adnexa | Sweden |
| Corneal ulcer | 36300 | 3700 | H160 | Eye and adnexa | Finland |
| Other keratitis | 36301, 36302, 36308, 36309, 36390, 36391, 36392, 36393, 36394, 36398, 36399, 36001, 36002, 36905 | 3702, 3703, 3705, 3706 | H161, H162, H163, H164, H168, H169 | Eye and adnexa | Finland |
| Other keratitis | 36301, 36302, 36308, 36309, 36390, 36391, 36392, 36393, 36394, 36398, 36399, 36001, 36002, 36905 | 370C, 370D, 370F, 370G | H161, H162, H163, H164, H168, H169 | Eye and adnexa | Sweden |
| Keratoconus | 37103 | 371G | H186 | Eye and adnexa | Sweden |
| Acute and subacute iridocyclitis | 36400, 36401 | 3640 | H200 | Eye and adnexa | Finland |
| Acute and subacute iridocyclitis | 36400, 36401 | 364A | H200 | Eye and adnexa | Sweden |
| Other cataract | 37400, 37401, 37403, 37404, 37405, 37406, 37407, 37408, 37409 | 3660, 3662, 3663, 3664, 3665, 3668, 3669 | H26 | Eye and adnexa | Finland |
| Other cataract | 37400, 37401, 37403, 37404, 37405, 37406, 37407, 37408, 37409 | 366A, 366C, 366D, 366E, 366F, 366W, 366X | H26 | Eye and adnexa | Sweden |
| Aphakia | 37872 | 3793A | H270 | Eye and adnexa | Finland |
| Chorioretinal inflammation | 365, 36700 | 3630, 3631, 3632 | H30, H320 | Eye and adnexa | Finland |
| Retinal detachment with retinal break | 37602 | 361A | H330 | Eye and adnexa | Sweden |
| Other, unspecified, and serous retinal detachments | 37600, 37601, 37603, 37609 | 3618, 3619 | H332, H335 | Eye and adnexa | Finland |
| Other, unspecified, and serous retinal detachments | 37600, 37601, 37603, 37609 | 361W, 361X | H332, H335 | Eye and adnexa | Sweden |
| Other and unspecified glaucoma | 3752, 37598, 37599 | 3656X, 3656C, 3658, 3659 | H408, H409 | Eye and adnexa | Finland |
| Optic neuritis | 36702, 36703, 36709 | 3773 | H46 | Eye and adnexa | Finland |

| | | | | | |
|---|---|---|---|---|---|
| Convergent concomitant strabismus | 37300 | 3780 | H500 | Eye and adnexa | Finland |
| Convergent concomitant strabismus | 37300 | 378A | H500 | Eye and adnexa | Sweden |
| Divergent concomitant strabismus | 37301 | 3781 | H501 | Eye and adnexa | Finland |
| Divergent concomitant strabismus | 37301 | 378B | H501 | Eye and adnexa | Sweden |
| Vertical strabismus | 37302 | 3783 | H502 | Eye and adnexa | Finland |
| Heterophoria | 37303 | 3784 | H505 | Eye and adnexa | Finland |
| Heterophoria | 37303 | 378E | H505 | Eye and adnexa | Sweden |
| Hypermetropia | 37001 | 3670 | H520 | Eye and adnexa | Finland |
| Myopia | 37002 | 3671 | H521 | Eye and adnexa | Finland |
| Astigmatism | 37000 | 3672 | H522 | Eye and adnexa | Finland |
| Anisometropia and aniseikonia | 37002 | 3673 | H523 | Eye and adnexa | Finland |
| Presbyopia | 37002 | 3674 | H524 | Eye and adnexa | Finland |
| Visual disturbances | -- | -- | H53 | Eye and adnexa | Finland |
| Amblyopia ex anopsia | 37720 | 3680 | H530 | Eye and adnexa | Finland |
| Otitis externa | 380 | -- | H60 | Ear and mastoid process | Sweden |
| Otitis externa, unspecified | 38002, 38003, 38004, 38008, 38009 | 3801A, 3801B | H609 | Ear and mastoid process | Finland |
| Otitis externa, unspecified | 38002, 38003, 38004, 38008, 38009 | 380B | H609 | Ear and mastoid process | Sweden |
| Perichondritis of external ear | 38798 | 3800 | H610 | Ear and mastoid process | Finland |
| Nonsuppurative otitis media | -- | 381 | H65 | Ear and mastoid process | Finland, Sweden |
| Acute suppurative otitis media | 3810 | 3820 | H660 | Ear and mastoid process | Finland |
| Acute suppurative otitis media | 3810 | 382A | H660 | Ear and mastoid process | Sweden |
| Otitis media, unspecified | 3819 | -- | H669 | Ear and mastoid process | Finland |
| Eustachian salpingitis and obstruction | 38400, 38401, 38402 | 3816 | H680, H681 | Ear and mastoid process | Finland |
| Eustachian salpingitis and obstruction | 38400, 38401, 38402 | 381G | H680, H681 | Ear and mastoid process | Sweden |
| Acute mastoiditis | 3820, 3830 | 3830 | H700 | Ear and mastoid process | Finland |
| Chronic mastoiditis | 3821, 3831 | 3831 | H701 | Ear and mastoid process | Finland |
| Cholesteatoma of middle ear | 38700 | 3853 | H71 | Ear and mastoid process | Finland |
| Otosclerosis | 38699 | 387 | H80 | Ear and mastoid process | Finland |
| Sensorineural hearing loss | -- | 3891 | H903, H904, H905 | Ear and mastoid process | Finland |
| Sensorineural hearing loss | -- | 389B | H903, H904, H905 | Ear and mastoid process | Sweden |
| Other specified/unspecified hearing loss | 389 | 3898,3899 | H918, H919 | Ear and mastoid process | Finland |
| Other specified/unspecified hearing loss | 389 | 389W,389X | H918, H919 | Ear and mastoid process | Sweden |

| | | | | | |
|---|---|---|---|---|---|
| Hypertension, essential | 401,402,403,404 | 401,403 | I10 | Circulatory system | Finland, Sweden |
| Hypertensive Renal Disease | 40399 | 4039 | I12 | Circulatory system | Finland |
| Angina pectoris | 413 | 413,4110,4111 | I20 | Circulatory system | Finland |
| Myocardial infarction, strict | 410 | 410 | I21, I22 | Circulatory system | Finland, Sweden |
| Pulmonary embolism | 450 | 415 | I26 | Circulatory system | Finland, Sweden |
| Pericarditis | 420 | 420 | I30, I32 | Circulatory system | Finland, Sweden |
| Non-rheumatic valve diseases | 394, 395, 396, 4240, 4241 | 424 | I34, I35, I36, I37 | Circulatory system | Finland, Sweden |
| Myocarditis | 422, 742 | 422 | I40, I41 | Circulatory system | Finland, Sweden |
| Paroxysmal tachycardia | 4275 | 4270, 4271, 4272 | I47 | Circulatory system | Finland |
| Paroxysmal tachycardia | 4275 | 427A, 427B, 427C | I47 | Circulatory system | Sweden |
| Atrial fibrillation and flutter | 42792 | 4273 | I48 | Circulatory system | Finland |
| Atrial fibrillation and flutter | 42792 | 427D | I48 | Circulatory system | Sweden |
| Other arrhythmias | 42790, 42791, 42798, 42799 | 4274, 4275, 4276, 4278 | I49 | Circulatory system | Finland |
| Other arrhythmias | 42790, 42791, 42798, 42799 | 427E, 427F, 427G, 427W | I49 | Circulatory system | Sweden |
| Heart failure, strict | 42700, 42710, 428, 7824 | 4029B, 404, 4148, 428 | I50, I110, I130, I132 | Circulatory system | Finland |
| Heart failure, strict | 42700, 42710, 428, 7824 | 402X, 404, 414W, 428 | I50, I110, I130, I132 | Circulatory system | Sweden |
| Other or ill-defined heart diseases | 42899, 42999 | 429 | I51, I52 | Circulatory system | Finland, Sweden |
| Subarachnoid haemorrhage | 430 | 430 | I60 | Circulatory system | Finland, Sweden |
| Intracerebral haemorrhage | 431 | 431 | I61 | Circulatory system | Sweden |
| Ischaemic Stroke, excluding all haemorrhages | 433, 434, 436 | 4330A, 4331A, 4339A, 4340, 4341, 4349A, 436 | I630, I631, I632, I633, I634, I635, I638, I639, I64 | Circulatory system | Finland |
| Ischaemic Stroke, excluding all haemorrhages | 433, 434, 436 | 433A, 433B, 433X, 434A, 434B, 434X, 436 | I630, I631, I632, I633, I634, I635, I638, I639, I64 | Circulatory system | Sweden |
| Aortic aneurysm | 441, 930 | 4411, 4412, 4413, 4414, 4415, 4419, 0930 | I711, I712, I713, I714, I715, I716, I718, I719 | Circulatory system | Finland |
| DVT of lower extremities | 4510 | 4511A, 4512A | I802, I803 | Circulatory system | Finland |
| DVT of lower extremities | 4510 | 451B, 451C | I802, I803 | Circulatory system | Sweden |
| Other embolism and thrombosis | 453 | 453 | I82 | Circulatory system | Finland, Sweden |
| Varicose veins | 454 | 454 | I83 | Circulatory system | Finland, Sweden |
| Varicose veins of other sites | 45610, 45698 | 4563, 4564, 4565, 4566, 4568 | I86 | Circulatory system | Finland |
| Varicose veins of other sites | 45610, 45698 | 456D, 456E, 456F, 456G, 456W | I86 | Circulatory system | Sweden |
| Nonspecific lymphadenitis | 2891, 2892, 2893 | 2891, 2892 | I88 | Circulatory system | Finland |
| Nonspecific lymphadenitis | 2891, 2892, 2893 | 289B, 289C | I88 | Circulatory system | Sweden |

| | | | | | |
|---|---|---|---|---|---|
| Acute nasopharyngitis (common cold) | 460 | 460 | J00 | Respiratory system | Finland, Sweden |
| Acute sinusitis | 461 | 461 | J01 | Respiratory system | Finland, Sweden |
| Acute pharyngitis | 462 | 462 | J02 | Respiratory system | Finland, Sweden |
| Other and unspecified tonsillitis | 463 | 463 | J03 | Respiratory system | Finland, Sweden |
| Acute laryngitis and tracheitis | 46401, 46403, 46409, 50803 | 4640, 4641, 4642, 4644 | J04, J050 | Respiratory system | Finland |
| Acute laryngitis and tracheitis | 46401, 46402, 46409, 50800 | 464A, 464B, 464C, 464E | J04, J050 | Respiratory system | Sweden |
| Acute epiglottitis | 46403 | 464D | J051 | Respiratory system | Sweden |
| Acute upper respiratory infections of multiple and unspecified sites | 465 | 465 | J06 | Respiratory system | Finland, Sweden |
| All influenza | 470, 472, 473, 474 | 487 | J101, J108, J111, J118 | Respiratory system | Finland, Sweden |
| Viral pneumonia (unknown virus, not influenza) | 480 | 480 | J128, J129 | Respiratory system | Sweden |
| Pneumonia due to Streptococcus pneumoniae | 48199 | 481 | J13 | Respiratory system | Finland, Sweden |
| Bacterial pneumonia (organism specified) | 48199, 482 | 481, 482, 483 | J13, J14, J15, J16, J17 | Respiratory system | Finland |
| Pneumonia due to other infectious organisms, not elsewhere classified | 48399 | 483 | J16, J172, J173, J178 | Respiratory system | Finland, Sweden |
| Pneumonia, organism unspecified | 48502, 48509, 486 | 485 | J18 | Respiratory system | Finland, Sweden |
| Acute bronchitis | -- | 4660 | J20 | Respiratory system | Finland |
| Acute bronchitis | -- | 466A | J20 | Respiratory system | Sweden |
| Unspecified acute lower respiratory infection | 46699 | -- | J22 | Respiratory system | Finland, Sweden |
| Vasomotor and allergic rhinitis | 507 | 477 | J30 | Respiratory system | Finland, Sweden |
| Chronic rhinitis, nasopharyngitis and pharyngitis | 502 | 472 | J31 | Respiratory system | Finland |
| Chronic sinusitis | 503 | 473 | J32 | Respiratory system | Finland, Sweden |
| Nasal polyp | 505 | 471 | J33 | Respiratory system | Finland, Sweden |
| Diseases of vocal cords and larynx, other diseases of upper respiratory tract, not elsewhere classified | 50800, 50801, 50802, 50804, 50805, 50806, 50808, 50809 | 478 | J34, J38, J39 | Respiratory system | Finland, Sweden |
| Deviated nasal septum | 50499 | 470 | J342 | Respiratory system | Finland, Sweden |
| Chronic diseases of tonsils and adenoids | 500 | 474 | J35 | Respiratory system | Finland, Sweden |
| Peritonsillar abscess | 501 | 475 | J36 | Respiratory system | Finland, Sweden |
| Bronchitis, not specified as acute or chronic | 49099 | 490 | J40 | Respiratory system | Finland, Sweden |
| Unspecified chronic bronchitis | 49109 | 4919 | J42 | Respiratory system | Finland |

| | | | | | |
|---|---|---|---|---|---|
| Non-allergic asthma (mode) | -- | -- | J451 | Respiratory system | Finland |
| Bronchiectasis | 51899 | 494 | J47 | Respiratory system | Finland |
| Other interstitial pulmonary diseases | 48499, 51701 | 515, 516 | J84 | Respiratory system | Finland |
| Pneumothorax | 51299 | 512 | J93 | Respiratory system | Finland, Sweden |
| Other pleural conditions | 51101, 51109 | 511A, 511W, 511X | J94 | Respiratory system | Sweden |
| Other respiratory disorders and diseases | 51900, 51901, 51921, 51922, 51925, 51928, 51991, 51992, 51993, 51994, 51998 | 5180, 5181, 5188, 519 | J95, J96, J98 | Respiratory system | Finland |
| Other respiratory disorders and diseases | 51900, 51901, 51921, 51922, 51925, 51928, 51991, 51992, 51993, 51994, 51998 | 518A,518B,518W,519 | J95, J96, J98 | Respiratory system | Sweden |
| Oesophagitis | 5301 | 530B | K20 | Digestive system | Sweden |
| Gastro-oesophageal reflux disease | -- | 5301A | K21 | Digestive system | Finland |
| Gastro-oesophageal reflux disease | -- | 530B | K21 | Digestive system | Sweden |
| Other diseases of oesophagus | 5302, 5309 | 5305, 5306, 5307, 5308, 5309 | K224, K225, K226, K228, K229 | Digestive system | Finland |
| Other diseases of oesophagus | 5302, 5309 | 530F, 530G, 530H, 530W, 530X | K224, K225, K226, K228, K229 | Digestive system | Sweden |
| Barret oesophagus | -- | 530B | K227 | Digestive system | Sweden |
| Duodenal ulcer | -- | 532 | K26 | Digestive system | Finland |
| Acute gastritis | 53500, 53501, 53502 | 5350 | K290, K291 | Digestive system | Finland |
| Acute gastritis | 53500, 53501, 53502 | 535A | K290, K291 | Digestive system | Sweden |
| Alcoholic gastritis | -- | 5353 | K292 | Digestive system | Finland |
| Chronic gastritis | 53503 | 5351, 5354 | K293, K294, K295 | Digestive system | Finland |
| Other gastritis (incl. Duodenitis) | 53504, 53505, 53506, 53507, 53508, 53509 | 5352, 5355, 5356 | K296, K297, K298, K299 | Digestive system | Finland |
| Other gastritis (incl. Duodenitis) | 53504, 53505, 53506, 53507, 53508, 53509 | 535C, 535F, 535G | K296, K297, K298, K299 | Digestive system | Sweden |
| Functional dyspepsia | 5361 | 5368 | K30 | Digestive system | Finland |
| Functional dyspepsia | 5361 | 536W | K30 | Digestive system | Sweden |
| Other diseases of stomach and duodenum | 5360, 5369, 537 | 5361, 5362, 5368X, 5369, 537 | K31 | Digestive system | Finland |
| Acute appendicitis | 540 | 540 | K35 | Digestive system | Finland, Sweden |
| Other appendicitis | 541, 542 | 541, 542 | K36, K37 | Digestive system | Finland, Sweden |
| Other diseases of appendix | 543 | 543 | K38 | Digestive system | Finland |
| Inguinal hernia | 550 | 550 | K40 | Digestive system | Finland, Sweden |
| Umbilical hernia | 5511 | 5511, 5521, 5531 | K42 | Digestive system | Finland |
| Umbilical hernia | 5511 | 551B, 552B, 553B | K42 | Digestive system | Sweden |
| Ventral hernia | 5512 | 5512, 5522, 5532 | K43 | Digestive system | Finland |
| Ventral hernia | 5512 | 551C, 552C, 553C | K43 | Digestive system | Sweden |
| Diaphragmatic hernia | 5513 | 551D, 552D, 553D | K44 | Digestive system | Sweden |
| Ulcerative colitis | 5631,569 | 5560A, 5561A, 5562A, 5563A, 5568X | K51 | Digestive system | Finland |
| Ulcerative colitis | 5631,569 | 556 | K51 | Digestive system | Sweden |
| Other noninfective gastroenteritis and colitis | 562,5639 | 558, 5564 | K52 | Digestive system | Finland |

| | | | | | |
|---|---|---|---|---|---|
| Other noninfective gastroenteritis and colitis | 562,5639 | 558,556 | K52 | Digestive system | Sweden |
| Paralytic ileus | 5601 | 560B | K560 | Digestive system | Sweden |
| Other or unspecified ileus, impaction, or obstruction | 5609 | 5608X, 5609X | K564, K566, K567 | Digestive system | Finland |
| Other or unspecified ileus, impaction, or obstruction | 5609 | 560W, 560X | K564, K566, K567 | Digestive system | Sweden |
| Intestinal adhesions without obstruction | 56095, 56096 | 5608A | K565 | Digestive system | Finland |
| Intestinal adhesions without obstruction | 56095, 56096 | 560W | K565 | Digestive system | Sweden |
| Irritable bowel syndrome | 56419 | 5641 | K58 | Digestive system | Finland |
| Irritable bowel syndrome | 56419 | 564B | K58 | Digestive system | Sweden |
| Other functional intestinal disorders | 56400, 56410, 56411, 56490, 56498, 56499 | 5640, 5642, 5645, 5646, 5647, 5648, 5649 | K59 | Digestive system | Finland |
| Other functional intestinal disorders | 56400, 56410, 56411, 56490, 56498, 56499 | 564A, 564C, 564F, 564G, 564H, 564W, 564X | K59 | Digestive system | Sweden |
| Fissure and fistula of anal and rectal regions | 565 | 565 | K60 | Digestive system | Finland, Sweden |
| Abscess of anal and rectal regions | 566 | 566 | K61 | Digestive system | Finland, Sweden |
| Other diseases of anus and rectum | 56906, 56907 | 5690, 5691, 5692, 5694 | K62 | Digestive system | Finland |
| Other diseases of anus and rectum | 56906, 56907 | 569A, 569B, 569C, 569E | K62 | Digestive system | Sweden |
| Other diseases of intestine | 56900, 56901, 56902, 56903, 56904, 56905, 56908, 56909 | 5695, 5696, 5698, 5699 | K63 | Digestive system | Finland |
| Other diseases of intestine | 56900, 56901, 56902, 56903, 56904, 56905, 56908, 56909 | 569F, 569G, 569W, 569X | K63 | Digestive system | Sweden |
| Haemorrhoids and perianal venous thrombosis | 455 | 455 | K64 | Digestive system | Finland, Sweden |
| Acute peritonitis | 567 | 567 | K65 | Digestive system | Finland, Sweden |
| Other diseases of peritoneum | 56900, 56901, 56903, 56904, 56905, 56906, 56907, 56908, 56909 | 568 | K66 | Digestive system | Finland, Sweden |
| Alcoholic liver disease | 5710 | 5710, 5711, 5712, 5713 | K70 | Digestive system | Finland |
| Other inflammatory liver diseases | 57300 | 5720 | K75 | Digestive system | Finland |
| Other diseases of liver | 57301, 57302, 57303, 57308, 57309 | 5718, 5719, 5721, 5722, 5723, 5724, 5728 | K76 | Digestive system | Finland |
| Other diseases of liver | 57301, 57302, 57303, 57308, 57309 | 571W, 571X, 572B, 572C, 572D, 572E, 572W | K76 | Digestive system | Sweden |
| Cholelithiasis | 574 | 574 | K80 | Digestive system | Finland, Sweden |
| Cholecystitis | 57500, 57501, 57502, 57503 | 5750, 5751 | K81 | Digestive system | Finland |
| Cholecystitis | 57500, 57501, 57502, 57503 | 575A, 575B | K81 | Digestive system | Sweden |

| | | | | | |
|---|---|---|---|---|---|
| Acute pancreatitis | 5770 | 5770 | K85 | Digestive system | Finland |
| Alcohol-induced acute pancreatitis | -- | 5770D, 5770E | K852 | Digestive system | Finland |
| Alcohol-induced chronic pancreatitis | -- | 5771C,5771D | K860 | Digestive system | Finland |
| Coeliac disease | -- | 5790 | K900 | Digestive system | Finland |
| Intestinal malabsorption | 2691 | 5791, 5792, 5794, 5798 | K901, K902, K903, K904, K908, K909 | Digestive system | Finland |
| Intestinal malabsorption | 2691 | 579B, 579C, 579E, 579W | K901, K902, K903, K904, K908, K909 | Digestive system | Sweden |
| Impetigo | 684,70708 | 684 | L01 | Skin and subcutaneous tissue | Finland |
| Cutaneous abscess, furuncle, and carbuncle | 680 | 680 | L02 | Skin and subcutaneous tissue | Finland, Sweden |
| Cellulitis | 681,682 | 681,682 | L03 | Skin and subcutaneous tissue | Finland, Sweden |
| Acute lymphadenitis | 683 | 683 | L04 | Skin and subcutaneous tissue | Finland, Sweden |
| Pilonidal cyst | -- | 685 | L05 | Skin and subcutaneous tissue | Finland, Sweden |
| Pyoderma | 68600 | 6860 | L080 | Skin and subcutaneous tissue | Finland |
| Other and unspecified local infections of skin and subcutaneous tissue | 68690, 68691, 68698 | 6868, 6869 | L088, L089 | Skin and subcutaneous tissue | Finland |
| Atopic dermatitis | 691 | 6918A, 6918B, 6918C | L20 | Skin and subcutaneous tissue | Finland |
| Atopic dermatitis | 691 | 691W | L20 | Skin and subcutaneous tissue | Sweden |
| Seborrhoeic dermatitis | 690, 70630 | 690 | L21 | Skin and subcutaneous tissue | Finland |
| Allergic contact dermatitis | -- | 6920B, 6921B, 6922B, 6924B, 6924D, 6924H, 6924L, 6925B, 6925D, 6926B, 6926D | L23 | Skin and subcutaneous tissue | Finland |
| Other contact dermatitis | 6921, 6922, 6923, 6924, 6926, 69281 | 6920A, 6921A, 6922A, 6923, 6924A, 6924C, 6924E, 6924F, 6924G, 6924K, 6924M, 6924N, 6925A, 6925C, 6925X, 6926A, 6926C, 6926X, 6927, 6928, 6929 | L24, L25 | Skin and subcutaneous tissue | Finland |
| Pruritus | 6980, 6981, 6989 | 698A, 698B, 698W, 698X | L29 | Skin and subcutaneous tissue | Sweden |
| Other and unspecified dermatitis | 69282, 69288, 6929 | -- | L308, L309 | Skin and subcutaneous tissue | Finland, Sweden |
| Psoriasis vulgaris | -- | 6961A | L400 | Skin and subcutaneous tissue | Finland |
| Psoriasis vulgaris | -- | 696B | L400 | Skin and subcutaneous tissue | Sweden |
| Other and unspecified psoriasis | 69619 | 6968X | L408, L409 | Skin and subcutaneous tissue | Finland |
| Other and unspecified psoriasis | 69619 | 696W | L408, L409 | Skin and subcutaneous tissue | Sweden |

| | | | | | |
|---|---|---|---|---|---|
| Allergic urticaria | -- | 7080 | L500 | Skin and subcutaneous tissue | Finland |
| Allergic urticaria | -- | 708A | L500 | Skin and subcutaneous tissue | Sweden |
| Other and unspecified urticaria | 708 | 7088, 7089 | L508, L509 | Skin and subcutaneous tissue | Finland |
| Other and unspecified urticaria | 708 | 708W, 708X | L508, L509 | Skin and subcutaneous tissue | Sweden |
| Erythema nodosum | 69520 | 6952 | L52 | Skin and subcutaneous tissue | Finland |
| Other and unspecified acute skin changes due to ultraviolet radiation | 6928 | 6927B, 6927C, 6927D, 6927E, 6927H, 6927X | L568, L569 | Skin and subcutaneous tissue | Finland |
| Ingrowing nail | 70302 | 7030 | L600 | Skin and subcutaneous tissue | Finland |
| Acne vulgaris | 70610 | 7061A | L700 | Skin and subcutaneous tissue | Finland |
| Acne vulgaris | 70610 | 706B | L700 | Skin and subcutaneous tissue | Sweden |
| Rosacea | -- | -- | L71 | Skin and subcutaneous tissue | Sweden |
| Follicular cysts of skin and subcutaneous tissue | 70620 | 7062A | L72 | Skin and subcutaneous tissue | Finland |
| Follicular cysts of skin and subcutaneous tissue | 70620 | 706C | L72 | Skin and subcutaneous tissue | Sweden |
| Corns and callosities | 700 | 700 | L84 | Skin and subcutaneous tissue | Finland |
| Scar conditions and fibrosis of skin | 70906,70907 | 709C | L905 | Skin and subcutaneous tissue | Sweden |
| Hypertrophic scar | 70130 | 7014B | L910 | Skin and subcutaneous tissue | Finland |
| Hypertrophic scar | 70130 | 701E | L910 | Skin and subcutaneous tissue | Sweden |
| Other and unspecified disorders of skin and subcutaneous tissue | 70198, 70908, 70909 | 7019X, 7098C, 7098X, 7099 | L988, L989 | Skin and subcutaneous tissue | Finland |
| Pyogenic arthritis | 710 | 7110, 7119 | M00, M01 | Musculoskeletal system | Finland |
| Pyogenic arthritis | 710 | 711A, 711X | M00, M01 | Musculoskeletal system | Sweden |
| Reactive arthropathies | 71490 | 7111, 7115, 7118, 713 | M02, M03 | Musculoskeletal system | Finland |
| Reactive arthropathies | 71490 | 711B, 711F, 711W, 713 | M02, M03 | Musculoskeletal system | Sweden |
| Polyarthropathies | -- | -- | M05, M06, M07, M08, M09, M10, M11, M12, M13, M14 | Musculoskeletal system | Finland, Sweden |
| Other/unspecified seropositive rheumatoid arthritis | -- | 7140A | M058, M059 | Musculoskeletal system | Finland |
| Seronegative rheumatoid arthritis | -- | 7140B | M060 | Musculoskeletal system | Finland |
| Juvenile idiopathic arthritis | 7120 | 7143A | M080 | Musculoskeletal system | Finland |
| Juvenile idiopathic arthritis | 7120 | 714D | M080 | Musculoskeletal system | Sweden |
| Other juvenile arthritis | -- | 7143B, 7143C, 7143D, 7143X | M081, M082, M083, M084, M088, M089, M09 | Musculoskeletal system | Finland |

| | | | | | |
|---|---|---|---|---|---|
| Monoarthritis, not elsewhere classified | 71100, 71101, 71102, 71103, 71104, 71105, 71106 | 7166 | M131 | Musculoskeletal system | Finland |
| Monoarthritis, not elsewhere classified | 71100, 71101, 71102, 71103, 71104, 71105, 71106 | 716G | M131 | Musculoskeletal system | Sweden |
| Other specific/unspecified arthritis | 71108, 71109, 71498 | 7169 | M138, M139 | Musculoskeletal system | Finland |
| Coxarthrosis | 71300 | 715B,715C | M16 | Musculoskeletal system | Sweden |
| Gonarthrosis | 71301 | 7151F,7152F | M17 | Musculoskeletal system | Finland |
| Gonarthrosis | 71301 | 715B,715C | M17 | Musculoskeletal system | Sweden |
| Other arthrosis | 71302, 71308, 71309, 71320 | 7151A, 7151B, 7151C, 7151D, 7151G, 7151H, 7151K, 7151X, 7152A, 7152B, 7152C, 7152D, 7152G, 7152H, 7152K, 7152X | M18, M19 | Musculoskeletal system | Finland |
| Other arthrosis | 71302, 71308, 71309, 71320 | 715B, 715C | M18, M19 | Musculoskeletal system | Sweden |
| Other joint disorders | -- | -- | M20, M21, M22, M23, M24, M25 | Musculoskeletal system | Finland, Sweden |
| Hallux valgus (acquired) | 737, 73099 | 7350, 7271 | M201 | Musculoskeletal system | Finland |
| Hallux valgus (acquired) | 737, 73099 | 735A, 727B | M201 | Musculoskeletal system | Sweden |
| Flat foot [pes planus] (acquired) | 736 | 734 | M214 | Musculoskeletal system | Finland |
| Other specified acquired deformities of limbs | 73808, 73809 | 7368,7369 | M218, M219 | Musculoskeletal system | Finland |
| Other disorders of patella | -- | 7177 | M222, M223, M224, M228, M229 | Musculoskeletal system | Finland |
| Other disorders of patella | -- | 717H | M222, M223, M224, M228, M229 | Musculoskeletal system | Sweden |
| Meniscus derangement | -- | 7170, 7174, 7175 | M230, M231, M232, M233 | Musculoskeletal system | Finland |
| Meniscus derangement | -- | 717A, 717E,7 17F | M230, M231, M232, M233 | Musculoskeletal system | Sweden |
| Internal derangement of knee | 72410 | 7176, 7178, 7179 | M234, M235, M236, M238, M239 | Musculoskeletal system | Finland |
| Internal derangement of knee | 72410 | 717G, 717W, 717X | M234, M235, M236, M238, M239 | Musculoskeletal system | Sweden |
| Loose body in joint | 72901 | 7181 | M240 | Musculoskeletal system | Finland |
| Loose body in joint | 72901 | 718B | M240 | Musculoskeletal system | Sweden |
| Other articular cartilage disorders | 7249 | 7180 | M241 | Musculoskeletal system | Finland |
| Other articular cartilage disorders | 7249 | 718A | M241 | Musculoskeletal system | Sweden |
| Other specific joint derangements/joint disorders | 72902, 72903 | 7188, 7189 | M242, M248, M249, M25 | Musculoskeletal system | Finland |
| Other specific joint derangements/joint disorders | 72902, 72903 | 718W, 718X | M242, M248, M249, M25 | Musculoskeletal system | Sweden |
| Pathological/recurrent dislocation and subluxation of joint, not elsewhere classified | 72904 | 7182 | M243 | Musculoskeletal system | Finland |

| | | | | | |
|---|---|---|---|---|---|
| Pathological/recurrent dislocation and subluxation of joint, not elsewhere classified | 72904 | 718C | M243 | Musculoskeletal system | Sweden |
| Recurrent dislocation and subluxation of joint | -- | 7183 | M244 | Musculoskeletal system | Finland |
| Recurrent dislocation and subluxation of joint | -- | 718D | M244 | Musculoskeletal system | Sweden |
| Systemic lupus erythematosus | 7431 | 7100 | M32 | Musculoskeletal system | Finland |
| Systemic lupus erythematosus | 7431 | 710A | M32 | Musculoskeletal system | Sweden |
| Other specified/unspecified systemic involvement of connective tissue | 73491, 73498, 73499 | 710W,710X | M358, M359 | Musculoskeletal system | Sweden |
| Torticollis | 7172 | 7235 | M436 | Musculoskeletal system | Finland |
| Other specified/unspecified deforming dorsopathies | 735 | 7328, 7329, 7378, 7379 | M438, M439 | Musculoskeletal system | Finland |
| Other specified/unspecified deforming dorsopathies | 735 | 732W, 732X, 737W, 737X | M438, M439 | Musculoskeletal system | Sweden |
| Ankylosing spondylitis | 7124 | 7200 | M45 | Musculoskeletal system | Finland |
| Ankylosing spondylitis | 7124 | 720A | M45 | Musculoskeletal system | Sweden |
| Spondyloarthritis | 7124, 7131, 72699 | 720 | M45, M46 | Musculoskeletal system | Finland |
| Spondylopathies | -- | -- | M45, M46, M47, M48, M49 | Musculoskeletal system | Finland, Sweden |
| Cervical disc disorders | 7250 | 7220, 7224, 7227, 7228A | M50 | Musculoskeletal system | Finland |
| Other intervertebral disc disorders | 72510, 72519, 72588, 72599 | 7221, 7223, 7225, 7226, 7229 | M51 | Musculoskeletal system | Finland |
| Other intervertebral disc disorders | 72510, 72519, 72588, 72599 | 722B, 722D, 722F, 722G, 722X | M51 | Musculoskeletal system | Sweden |
| Radiculopathy | 7283, 7288 | 7234, 7244 | M541 | Musculoskeletal system | Finland |
| Radiculopathy | 7283, 7288 | 723E, 724E | M541 | Musculoskeletal system | Sweden |
| Cervicalgia | 7289 | 7231 | M542 | Musculoskeletal system | Finland |
| Cervicalgia | 7289 | 723B | M542 | Musculoskeletal system | Sweden |
| Sciatica, lumbago | 7170 | -- | M543, M544 | Musculoskeletal system | Finland, Sweden |
| Low back pain | 72870 | 7242 | M545 | Musculoskeletal system | Finland |
| Low back pain | 72870 | 724C | M545 | Musculoskeletal system | Sweden |
| Pain in thoracic spine | 7285 | 7241 | M546 | Musculoskeletal system | Finland |
| Other/unspecified dorsalgia | 7289 | 7245 | M548, M549 | Musculoskeletal system | Finland |
| Other/unspecified dorsalgia | 7289 | 724F | M548, M549 | Musculoskeletal system | Sweden |
| Other specified disorders of muscle | 73390 | 7288,7289 | M628, M629 | Musculoskeletal system | Finland |
| Disorders of synovium and tendon | -- | -- | M65, M66, M67, M68 | Musculoskeletal system | Finland, Sweden |
| Radial styloid tenosynovitis [de Quervain] | -- | 727A | M654 | Musculoskeletal system | Sweden |

| | | | | | |
|---|---|---|---|---|---|
| Other/unspecified synovitis and tenosynovitis | 73100, 73101, 73102, 73103 | 7270B, 7270C, 7270X | M658, M659 | Musculoskeletal system | Finland |
| Other/unspecified synovitis and tenosynovitis | 73100, 73101, 73102, 73103 | 727A | M658, M659 | Musculoskeletal system | Sweden |
| Spontaneous rupture of synovium and tendon | -- | 7276 | M66 | Musculoskeletal system | Finland |
| Ganglion | -- | 7274 | M674 | Musculoskeletal system | Finland |
| Ganglion | -- | 727E | M674 | Musculoskeletal system | Sweden |
| Prepatellar bursitis | -- | 7266E | M704 | Musculoskeletal system | Finland |
| Other bursal cyst | -- | 7274 | M713 | Musculoskeletal system | Finland |
| Other bursitis, not elsewhere classified | 73105, 73109 | 7273 | M715 | Musculoskeletal system | Finland |
| Other bursitis, not elsewhere classified | 73105, 73109 | 727D | M715 | Musculoskeletal system | Sweden |
| Rotator cuff syndrome | -- | 7261X | M751 | Musculoskeletal system | Finland |
| Achilles tendinitis | -- | 7267A | M766 | Musculoskeletal system | Finland |
| Other/unspecified enthesopathies, not elsewhere classified | 73105, 73109 | 7268, 7269 | M778, M779 | Musculoskeletal system | Finland |
| Other/unspecified enthesopathies, not elsewhere classified | 73105, 73109 | 726W, 726X | M778, M779 | Musculoskeletal system | Sweden |
| Rheumatism, unspecified | 71899 | 7290 | M790 | Musculoskeletal system | Finland |
| Myalgia | -- | 729B | M791 | Musculoskeletal system | Sweden |
| Pain in limb | -- | -- | M796 | Musculoskeletal system | Finland, Sweden |
| Other specified/unspecified soft tissue disorders | 71798 | 729W, 729X | M798, M799 | Musculoskeletal system | Sweden |
| Osteomyelitis | 720 | 7300, 7301, 7302, 7303, 7309 | M86 | Musculoskeletal system | Finland |
| Osteomyelitis | 720 | 730A, 730B, 730C, 730D, 730X | M86 | Musculoskeletal system | Sweden |
| Other specified/unspecified disorders of bone/cartilage | 72398, 72399 | 7339 | M898, M899, M948, M949 | Musculoskeletal system | Finland |
| Other specified/unspecified disorders of bone/cartilage | 72398, 72399 | 733X | M898, M899, M948, M949 | Musculoskeletal system | Sweden |
| Juvenile osteochondrosis | 72209, 72228, 72229 | 7321X, 7323, 7324, 7325, 7326 | M91, M92 | Musculoskeletal system | Finland |
| Other osteochondropathies | 72200, 72210, 72211, 72219, 72220, 72221, 72222, 72223, 72288, 72299 | 7322, 7327, 7328, 7329 | M93 | Musculoskeletal system | Finland |
| Other osteochondropathies | 72200, 72210, 72211, 72219, 72220, 72221, 72222, 72223, 72288, 72299 | 732C, 732H, 732W, 732X | M93 | Musculoskeletal system | Sweden |
| Other acquired deformities of musculoskeletal system and connective tissue | 73808, 73809 | 738 | M95 | Musculoskeletal system | Finland, Sweden |
| Acute nephritic syndrome | 580 | 5800, 5808, 5809 | N00 | Genitourinary system | Finland |

| | | | | | |
|---|---|---|---|---|---|
| Acute nephritic syndrome | 580 | 580A, 580W, 580X | N00 | Genitourinary system | Sweden |
| Chronic nephritic syndrome | 582 | 582 | N03 | Genitourinary system | Finland, Sweden |
| Unspecified nephritic syndrome | 583 | 583 | N05 | Genitourinary system | Finland |
| Acute tubulo-interstitial nephritis | 5901 | 5901 | N10 | Genitourinary system | Finland |
| Acute tubulo-interstitial nephritis | 5901 | 590B | N10 | Genitourinary system | Sweden |
| Chronic tubulo-interstitial nephritis | 5900 | 5900 | N11 | Genitourinary system | Finland |
| Chronic tubulo-interstitial nephritis | 5900 | 590A | N11 | Genitourinary system | Sweden |
| Tubulo-interstitial nephritis, not specified as acute or chronic | 5909 | 5909 | N12 | Genitourinary system | Finland |
| Hydronephrosis | 591 | 591 | N130, N131, N132, N133 | Genitourinary system | Finland, Sweden |
| Other obstructive and reflux uropathy | 5933, 5934, 5935 | 5933, 5934, 5935, 5937 | N134, N135, N136, N137, N138, N139 | Genitourinary system | Finland |
| Other obstructive and reflux uropathy | 5933, 5934, 5935 | 593D, 593E, 593F, 593H | N134, N135, N136, N137, N138, N139 | Genitourinary system | Sweden |
| Other renal tubulo-interstitial diseases | 5902, 5909 | 5902,5909 | N15 | Genitourinary system | Finland |
| Calculus of kidney and ureter | 592 | 592 | N20 | Genitourinary system | Finland, Sweden |
| Other specified disorders of kidney and ureter | 59322 | 5930, 5931, 5938A, 5938C, 5938X | N280, N288, N289 | Genitourinary system | Finland |
| Cystitis | 595 | 595 | N30 | Genitourinary system | Finland, Sweden |
| Other disorders of bladder | 596 | 596F, 596W, 596X | N32 | Genitourinary system | Sweden |
| Urethritis and urethral syndrome | 597 | -- | N34 | Genitourinary system | Finland, Sweden |
| Urethral stricture | 598 | -- | N35 | Genitourinary system | Finland, Sweden |
| Other disorders of urethra and urinary system | 599 | 5938B, 5965C, 5965D, 5965E, 5965F, 5965X, 6256 | N36, N39 | Genitourinary system | Finland |
| Other disorders of urethra and urinary system | 599 | 593W, 596F, 599, 625G | N36, N39 | Genitourinary system | Sweden |
| Inflammatory diseases of prostate (prostatitis) | 601 | 601 | N41 | Genitourinary system | Finland, Sweden |
| Hydrocele | 603 | 603 | N430, N431, N432, N433 | Genitourinary system | Finland, Sweden |
| Spermatocele | 6076 | 6081 | N434 | Genitourinary system | Finland |
| Torsion of testis | 6077 | 6082 | N44 | Genitourinary system | Finland |
| Torsion of testis | 6077 | 608C | N44 | Genitourinary system | Sweden |
| Orchitis and epididymitis | 604 | 604 | N45 | Genitourinary system | Finland, Sweden |
| Redundant prepuce, phimosis and paraphimosis | 605 | 605 | N47 | Genitourinary system | Finland, Sweden |
| Other disorders of penis | 6070, 6071, 6072, 6073, 60790 | 6070A, 6071A, 6071B, 6071X, 6072A, 6073A, 6078A, 6078X, 6079X | N480, N481, N482, N483, N485, N486, N488, N489 | Genitourinary system | Finland |

| | | | | | |
|---|---|---|---|---|---|
| Other disorders of penis | 6070, 6071, 6072, 6073, 60790 | 607A, 607B, 607C, 607D, 607W, 607X | N480, N481, N482, N483, N485, N486, N488, N489 | Genitourinary system | Sweden |
| Other disorders of male genital organs | 60780, 60791, 60798 | 6083, 6088 | N50 | Genitourinary system | Finland |
| Other disorders of male genital organs | 60780, 60791, 60798 | 608D, 608W | N50 | Genitourinary system | Sweden |
| Benign mammary dysplasia | 610 | 610 | N60 | Genitourinary system | Finland, Sweden |
| Hypertrophy of breast | 6111 | 6111 | N62 | Genitourinary system | Finland |
| Hypertrophy of breast | 6111 | 611B | N62 | Genitourinary system | Sweden |
| Unspecified lump in breast | -- | 6117B | N63 | Genitourinary system | Finland |
| Other disorders of breast | 6112, 6119 | 6112, 6113, 6114, 6115, 6116, 6117A, 6117C, 6117D, 6117E, 6118, 6119 | N64 | Genitourinary system | Finland |
| Other disorders of breast | 6112, 6119 | 611C, 611D, 611E, 611F, 611G, 611H, 611W, 611X | N64 | Genitourinary system | Sweden |
| Salpingitis and oophoritis | 612, 613, 614 | 6140, 6141, 6142 | N70 | Genitourinary system | Finland |
| Salpingitis and oophoritis | 612, 613, 614 | 614A, 614B, 614C | N70 | Genitourinary system | Sweden |
| Cyst of Bartholin Gland | -- | 6162 | N750 | Genitourinary system | Finland |
| Other and unspecified disease of Bartholin gland | 62212, 62213 | -- | N758, N759 | Genitourinary system | Finland |
| Vaginitis/vulvovaginitis/vulvitis/abscess of vulva | 6221 | 6161 | N760, N761, N762, N763, N764 | Genitourinary system | Finland |
| Vaginitis/vulvovaginitis/vulvitis/abscess of vulva | 6221 | 616B | N760, N761, N762, N763, N764 | Genitourinary system | Sweden |
| Other inflammation of vagina/vulva | 62218, 62219 | 6168, 6169 | N768 | Genitourinary system | Finland |
| Other inflammation of vagina/vulva | 62218, 62219 | 616W, 616X | N768 | Genitourinary system | Sweden |
| Vulvovaginal ulceration/inflammation in other diseases | -- | -- | N77 | Genitourinary system | Finland |
| Torsion of ovary, ovarian pedicle, and fallopian tube | 6150 | 6205 | N835 | Genitourinary system | Finland |
| Torsion of ovary, ovarian pedicle, and fallopian tube | 6150 | 620F | N835 | Genitourinary system | Sweden |
| Polyp of the female genital tract | 62520 | 6210, 6227, 6237, 6246 | N84 | Genitourinary system | Finland |
| Polyp of the female genital tract | 62520 | 621A, 622H, 623H, 624G | N84 | Genitourinary system | Sweden |
| Erosion and ectropion of cervix uteri | 62191 | 6220A | N86 | Genitourinary system | Finland |
| Erosion and ectropion of cervix uteri | 62191 | 622A | N86 | Genitourinary system | Sweden |
| Other noninflammatory disorders of cervix uteri | 62191 | 6222, 6223, 6224, 6225, 6226, 6228, 6229 | N88 | Genitourinary system | Finland |

| | | | | | |
|---|---|---|---|---|---|
| Other noninflammatory disorders of cervix uteri | 62191 | 622C, 622D, 622E, 622F, 622G, 622W, 622X | N88 | Genitourinary system | Sweden |
| Other noninflammatory disorders of vagina | 6200, 6210, 6293, 62950, 6297 | 623 | N89 | Genitourinary system | Finland, Sweden |
| Other noninflammatory disorders of vulva and perineum | 62911, 62920 | 624 | N90 | Genitourinary system | Finland |
| Amenorrhoea | 6260 | 6260 | N910, N911, N912 | Genitourinary system | Finland |
| Amenorrhoea | 6260 | 626A | N910, N911, N912 | Genitourinary system | Sweden |
| Oligomenorrhoea | 6261 | 6261 | N913, N914, N915 | Genitourinary system | Finland |
| Oligomenorrhoea | 6261 | 626B | N913, N914, N915 | Genitourinary system | Sweden |
| Excessive, frequent, and irregular menstruation | 6262, 6264 | 6262, 6263, 6264, 6265, 6266, 6270 | N92 | Genitourinary system | Finland |
| Excessive, frequent, and irregular menstruation | 6262,6264 | 626C, 626D, 626E, 626F, 626G, 627A | N92 | Genitourinary system | Sweden |
| Other abnormal uterine and vaginal bleeding | 6265, 6266, 6267, 6269 | 6267, 6268, 6269 | N93 | Genitourinary system | Finland |
| Other abnormal uterine and vaginal bleeding | 6265, 6266, 6267 ,6269 | 626H, 626W, 626X | N93 | Genitourinary system | Sweden |
| Pain and other conditions associated with female genital organs and menstrual cycle | 6263 | 6250, 6251, 6252, 6253, 6254, 6255, 6258, 6259 | N94 | Genitourinary system | Finland |
| Pain and other conditions associated with female genital organs and menstrual cycle | 6263 | 625A, 625B, 625C, 625D, 625E, 625F, 625W, 625X | N94 | Genitourinary system | Sweden |
| Congenital hydrocephalus | 742 | 7423 | Q03 | Congenital anomalies | Finland |
| Congenital malformations of eye, ear, face, and neck | 744, 745 | 743, 744 | Q10, Q11, Q12, Q13, Q14, Q15, Q16, Q17, Q18 | Congenital anomalies | Finland |
| Congenital anomalies of heart | 746 | 745,746 | Q20, Q21, Q22, Q23, Q24 | Congenital anomalies | Finland |
| Congenital malformations of the circulatory system | 746, 747 | 745, 746, 747 | Q20, Q21, Q22, Q23, Q24, Q25, Q26, Q27, Q28 | Congenital anomalies | Finland |
| Congenital anomalies of digestive system | 749, 750, 751 | 749, 750, 751 | Q35, Q36, Q37, Q38, Q39, Q40, Q41, Q42, Q43, Q44, Q45 | Congenital anomalies | Finland |
| Other congenital malformations of the digestive system | 750, 751 | 750, 751 | Q38, Q39, Q40, Q41, Q42, Q43, Q44, Q45 | Congenital anomalies | Finland |
| Congenital anomalies of genital organs | 752 | 752 | Q50, Q51, Q52, Q53, Q54, Q55, Q56 | Congenital anomalies | Finland |
| Congenital anomalies of urinary system | 753 | 753 | Q60, Q61, Q62, Q63, Q64 | Congenital anomalies | Finland |
| Congenital anomalies of musculoskeletal system | 754, 755, 756 | 754, 755, 756 | Q65, Q66, Q67, Q68, Q69, Q70, Q 71, Q72, Q73, Q74, Q75, Q76, Q77, Q78, Q79 | Congenital anomalies | Finland |

| | | | | | |
|---|---|---|---|---|---|
| Childhood allergy (age < 16) | -- | 6918, 477 | J450, L20, J301, J302, J303, J304, L236, J3019 | Other | Finland |
| Retention of urine | 7861 | 7882 | R33 | Other | Finland |
| Fracture of femur | 820 | 820 | S72 | Other | Finland |
| Toxic effect of ethanol | 9800 | 9800 | T510 | Other | Finland |

#### 5.2. Diseases not included due to virtually complete lack of children

We excluded 11 rare diseases from the analysis for both women and men, since receiving diagnosis of these diseases resulted in an almost complete lack of children in the observational period **(Table S6)**. Specifically, we filtered out diseases that had more than 30 individuals affected from the sibling-based design but less than 5 of affected individuals had live-born children at the end of their reproductive lifespan (up to 45 years old for women, 50 for men).

**Table S6**: Diseases not included in the analysis due to resulting in an almost complete lack of children.

| Disease | ICD-8 | ICD-9 | ICD-10 | Category | Population |
|---|---|---|---|---|---|
| Lymphoid leukaemia | 204 | 204 | C91 | Neoplasms | Finland, Sweden |
| Myeloid leukaemia | 205 | 205 | C92 | Neoplasms | Finland, Sweden |
| Moderate intellectual disability | 312 | 3180 | F71 | Mental-behavioural | Finland |
| Moderate intellectual disability | 312 | 318A | F71 | Mental-behavioural | Sweden |
| Severe intellectual disability | 313 | 3181 | F72 | Mental-behavioural | Finland |
| Severe intellectual disability | 313 | 318B | F72 | Mental-behavioural | Sweden |
| Profound intellectual disability | 314 | 3182 | F73 | Mental-behavioural | Finland |
| Profound intellectual disability | 314 | 318C | F73 | Mental-behavioural | Sweden |
| Other and unspecified intellectual disability | 315 | 3199 | F78, F79 | Mental-behavioural | Finland |
| Other and unspecified intellectual disability | 315 | 319 | F78, F79 | Mental-behavioural | Sweden |
| Muscular dystrophy | 33030,33090 | 3591 | G710 | Nervous system | Finland |
| Muscular dystrophy | 33030,33090 | 359B | G710 | Nervous system | Sweden |
| Hemiplegia | 34400 | 342 | G81 | Nervous system | Finland, Sweden |
| Other and unspecified paralytic syndromes | 34408,34409 | 344 | G838, G839 | Nervous system | Finland, Sweden |
| Disorders of brain, other and unspecified | 34798,34799 | 3488,3489 | G938, G939, G948 | Nervous system | Finland |
| Disorders of brain, other and unspecified | 34798,34799 | 348W,348X | G938, G939, G948 | Nervous system | Sweden |
| Congenital hydrocephalus | 742 | 7423 | Q03 | Congenital anomalies | Finland |

#### 5.3. Disease prevalence from Finland and Sweden

**Figure S3:** Prevalence by age 45 in women (Panel A) and 50 in men (Panel B) for 213 disease diagnoses that were available in both Finland and Sweden. Each dot represents a disease diagnosis. Colors are assigned by disease categories.

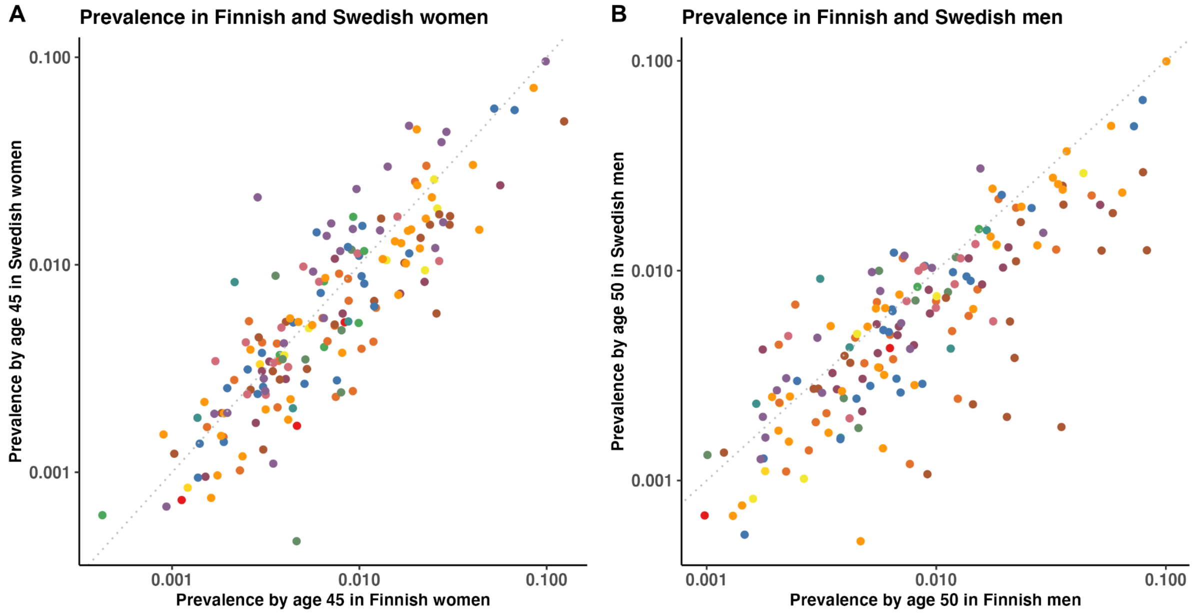

### 6. Relationship of disease diagnoses with childlessness

#### 6.1. Effects of each disease diagnosis

We examined the relationship of 403 disease diagnoses with childlessness by age 45 in women and 50 in men in 71,524 pairs of full-sisters (35,544 in Finland and 35,980 in Sweden) and 77,622 full-brothers (38,928 in Finland and 38,694 in Sweden) who were discordant on childlessness, using a matched case-control design. Only sibling pairs who were also discordant on the disease status were used for the estimation of the disease effect. We therefore reported the number of sibling pairs discordant on both outcome and exposure for each country (**Table S7**). Bonferroni correction was used to control for the familywise error rate.

For each disease diagnosis, we used a fixed-effect inverse-variance weighting method to aggregate summary statistics from two countries and tested for heterogeneity. Overall, the effects were highly consistent between Finnish and Swedish data and, after multiple testing corrections, only two diseases (acute alcohol intoxication and epilepsy) showed significantly heterogeneous effects between the two countries.

There were 74 disease diagnoses that were significantly associated with childlessness ($P<1.5\times10^{-4}$, after multiple-testing correction) in at least one sex, with 33 disease diagnoses shared among women and men. Results can also be explored on the interactive online dashboard available at https://dsgelrs.shinyapps.io/DiseaseSpecificLRS/.

**Table S7**: Relationship of disease diagnoses with childlessness by age 45 in women and 50 in men.

(**Table S7** is too large therefore see sheet "S7_EffectOnChildlessness" from "SUBMIT_TableS7_S9_S11_S12_S17_DiseaseSpecificChildlessness.xlsx"))

#### 6.2. Effects by disease category

To quantify the average disease effects on childlessness for each disease category, we meta-analyzed effects for diseases of the same category for each sex using a random-effect model to account for heterogeneity across different diseases. Among all 16 disease categories, on average, mental-behavioral, congenital anomalies, and endocrine-nutritional-metabolic disorders had the strongest association (**Table S8**).

**Table S8**: Relationship of disease diagnoses with childlessness by age 45 in women and 50 in men by disease category.

| Category | Sex | OR | OR [95% CI lower] | OR [95% CI upper] | P value | P value from heterogeneity test |
|---|---|---|---|---|---|---|
| Infectious-parasitic | Women | 1.07 | 0.97 | 1.18 | 0.20 | 1.62E-03 |
| Infectious-parasitic | Men | 0.97 | 0.90 | 1.05 | 0.51 | 0.43 |
| Neoplasms | Women | 1.16 | 0.77 | 1.75 | 0.47 | 7.14E-09 |
| Neoplasms | Men | 1.21 | 0.79 | 1.83 | 0.38 | 2.03E-07 |
| Blood and immune mechanism | Women | 1.24 | 0.95 | 1.61 | 0.11 | 0.18 |
| Blood and immune mechanism | Men | 1.09 | 0.83 | 1.42 | 0.54 | 0.7 |
| Endocrine-nutritional-metabolic | Women | 1.39 | 1.06 | 1.84 | 0.02 | 1.35E-10 |
| Endocrine-nutritional-metabolic | Men | 2.02 | 1.59 | 2.57 | 7.37E-09 | 0.08 |
| Mental-behavioural | Women | 3.08 | 2.55 | 3.72 | 1.25E-31 | 4.50E-71 |
| Mental-behavioural | Men | 3.22 | 2.61 | 3.97 | 6.79E-28 | 7.86E-128 |
| Nervous system | Women | 1.82 | 1.08 | 3.08 | 0.03 | 1.82E-43 |
| Nervous system | Men | 1.62 | 1.03 | 2.56 | 0.04 | 2.67E-45 |
| Eye and adnexa | Women | 1.31 | 1.12 | 1.52 | 7.01E-04 | 0.01 |
| Eye and adnexa | Men | 1.53 | 1.24 | 1.89 | 6.16E-05 | 7.75E-14 |
| Ear and mastoid process | Women | 1.37 | 1.16 | 1.61 | 1.43E-04 | 0.12 |
| Ear and mastoid process | Men | 1.13 | 0.96 | 1.34 | 0.13 | 0.01 |
| Circulatory system | Women | 1.70 | 1.32 | 2.19 | 3.35E-05 | 1.17E-16 |
| Circulatory system | Men | 1.37 | 1.11 | 1.69 | 3.36E-03 | 3.34E-16 |
| Respiratory system | Women | 1.12 | 1.02 | 1.24 | 0.02 | 2.06E-23 |
| Respiratory system | Men | 0.97 | 0.91 | 1.02 | 0.23 | 1.71E-11 |
| Digestive system | Women | 1.10 | 1.00 | 1.21 | 0.05 | 8.33E-08 |
| Digestive system | Men | 1.26 | 1.13 | 1.40 | 1.91E-05 | 3.53E-19 |
| Skin and subcutaneous tissue | Women | 1.01 | 0.90 | 1.13 | 0.84 | 0.02 |
| Skin and subcutaneous tissue | Men | 1.04 | 0.98 | 1.10 | 0.23 | 0.66 |
| Musculoskeletal system | Women | 1.12 | 1.01 | 1.25 | 0.03 | 1.86E-16 |
| Musculoskeletal system | Men | 0.88 | 0.82 | 0.94 | 2.67E-04 | 2.88E-08 |
| Genitourinary system | Women | 0.97 | 0.88 | 1.07 | 0.52 | 1.58E-11 |
| Genitourinary system | Men | 1.10 | 1.00 | 1.21 | 0.05 | 2.86E-03 |
| Congenital anomalies | Women | 3.55 | 2.57 | 4.89 | 1.15E-14 | 0.01 |
| Congenital anomalies | Men | 1.83 | 1.34 | 2.50 | 1.62E-04 | 0.01 |
| Other | Women | 1.25 | 0.85 | 1.84 | 0.25 | 0.02 |
| Other | Men | 1.10 | 0.91 | 1.33 | 0.32 | 0.21 |

**6.3. Age-of-onset-dependent effects**

We hypothesized that the age when the disease was first diagnosed, a proxy for the age of onset, would influence the chances of being childless by either capturing disease severity or directly impacting factors underlying individuals' reproductive trajectory. Therefore, for disease diagnoses that were significantly associated with childlessness, we assessed whether there were any age-of-onset-dependent effects by considering the age of onset (8 groups <=15, 16-20, 21-25, 26-30, 31-35, 36-40, 41-45, or unaffected for both sexes and an additional group (46-50) for men) as fixed effects in conditional logistic regression. The analysis was performed separately for each disease (**Table S9**) in 71,524 full-sister and 77,622 full-brother pairs who were discordant on childlessness. We then meta-analyzed disease effects for each age of onset group using a random-effect model (**Table S10**).

**Table S9**: Age-of-onset-dependent effects for each disease diagnosis that significantly increased childlessness. (**Table S9** is too large therefore see sheet "S9_AgeOnsetEffects" from "SUBMIT_TableS7_S9_S11_S12_S17_DiseaseSpecificChildlessness.xlsx")

**Table S10**: Age-of-onset-dependent effects across all disease diagnoses that significantly increased childlessness.

| Sex | Age onset group | OR | OR [95% CI lower] | OR [95% CI upper] | P value | P value from heterogeneity test |
|---|---|---|---|---|---|---|
| Women | <=15 | 1.89 | 1.55 | 2.31 | 5.59E-10 | 1.24E-15 |
| Women | 16-20 | 2.53 | 1.98 | 3.25 | 2.05E-13 | 2.78E-21 |
| Women | 21-25 | 3.12 | 2.53 | 3.84 | 1.02E-26 | 1.88E-15 |
| Women | 26-30 | 2.68 | 2.12 | 3.38 | 1.18E-16 | 1.38E-08 |
| Women | 31-35 | 1.74 | 1.33 | 2.29 | 6.35E-05 | 3.08E-04 |
| Women | 36-40 | 1.24 | 0.55 | 2.80 | 0.60 | 0.02 |
| Men | <=15 | 2.27 | 1.86 | 2.77 | 5.89E-16 | 2.18E-05 |
| Men | 16-20 | 2.38 | 1.82 | 3.12 | 2.61E-10 | 4.24E-19 |
| Men | 21-25 | 2.81 | 2.20 | 3.60 | 2.38E-16 | 9.81E-50 |
| Men | 26-30 | 3.07 | 2.45 | 3.85 | 4.07E-22 | 1.34E-09 |
| Men | 31-35 | 2.57 | 1.99 | 3.32 | 5.82E-13 | 2.64E-03 |
| Men | 36-40 | 1.76 | 1.13 | 2.75 | 0.01 | 0.06 |
| Men | 41-45 | 1.03 | 0.49 | 2.17 | 0.93 | 0.35 |

**6.4. Stratification by parity for individuals with children**
We also know that fertility transitions are conditioned on previous births and for that reason a parity-specific analysis is essential[8]. We therefore also look beyond the average fertility level of a country, since the average obscures the distribution of parity and polarization within particular subgroups. This is particularly important when examining childlessness in parity polarised countries[9].

For individuals with children, we assessed whether the effects of diseases were consistent across parities by comparing childless individulas to those with one child, two children, or more than two children. When comparing childless individuals to their siblings with one child, nine out of 28 disease diagnoses examined in women and 15 out of 29 disease diagnoses in men remained significantly associated with childlessness (**Table S11**). Comparing to main analyses (childless individuals versus those with at least one child), 14 disease diagnoses in women and six in men had significantly reduced ORs (**Figure S4A-B**). For example, in women, the OR of schizophrenia on childlessness dropped from 11.6 [9.1-14.9] to 5.1 [3.7-7.0] (P difference= $P=6.9\times10^{-5}$) when restricting to individuals with one child. For individuals with high parities, such as having exactly two children (**Figure 4C-D**) or at least three or more children (**Figure S4E-F**), limited differences in ORs were observed compared to the main analyses. Jointly, these results indicated that, in Nordic countries such as Finland and Sweden, individuals with one child were more impacted by disease status than individuals with high parities, but to a less degree than individuals with no biological children.

**Table S11**. Relationship of disease diagnoses with childlessness by age 45 in women and 50 in men, stratified by parity of individuals with children.

(**Table S11** is too large therefore see sheet “S11_StrataParity” from “SUBMIT_TableS7_S9_S11_S12_S17_DiseaseSpecificChildlessness.xlsx”)

**Figure S4**. Relationship of disease diagnoses with childlessness by age 45 in women (Panel A, C, and E) and 50 in men (Panel B, D, and F) from main analysis (childless individuals were compared to those with at least one child) versus the analysis stratified by parities (childless individuals were compared to those with one child (Panel A and B), with two children (Panel C and D), or with at least three or more children (Panel E and F)), for disease diagnoses that were significantly increased childlessness from the main analysis.

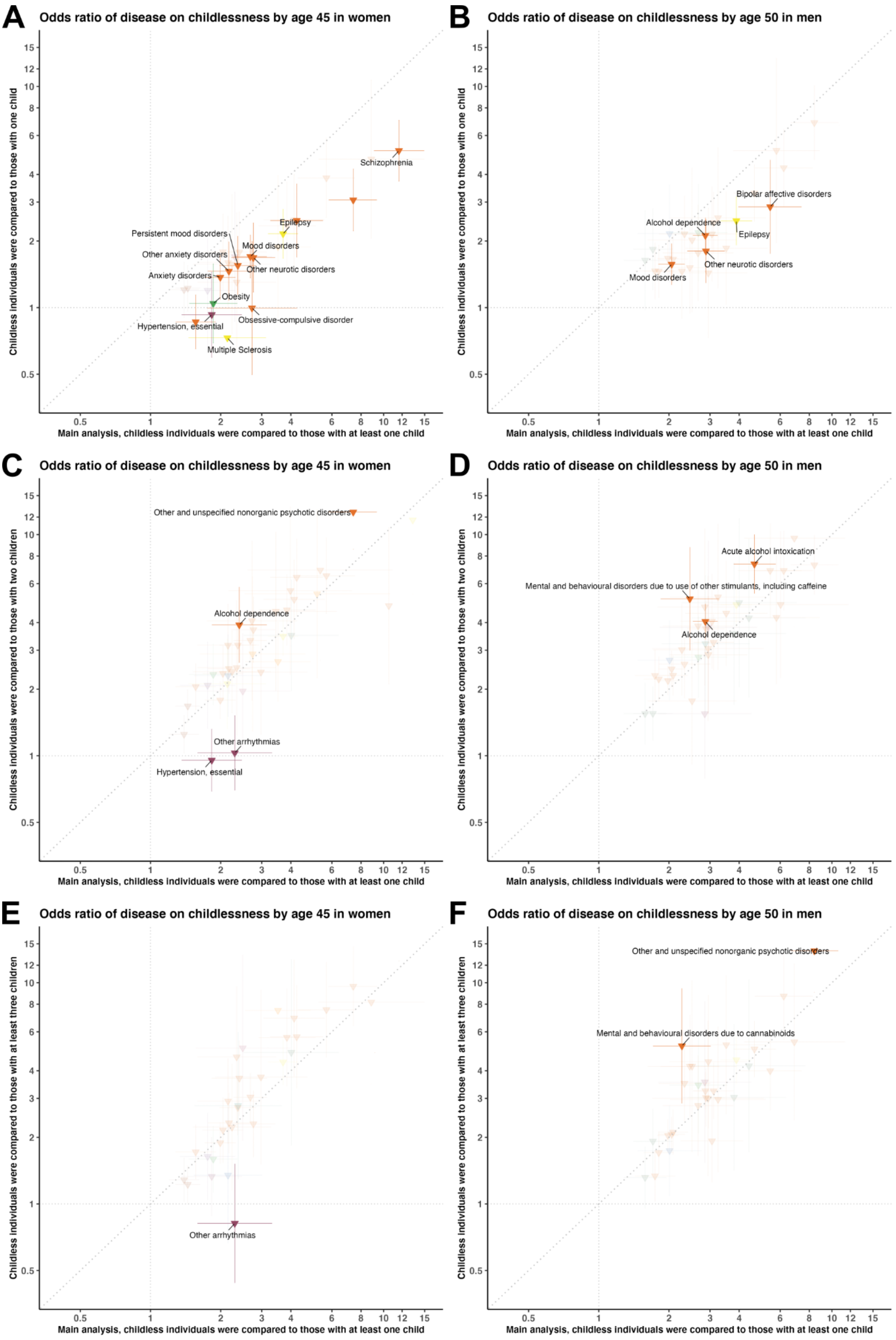
A
Odds ratio of disease on childlessness by age 45 in women
Childless individuals were compared to those with one child
Main analysis, childless individuals were compared to those with at least one child
Schizophrenia
Epilepsy
Persistent mood disorders
Mood disorders
Other anxiety disorders
Other neurotic disorders
Anxiety disorders
Obesity
Obsessive-compulsive disorder
Hypertension, essential
Multiple Sclerosis
B
Odds ratio of disease on childlessness by age 50 in men
Childless individuals were compared to those with one child
Main analysis, childless individuals were compared to those with at least one child
Bipolar affective disorders
Alcohol dependence
Epilepsy
Other neurotic disorders
Mood disorders
C
Odds ratio of disease on childlessness by age 45 in women
Childless individuals were compared to those with two children
Main analysis, childless individuals were compared to those with at least one child
Other and unspecified nonorganic psychotic disorders
Alcohol dependence
Other arrhythmias
Hypertension, essential
D
Odds ratio of disease on childlessness by age 50 in men
Childless individuals were compared to those with two children
Main analysis, childless individuals were compared to those with at least one child
Acute alcohol intoxication
Mental and behavioural disorders due to use of other stimulants, including caffeine
Alcohol dependence
E
Odds ratio of disease on childlessness by age 45 in women
Childless individuals were compared to those with at least three children
Main analysis, childless individuals were compared to those with at least one child
Other arrhythmias
F
Odds ratio of disease on childlessness by age 50 in men
Childless individuals were compared to those with at least three children
Main analysis, childless individuals were compared to those with at least one child
Other and unspecified nonorganic psychotic disorders
Mental and behavioural disorders due to cannabinoids

### 7. Number of unborn children and completed total fertility rate reduction related to disease diagnoses

To calculate the total fertility deficit related to diseases, for the studied cohort, the total number of unborn children per 100,000 people related to a disease in sex ($N_{prevented}$), both directly and indirectly, was calculated using the following equation:

$$N_{prevented} = N \times E(prevalence^{sex_i}) \times \Delta\, p^{sex_i}(childlessness) \times E(parity^{sex_i}) \quad (1)$$

where $E(prevalence^{sex_i})$ is the expected disease prevalence (cumulative incidence rate - CIR) in sex $i$, $\Delta$ $p^{sex_i}(childlessness)$ is the change in the probability of being childless due to a disease diagnosis, and $E(parity^{sex_i})$ is the expected parity in sex $i$ if unaffected (approximated by the average parity in the general population). The $\Delta\, p^{sex_i}(childlessness)$ was calculated using the following equation:

$$\Delta\, p^{sex_i}(childlessness) = (RR^{sex_i} - 1) \times p^{sex_i}(childlessness) \quad (2)$$

where $RR^{sex_i}$ is the risk ratio of childlessness in sex $i$ estimated from the sibling-based analysis and $p^{sex_i}(childlessness)$ is the probability of being childless in sex $i$ if unaffected (approximated by the proportion of childless individuals among the general population). Our approach assumes that the disease only impacts childlessness but not parity, which is supported by our observations (**Table S12**).

For the studied cohort, we also calculated the total fertility rate (TFR) for each sex as the sum of disease ($fr_{affected}^{age_j}$) and non-disease age-specific fertility rates ($fr_{unaffected}^{age_j}$) using the following equation:

$$TFR = \Sigma\, fr_{affected}^{age_j} + \Sigma\, fr_{unaffected}^{age_j} = \Sigma\,(c_{affected}^{age_j}/N^{age_j}) + \Sigma\,(c_{unaffected}^{age_j}/N^{age_j}) \quad (3)$$

where $c_{affected}^{age_j}$ is the total number of children for individuals affected by at least one disease (restricting to disease diagnoses that significantly increased childlessness) at age bracket $j$, $c_{unaffected}^{age_j}$ is the total number of children for unaffected individuals, and $N_{age_j}$ is the total number of individuals at the age bracket $j$.

We presented the reduction of completed TFR due to disease diagnoses as the difference in the TFR between unaffected ($TFR_{unaffected}$) and affected individuals ($TFR_{affected}$), which can be expressed as the following equation:

$$TFR_{unaffected} - TFR_{affected} = \Sigma\,(c_{unaffected}^{age_j}/n_{unaffected}^{age_j}) - \Sigma\,(c_{affected}^{age_j}/n_{affected}^{age_j}) \quad (4)$$

where $n_{unaffected}^{age_j}$ is the number of unaffected individuals at the age bracket $j$, and $n_{affected}^{age_j}$ is the number of affected individuals at the age bracket $j$.

**Table S12:** Relationship of disease diagnoses with parity among women (by age 45) and men (by age 50) with biological children.
(**Table S12** is too large therefore see sheet "S12_EffectOnParity" from "SUBMIT_TableS7_S9_S11_S12_S17_DiseaseSpecificChildlessness.xlsx")

**Table S13:** Number of children unborn per 100,000 people related to disease diagnoses.

| Disease | Sex | Category | N children | Lower 95%CI | Upper 95%CI |
|---|---|---|---|---|---|
| Schizophrenia | Women | Mental-behavioural | 612 | 550 | 670 |
| Acute and transient psychotic disorders | Women | Mental-behavioural | 389 | 314 | 462 |
| Mood disorders | Women | Mental-behavioural | 389 | 313 | 470 |
| Epilepsy | Women | Nervous system | 381 | 328 | 434 |
| Other and unspecified nonorganic psychotic disorders | Women | Mental-behavioural | 375 | 323 | 423 |
| Cerebral palsy | Women | Nervous system | 339 | 269 | 396 |
| Anxiety disorders | Women | Mental-behavioural | 257 | 192 | 325 |
| Mild intellectual disability | Women | Mental-behavioural | 234 | 190 | 267 |
| Type 1 diabetes | Women | Endocrine-nutritional-metabolic | 198 | 159 | 240 |

| | | | | | |
|---|---|---|---|---|---|
| Other specified and unspecified personality disorders | Women | Mental-behavioural | 150 | 117 | 183 |
| Persistent mood disorders | Women | Mental-behavioural | 121 | 91 | 152 |
| Bipolar affective disorders | Women | Mental-behavioural | 112 | 85 | 142 |
| Emotionally unstable personality disorder | Women | Mental-behavioural | 112 | 83 | 142 |
| Other neurotic disorders | Women | Mental-behavioural | 108 | 80 | 137 |
| Congenital malformations of the circulatory system | Women | Congenital anomalies | 107 | 67 | 146 |
| Schizoaffective disorder | Women | Mental-behavioural | 103 | 76 | 127 |
| Persistent delusional disorders | Women | Mental-behavioural | 97 | 59 | 126 |
| Other mental disorders due to brain damage and dysfunction and to physical disease | Women | Mental-behavioural | 94 | 56 | 120 |
| Pneumonia, organism unspecified | Women | Respiratory system | 91 | 58 | 124 |
| Congenital anomalies of heart | Women | Congenital anomalies | 85 | 49 | 120 |
| Other anxiety disorders | Women | Mental-behavioural | 77 | 53 | 102 |
| Anorexia (Inc. Atypical) | Women | Mental-behavioural | 75 | 52 | 101 |
| Juvenile idiopathic arthritis | Women | Musculoskeletal system | 72 | 50 | 95 |
| Malignant neoplasm of brain | Women | Neoplasms | 61 | 32 | 88 |
| Alcohol dependence | Women | Mental-behavioural | 57 | 37 | 80 |
| Acute upper respiratory infections of multiple and unspecified sites | Women | Respiratory system | 55 | 29 | 84 |
| Other juvenile arthritis | Women | Musculoskeletal system | 54 | 27 | 81 |
| Anxious personality disorder | Women | Mental-behavioural | 53 | 27 | 78 |
| Congenital anomalies of musculoskeletal system | Women | Congenital anomalies | 50 | 24 | 79 |
| Type 2 diabetes | Women | Endocrine-nutritional-metabolic | 49 | 23 | 77 |
| Mental and behavioural disorders due to opioids | Women | Mental-behavioural | 48 | 23 | 74 |
| Obesity, other/unspecified | Women | Endocrine-nutritional-metabolic | 47 | 27 | 69 |
| Congenital anomalies of nervous system | Women | Congenital anomalies | 47 | 21 | 71 |
| Congenital anomalies of digestive system | Women | Congenital anomalies | 43 | 19 | 67 |
| Other reaction to severe stress, and adjustment disorders | Women | Mental-behavioural | 42 | 22 | 64 |
| Other cataract | Women | Eye and adnexa | 42 | 25 | 59 |
| Other septicaemia | Women | Infectious-parasitic | 40 | 17 | 67 |
| Other and unspecified mood [affective] disorders | Women | Mental-behavioural | 40 | 18 | 65 |
| Myocarditis | Women | Circulatory system | 38 | 21 | 56 |
| Acute alcohol intoxication | Women | Mental-behavioural | 37 | 21 | 56 |
| Mental and behavioural disorders due to sedatives or hypnotics | Women | Mental-behavioural | 31 | 16 | 47 |
| Other or unspecified ileus, impaction, or obstruction | Women | Digestive system | 29 | 15 | 46 |
| Chronic tubulo-interstitial nephritis | Women | Genitourinary system | 29 | 14 | 47 |
| Hypertension, essential | Women | Circulatory system | 28 | 13 | 45 |
| Other arrhythmias | Women | Circulatory system | 28 | 14 | 44 |
| Mental and behavioural disorders due to multiple drug use and use of other psychoactive substances | Women | Mental-behavioural | 26 | 13 | 41 |
| Obsessive-compulsive disorder | Women | Mental-behavioural | 26 | 13 | 40 |
| Panic disorder | Women | Mental-behavioural | 25 | 12 | 40 |
| Multiple Sclerosis | Women | Nervous system | 23 | 10 | 38 |

| | | | | | |
|---|---|---|---|---|---|
| Other specified/unspecified hearing loss | Women | Ear and mastoid process | 22 | 10 | 35 |
| Systemic lupus erythematosus | Women | Musculoskeletal system | 20 | 9 | 34 |
| Schizophrenia | Men | Mental-behavioural | 893 | 849 | 929 |
| Anxiety disorders | Men | Mental-behavioural | 569 | 477 | 663 |
| Other and unspecified nonorganic psychotic disorders | Men | Mental-behavioural | 420 | 380 | 457 |
| Acute and transient psychotic disorders | Men | Mental-behavioural | 385 | 326 | 438 |
| Epilepsy | Men | Nervous system | 373 | 327 | 417 |
| Mood disorders | Men | Mental-behavioural | 368 | 295 | 444 |
| Alcohol dependence | Men | Mental-behavioural | 367 | 319 | 413 |
| Acute alcohol intoxication | Men | Mental-behavioural | 272 | 234 | 309 |
| Mild intellectual disability | Men | Mental-behavioural | 260 | 220 | 283 |
| Cerebral palsy | Men | Nervous system | 172 | 136 | 197 |
| Other neurotic disorders | Men | Mental-behavioural | 163 | 132 | 195 |
| Other specified and unspecified personality disorders | Men | Mental-behavioural | 163 | 132 | 194 |
| Other anxiety disorders | Men | Mental-behavioural | 157 | 125 | 193 |
| Schizoid personality disorder | Men | Mental-behavioural | 151 | 119 | 168 |
| Type 1 diabetes | Men | Endocrine-nutritional-metabolic | 143 | 107 | 181 |
| Bipolar affective disorders | Men | Mental-behavioural | 137 | 111 | 161 |
| Persistent mood disorders | Men | Mental-behavioural | 113 | 82 | 143 |
| Mental and behavioural disorders due to opioids | Men | Mental-behavioural | 94 | 62 | 126 |
| Other mental disorders due to brain damage and dysfunction and to physical disease | Men | Mental-behavioural | 77 | 58 | 94 |
| Schizoaffective disorder | Men | Mental-behavioural | 72 | 54 | 87 |
| Persistent delusional disorders | Men | Mental-behavioural | 67 | 42 | 88 |
| Malignant neoplasm of brain | Men | Neoplasms | 66 | 41 | 87 |
| Emotionally unstable personality disorder | Men | Mental-behavioural | 66 | 46 | 85 |
| Mental and behavioural disorders due to multiple drug use and use of other psychoactive substances | Men | Mental-behavioural | 63 | 42 | 84 |
| Myopia | Men | Eye and adnexa | 62 | 37 | 85 |
| Obesity, other/unspecified | Men | Endocrine-nutritional-metabolic | 61 | 42 | 81 |
| Convergent concomitant strabismus | Men | Eye and adnexa | 60 | 37 | 83 |
| Anisometropia and aniseikonia | Men | Eye and adnexa | 60 | 36 | 82 |
| Other reaction to severe stress, and adjustment disorders | Men | Mental-behavioural | 59 | 37 | 83 |
| Acute pancreatitis | Men | Digestive system | 59 | 31 | 90 |
| Presbyopia | Men | Eye and adnexa | 55 | 31 | 77 |
| Mental and behavioural disorders due to use of other stimulants, including caffeine | Men | Mental-behavioural | 54 | 35 | 73 |
| Congenital malformations of the circulatory system | Men | Congenital anomalies | 50 | 26 | 73 |
| Mental and behavioural disorders due to cannabinoids | Men | Mental-behavioural | 47 | 29 | 65 |
| Congenital anomalies of heart | Men | Congenital anomalies | 47 | 24 | 70 |
| Behavioural disorders | Men | Mental-behavioural | 46 | 22 | 70 |
| Obsessive-compulsive disorder | Men | Mental-behavioural | 45 | 31 | 57 |
| Divergent concomitant strabismus | Men | Eye and adnexa | 42 | 22 | 63 |
| Mental and behavioural disorders due to sedatives or hypnotics | Men | Mental-behavioural | 41 | 25 | 57 |

| | | | | | |
|---|---|---|---|---|---|
| Phobic anxiety disorders | Men | Mental-behavioural | 40 | 25 | 56 |
| Post-traumatic stress disorder | Men | Mental-behavioural | 37 | 17 | 57 |
| Panic disorder | Men | Mental-behavioural | 34 | 19 | 48 |
| Other specified/unspecified hearing loss | Men | Ear and mastoid process | 34 | 19 | 49 |
| Generalized anxiety disorder | Men | Mental-behavioural | 31 | 16 | 46 |
| Status epilepticus | Men | Nervous system | 29 | 17 | 39 |
| Heart failure, strict | Men | Circulatory system | 29 | 16 | 42 |
| Other or ill-defined heart diseases | Men | Circulatory system | 26 | 14 | 38 |
| Subarachnoid haemorrhage | Men | Circulatory system | 26 | 14 | 38 |

**Table S14:** Number of children unborn per 100,000 people by disease category and across all disease diagnoses, from people affected by at least one disease.

| **Category** | **N children** | **Lower 95%CI** | **Upper 95%CI** |
|---|---|---|---|
| ***Men*** | | | |
| Mental-behavioural | 2728 | 2586 | 2871 |
| Nervous system | 526 | 469 | 581 |
| Endocrine-nutritional-metabolic | 209 | 166 | 253 |
| Eye and adnexa | 169 | 125 | 214 |
| Circulatory system | 83 | 59 | 106 |
| Neoplasms | 65 | 40 | 86 |
| Digestive system | 55 | 27 | 85 |
| Ear and mastoid process | 51 | 29 | 73 |
| Congenital anomalies | 50 | 26 | 73 |
| **All** | **4032** | **3845** | **4219** |
| ***Women*** | | | |
| Mental-behavioural | 1489 | 1376 | 1605 |
| Nervous system | 596 | 525 | 668 |
| Endocrine-nutritional-metabolic | 255 | 207 | 305 |
| Congenital anomalies | 220 | 163 | 278 |
| Respiratory system | 192 | 135 | 252 |
| Musculoskeletal system | 121 | 88 | 155 |
| Circulatory system | 111 | 79 | 146 |
| Neoplasms | 60 | 32 | 88 |
| Genitourinary system | 50 | 24 | 80 |
| Eye and adnexa | 46 | 27 | 67 |
| Infectious-parasitic | 42 | 18 | 69 |
| Ear and mastoid process | 29 | 12 | 48 |
| Digestive system | 19 | 9 | 30 |
| **All** | **2539** | **2382** | **2698** |

**Table S15**: Estimated contribution of diseases to completed total fertility rates (TFR) in Finland.

| | **All** | | | **15-19 y.o.** | | | **20-24 y.o.** | | | **25-29 y.o.** | | | **30-34 y.o.** | | | **35-39 y.o.** | | | **40-44 y.o.** | | | **45-49 y.o.** | | |
|---|---|---|---|---|---|---|---|---|---|---|---|---|---|---|---|---|---|---|---|---|---|---|---|---|
| | **Fr** | **Low 95% CI** | **Up 95% CI** | **Fr** | **Low 95% CI** | **Up 95% CI** | **Fr** | **Low 95% CI** | **Up 95% CI** | **Fr** | **Low 95% CI** | **Up 95% CI** | **Fr** | **Low 95% CI** | **Up 95% CI** | **Fr** | **Low 95% CI** | **Up 95% CI** | **Fr** | **Low 95% CI** | **Up 95% CI** | **Fr** | **Low 95% CI** | **Up 95% CI** |
| ***Men*** | | | | | | | | | | | | | | | | | | | | | | | | |
| TFR | 1.730 | 1.730 | 1.730 | 0.037 | 0.036 | 0.037 | 0.281 | 0.279 | 0.282 | 0.562 | 0.56 | 0.563 | 0.483 | 0.481 | 0.484 | 0.251 | 0.25 | 0.252 | 0.092 | 0.091 | 0.093 | 0.025 | 0.024 | 0.025 |
| TFR, unaffected | 1.650 | 1.650 | 1.650 | 0.036 | 0.036 | 0.037 | 0.273 | 0.272 | 0.275 | 0.54 | 0.538 | 0.541 | 0.46 | 0.459 | 0.461 | 0.235 | 0.234 | 0.236 | 0.084 | 0.083 | 0.085 | 0.022 | 0.021 | 0.022 |
| TFR, affected | 0.080 | 0.080 | 0.080 | 0.001 | 0.000 | 0.001 | 0.007 | 0.007 | 0.008 | 0.022 | 0.022 | 0.023 | 0.023 | 0.022 | 0.023 | 0.016 | 0.015 | 0.016 | 0.008 | 0.008 | 0.008 | 0.003 | 0.003 | 0.003 |
| TFR, affected (%) | 4.614 | 4.614 | 4.615 | 1.52 | 1.354 | 1.68 | 2.651 | 2.574 | 2.727 | 3.951 | 3.885 | 4.017 | 4.723 | 4.647 | 4.799 | 6.189 | 6.076 | 6.301 | 8.897 | 8.69 | 9.1 | 12.505 | 12.065 | 12.928 |
| ***Women*** | | | | | | | | | | | | | | | | | | | | | | | | |
| TFR | 1.921 | 1.921 | 1.921 | 0.139 | 0.138 | 0.14 | 0.454 | 0.453 | 0.455 | 0.648 | 0.647 | 0.649 | 0.459 | 0.457 | 0.46 | 0.19 | 0.189 | 0.191 | 0.031 | 0.03 | 0.031 | - | | |
| TFR, unaffected | 1.816 | 1.816 | 1.816 | 0.135 | 0.134 | 0.136 | 0.436 | 0.434 | 0.437 | 0.618 | 0.616 | 0.619 | 0.43 | 0.428 | 0.431 | 0.172 | 0.171 | 0.173 | 0.026 | 0.026 | 0.027 | - | | |
| TFR, affected | 0.104 | 0.104 | 0.104 | 0.004 | 0.004 | 0.004 | 0.018 | 0.018 | 0.019 | 0.03 | 0.03 | 0.031 | 0.029 | 0.029 | 0.03 | 0.018 | 0.018 | 0.018 | 0.005 | 0.004 | 0.005 | - | | |
| TFR, affected (%) | 5.440 | 5.440 | 5.440 | 2.863 | 2.763 | 2.961 | 4.073 | 4.007 | 4.138 | 4.652 | 4.592 | 4.711 | 6.366 | 6.288 | 6.443 | 9.539 | 9.407 | 9.669 | 14.642 | 14.284 | 14.989 | - | | |

**Table S16**: Age-specific fertility rates by disease status (Finland only).

| | **All** | | | **15-19 y.o.** | | | **20-24 y.o.** | | | **25-29 y.o.** | | | **30-34 y.o.** | | | **35-39 y.o.** | | | **40-44 y.o.** | | | **45-49 y.o.** | | |
|---|---|---|---|---|---|---|---|---|---|---|---|---|---|---|---|---|---|---|---|---|---|---|---|---|
| | **Fr** | **Low 95% CI** | **Up 95% CI** | **Fr** | **Low 95% CI** | **Up 95% CI** | **Fr** | **Low 95% CI** | **Up 95% CI** | **Fr** | **Low 95% CI** | **Up 95% CI** | **Fr** | **Low 95% CI** | **Up 95% CI** | **Fr** | **Low 95% CI** | **Up 95% CI** | **Fr** | **Low 95% CI** | **Up 95% CI** | **Fr** | **Low 95% CI** | **Up 95% CI** |
| ***Men*** | | | | | | | | | | | | | | | | | | | | | | | | |
| TFR, unaffected | 1.785 | 1.785 | 1.785 | 0.037 | 0.036 | 0.037 | 0.283 | 0.281 | 0.284 | 0.576 | 0.574 | 0.577 | 0.502 | 0.5 | 0.503 | 0.264 | 0.263 | 0.265 | 0.098 | 0.097 | 0.099 | 0.026 | 0.026 | 0.027 |
| TFR, affected | 1.105 | 1.105 | 1.105 | 0.035 | 0.031 | 0.039 | 0.223 | 0.216 | 0.229 | 0.355 | 0.35 | 0.361 | 0.274 | 0.27 | 0.279 | 0.142 | 0.139 | 0.145 | 0.059 | 0.057 | 0.060 | 0.017 | 0.016 | 0.018 |
| Reduction TFR, unaffected -> TFR, affected (%) | 38.074 | 38.078 | 38.069 | 5.722 | 15.752 | -4.015 | 21.224 | 23.188 | 19.279 | 38.264 | 39.07 | 37.461 | 45.311 | 46.044 | 44.583 | 46.063 | 46.959 | 45.177 | 40.169 | 41.484 | 38.878 | 34.243 | 36.445 | 32.126 |
| ***Women*** | | | | | | | | | | | | | | | | | | | | | | | | |
| TFR, unaffected | 1.951 | 1.951 | 1.951 | 0.139 | 0.138 | 0.14 | 0.456 | 0.455 | 0.458 | 0.658 | 0.657 | 0.66 | 0.47 | 0.469 | 0.471 | 0.196 | 0.195 | 0.197 | 0.032 | 0.031 | 0.032 | - | | |
| TFR, affected | 1.543 | 1.543 | 1.543 | 0.128 | 0.123 | 0.133 | 0.409 | 0.403 | 0.415 | 0.489 | 0.483 | 0.494 | 0.34 | 0.336 | 0.344 | 0.15 | 0.147 | 0.153 | 0.027 | 0.026 | 0.028 | - | | |
| Reduction TFR, unaffected -> TFR, affected (%) | 20.906 | 20.909 | 20.903 | 7.754 | 10.711 | 4.836 | 10.272 | 11.33 | 9.221 | 25.776 | 26.428 | 25.127 | 27.577 | 28.264 | 26.894 | 23.427 | 24.364 | 22.499 | 16.021 | 17.936 | 14.165 | - | | |

### 8. Mediation effect by singlehood

Similar to our previous childlessness analysis, we estimated the effect of 403 disease diagnoses on the risk of being without a partner by age 45 in women and 50 in men. For index individuals in Finland, we obtained their longitudinal marriage and partnership information between 11 April 1971 and 31 December 2018 from the Population Information System. In Sweden, we collected information of married couples and cohabiting unions with biological children between 1 January 1977 and 31 December 2017 from Statistics Sweden. We defined individuals as partnerless if they did not have any abovementioned marriage or partnership registered by age 45 (women) or 50 (men).

### 8.1. Effects on singlehood

**Figure S5.** Relationship of 402 disease diagnoses with being without a partner by age 45 in women (Panel A) and 50 in men (Panel B). Only disease diagnoses that are significantly associated after multiple-testing correction are colored.

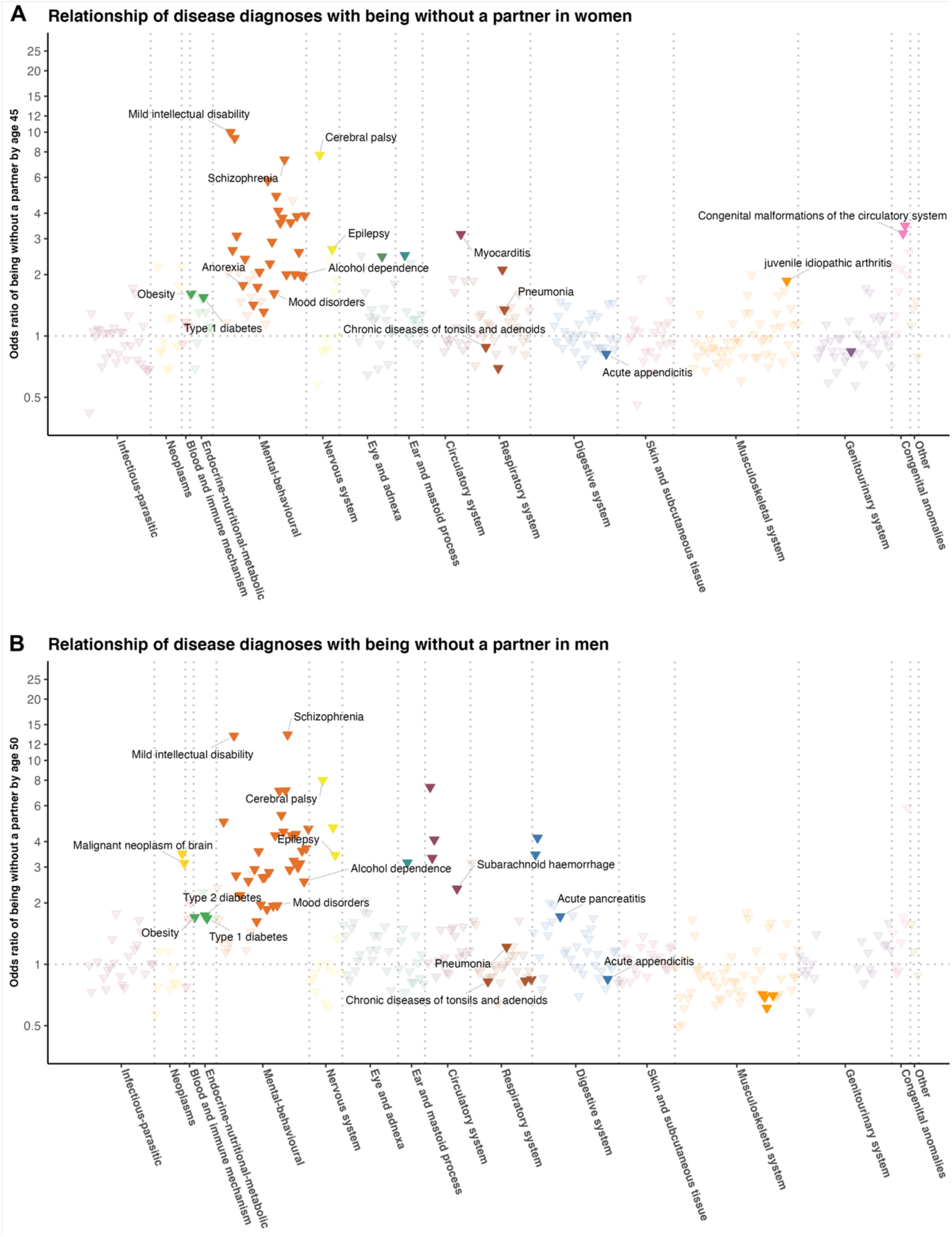

**Figure S6.** Sex-specific effects for the association between disease diagnoses and being without a partner by age 45 in women and 50 in men, for 68 disease diagnoses that were significantly associated with increased childlessness from the main analysis. Only disease diagnoses that are significantly different between sexes at a nominal P value are colored.

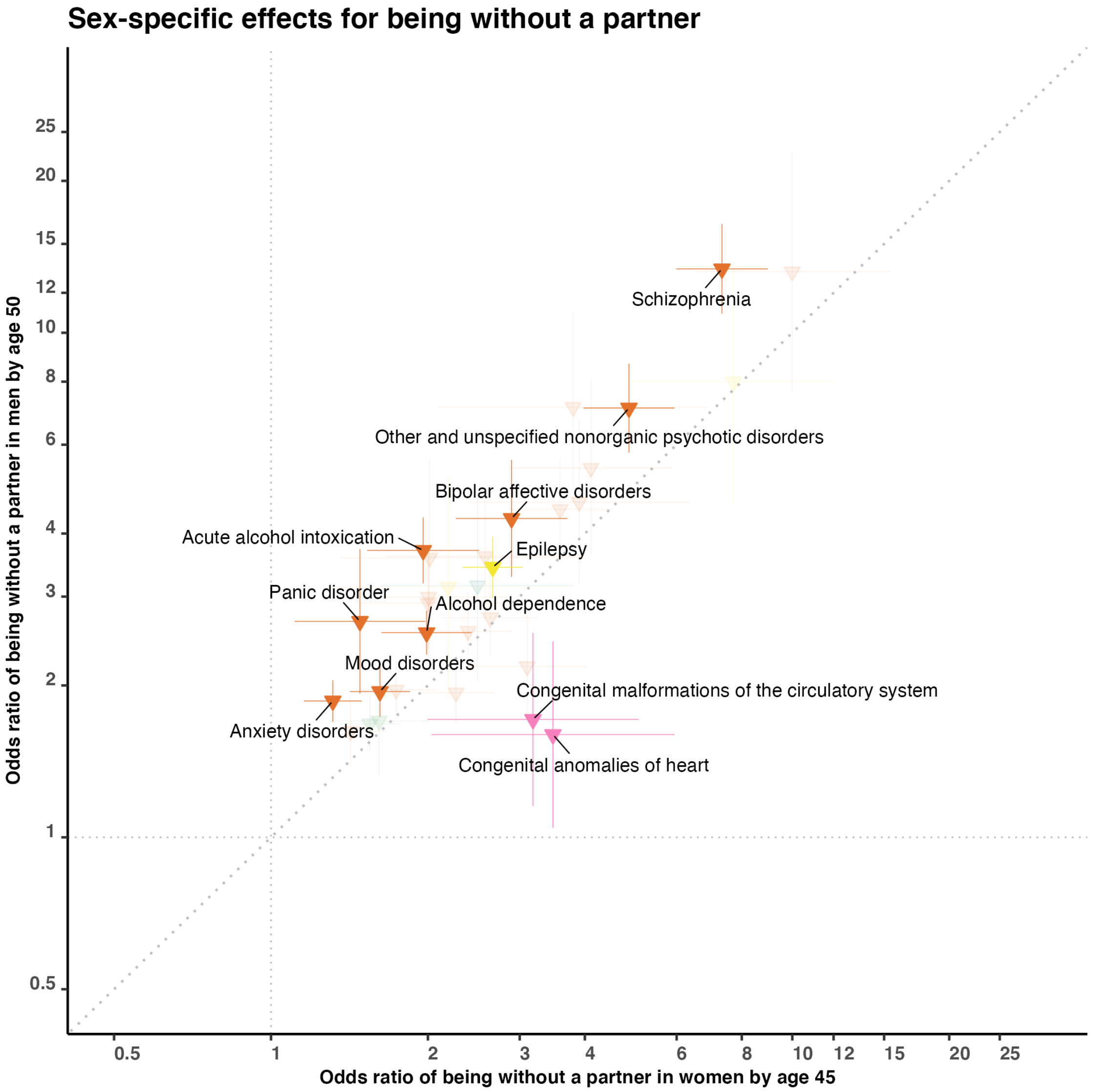

#### 8.2. Mediation analysis

To quantify the overall effect of singlehood on the association between disease diagnoses and childlessness, we performed linear regression for each sex using all disease diagnoses that were significantly associated with childlessness, with log ORs of disease diagnoses on childlessness as the dependent variable and log ORs on singlehood as the independent variable (**Figure S7**). High $R^2$ was observed in both men ($R^2$=0.85, regression coefficient =1.08, P=3.5×$10^{-23}$) and women ($R^2$=0.71, regression coefficient =0.96, P=1.0×$10^{-15}$). Also, the intercept was significant in women (0.42, P=1.0×$10^{-15}$) but not in men (0.11, P=0.13), demonstrating that there were additional effects not fully captured by union formation in women.

**Figure S7.** Relationship of disease diagnoses with childlessness (main analysis) versus singlehood by age 45 in women (Panel A) and 50 in men (Panel B), for 74 disease diagnoses that were significantly associated with childlessness from the main analysis. Only disease diagnoses that are significantly different between two outcomes at a nominal P value are colored.

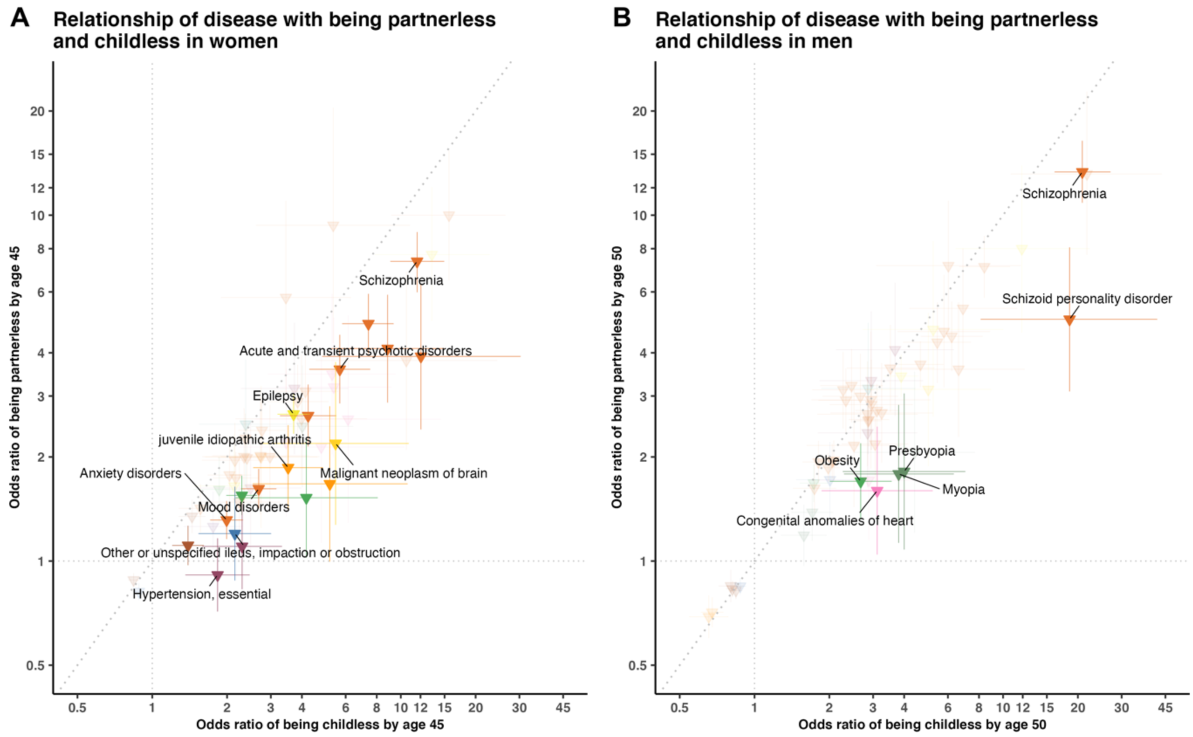


For each disease diagnosis, we performed a causal mediation analysis, with singlehood as a dichotomous latent factor. A median of 29.3% of the disease effects on childlessness in women and 37.9% in men was mediated by singlehood (**Table S17**). Different patterns were observed across diseases. For example, singlehood was a significant mediator for women diagnosed with schizophrenia (OR=2.1 [2.0-2.2] for indirect and 6.8 [5.0-9.2] for direct effect), but not for women with hypertension (1.2 [1.1-1.4] for indirect and 2.4 [1.6-3.5] for direct effect).

**Table S17**: Direct and indirect effects (mediated by singlehood) of disease on childlessness estimated from mediation analysis.

(**Table S17** is too large therefore see sheet "S17_MediationAnalysis" from "SUBMIT_TableS7_S9_S11_S12_S17_DiseaseSpecificChildlessness.xlsx")

#### 8.3. Effects among individuals with registered partners

**Figure S8.** Relationship of 169 disease diagnoses with childlessness among women (by age 45) (Panel A) and men (by age 50) (Panel B) with registered partners. Only disease diagnoses that are significantly associated after multiple-testing correction are colored.

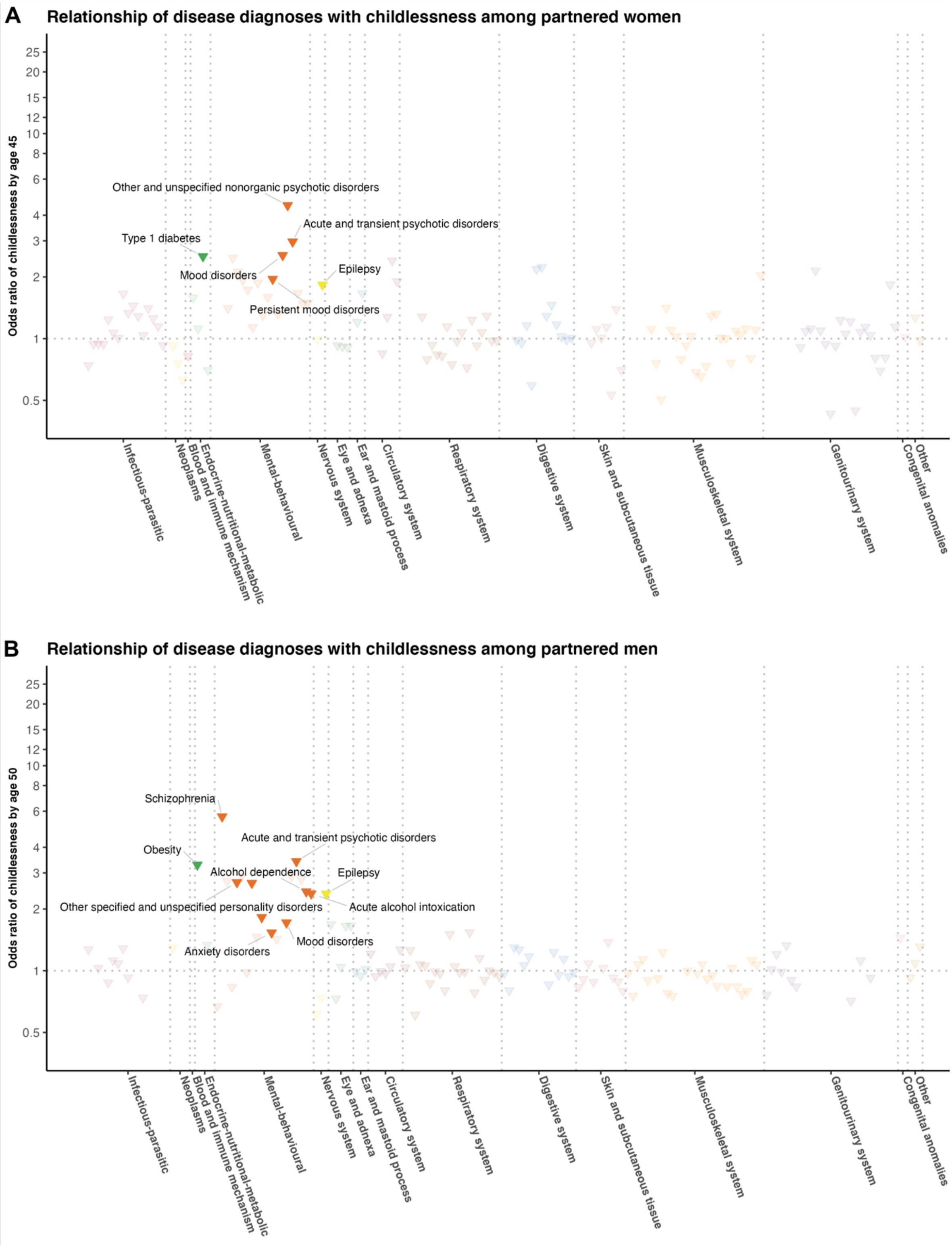

**Figure S9:** Relationship of disease diagnoses with childlessness by age 45 in women (Panel A) and 50 in men (Panel B) from the general population (main analysis) versus people with registered partners, for 36 disease diagnoses that were significantly associated with childlessness from the main analysis.

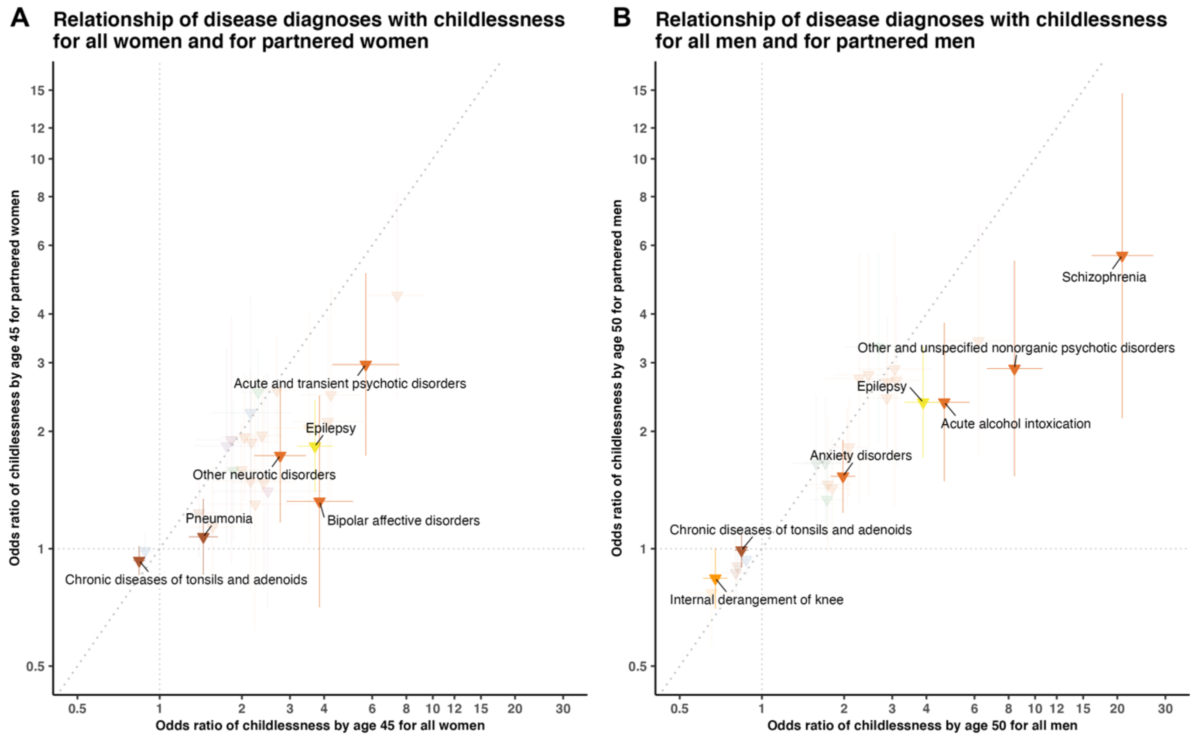

### 9. Population-based and sensitivity analyses

To estimate the population-level effect of diseases on childlessness we performed a nested incident-matched case-control design which matched each childless case to one control based on sex, birth year, municipality of birth (545 from Finland and 1,091 from Sweden), and the highest parental education level (International Standard Classification of Education 1997). For the matching process, among people with the same birth municipality and parental education background, we firstly searched for the individual born at the same year as the control and then extended to the individual with the closest birth year (up to 5 years of difference). There was no overlap among matched controls. Similar to the sibling-based analysis, we only considered the disease diagnosis onset at least one year before the age at first birth of the matched control and examined associations between diseases and childlessness using a conditional logistic regression model. In total, we used 226,860 matched pairs of women (107,375 in Finland and 119,485 in Sweden) and 274,941 of men (127,689 in Finland and 147,252 in Sweden) for the population-based analysis.

**Figure S10.** Relationship of 403 disease diagnoses with childlessness by age 45 in women (Panel A) and 50 in men (Panel B) from using a nested incident-matched case-control design, matched by birth year, birth municipality, and highest parental education level. Only disease diagnoses that are significantly associated after multiple-testing correction are colored.

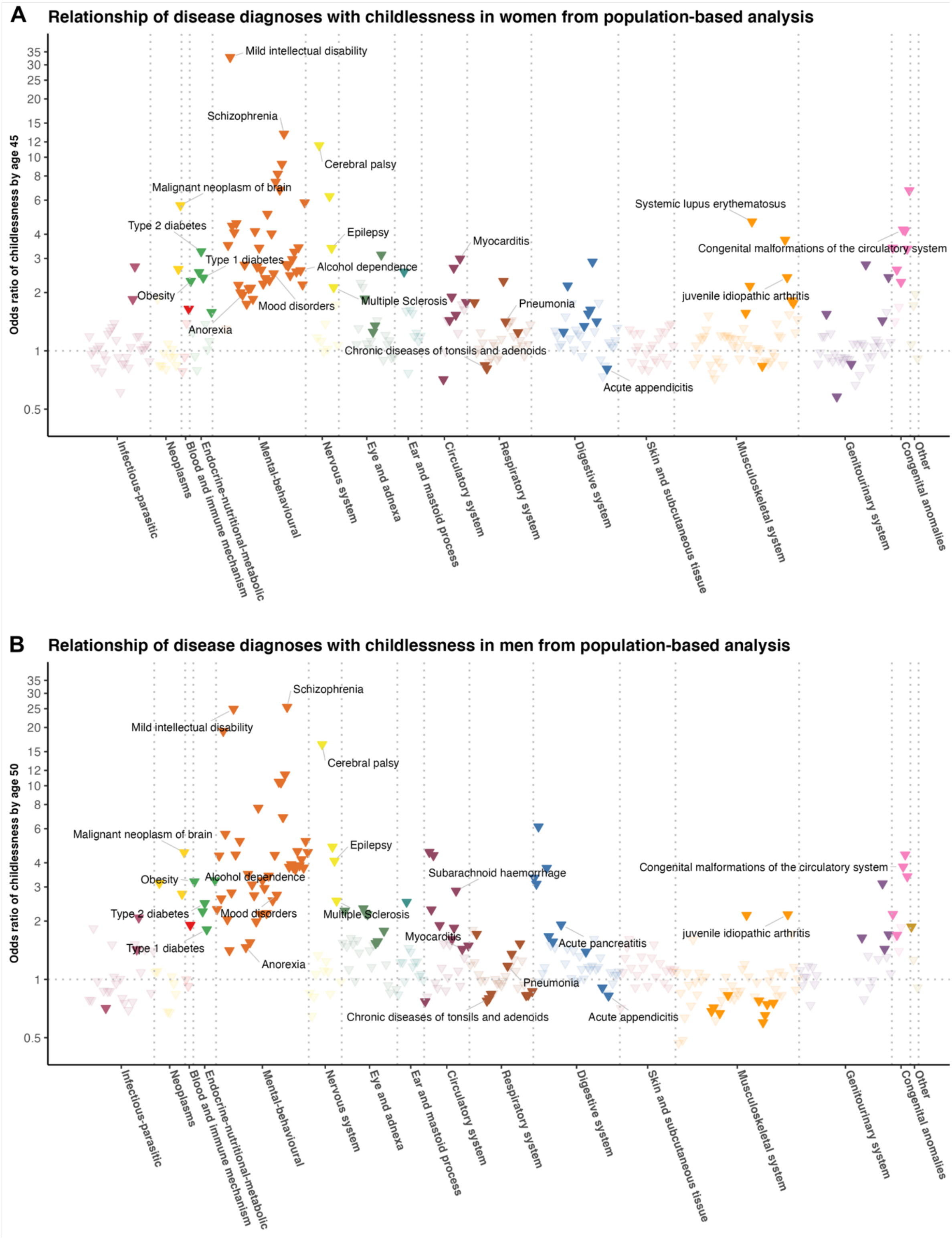

**Figure S11.** Relationship of disease diagnoses with childlessness by age 45 in women (Panel A) and 50 in men (Panel B) from the sibling-based (main analysis) versus population-based analyses, for 74 disease diagnoses that were significantly associated with childlessness from the main analysis.

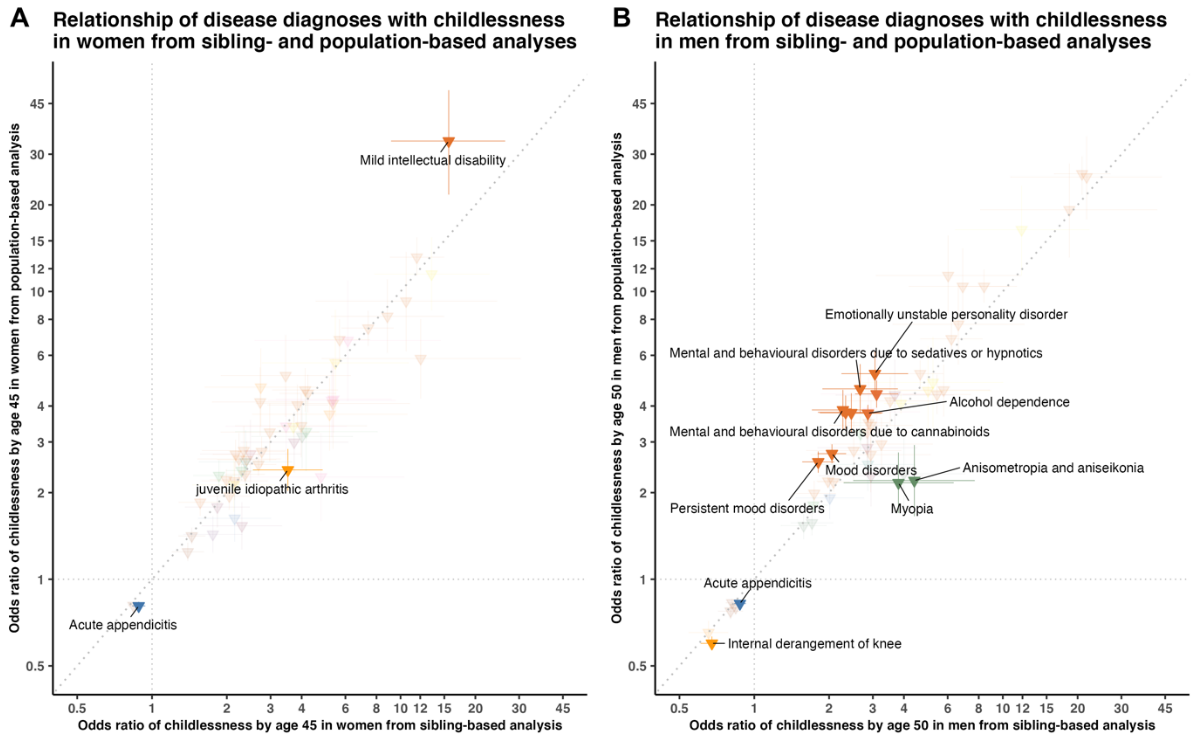

**Figure S12.** Relationship of 365 disease diagnoses with childlessness restricted to women alive by age 45 (Panel A) and men by 50 (Panel B), using a sibling-based design. Only disease diagnoses that are significantly associated after multiple-testing correction are colored.

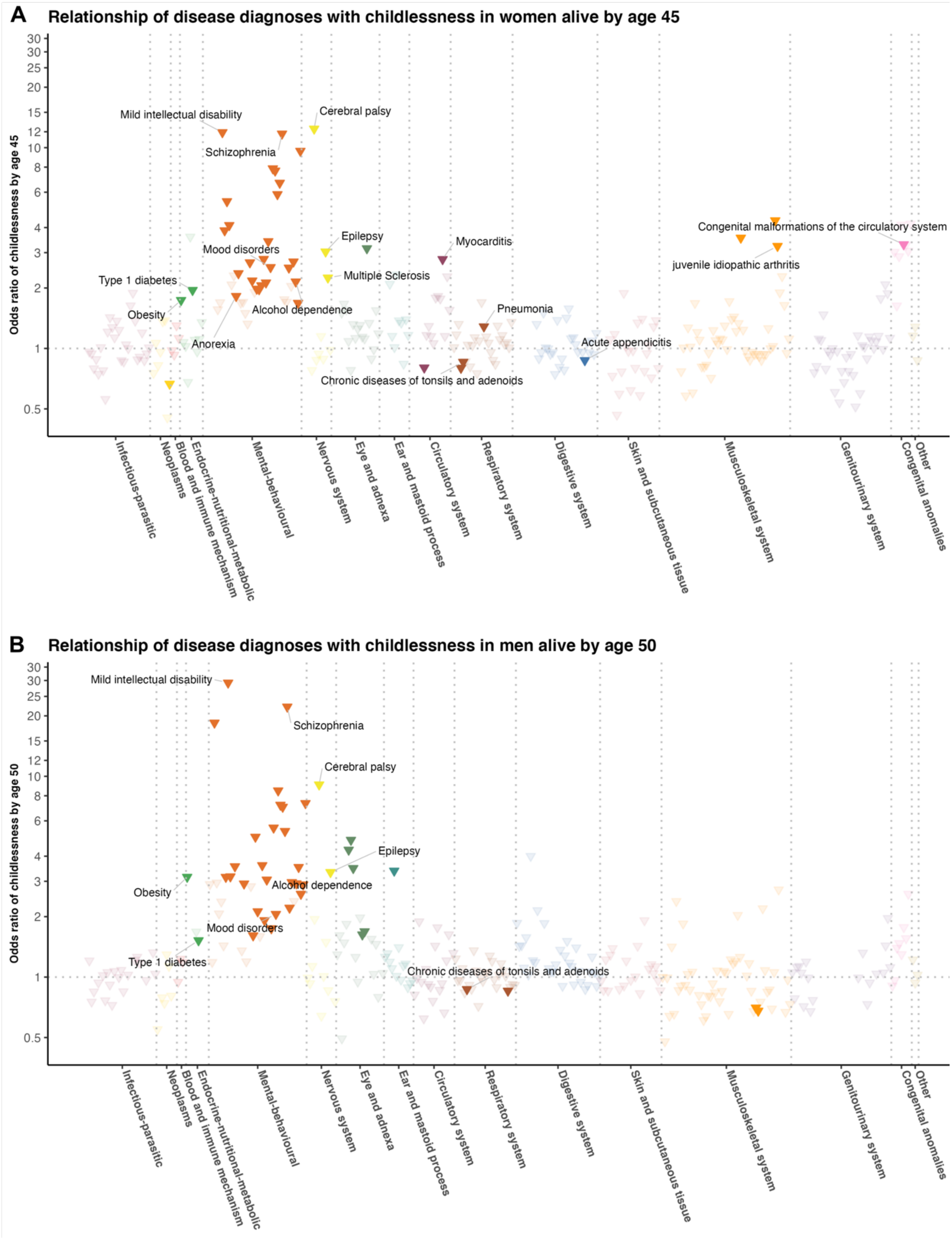

**Figure S13.** Relationship of disease diagnoses with childlessness in the general population (main analysis) versus the sensitivity analysis which restricted to women alive by age 45 (Panel A) and men by 50 (Panel B), for 71 disease diagnoses that were significantly associated with childlessness from the main analysis.

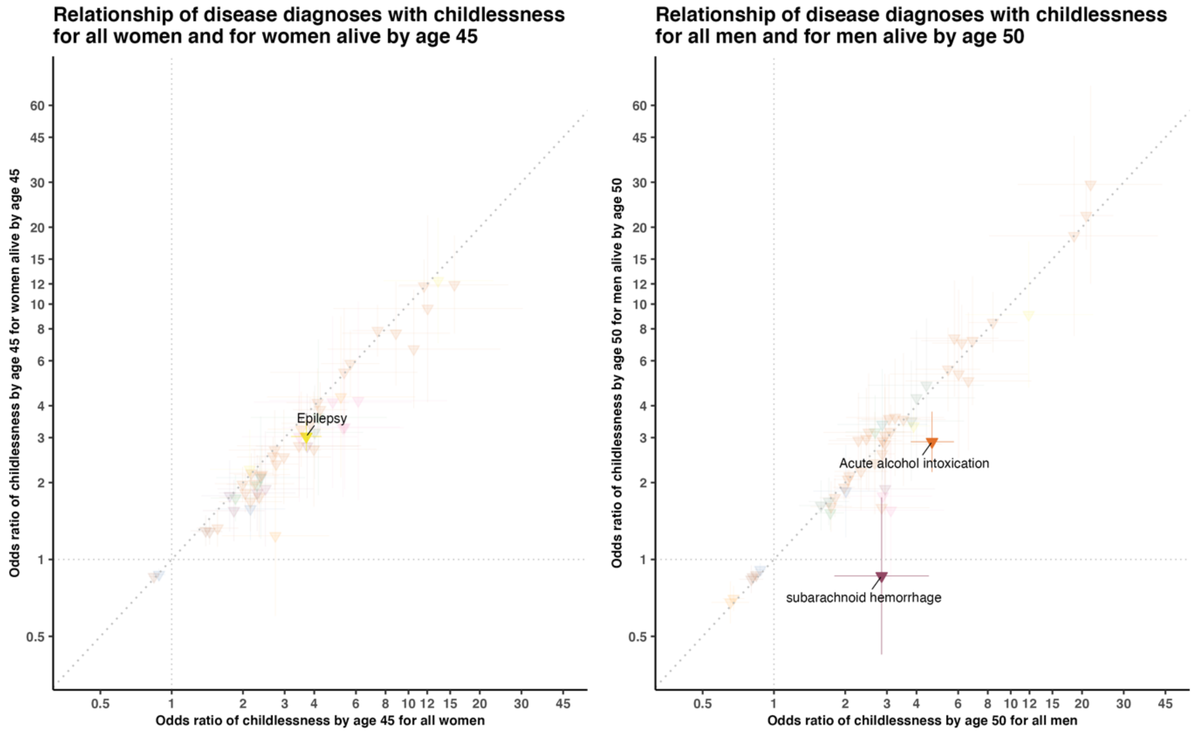

**Figure S14:** Relationship of 403 disease diagnoses with childlessness by age 45 in women (Panel A) and 50 in men (Panel B) from using a Cox proportional hazards regression model stratified by same-sex full-siblings. Only disease diagnoses that are significantly associated after multiple-testing correction are colored.

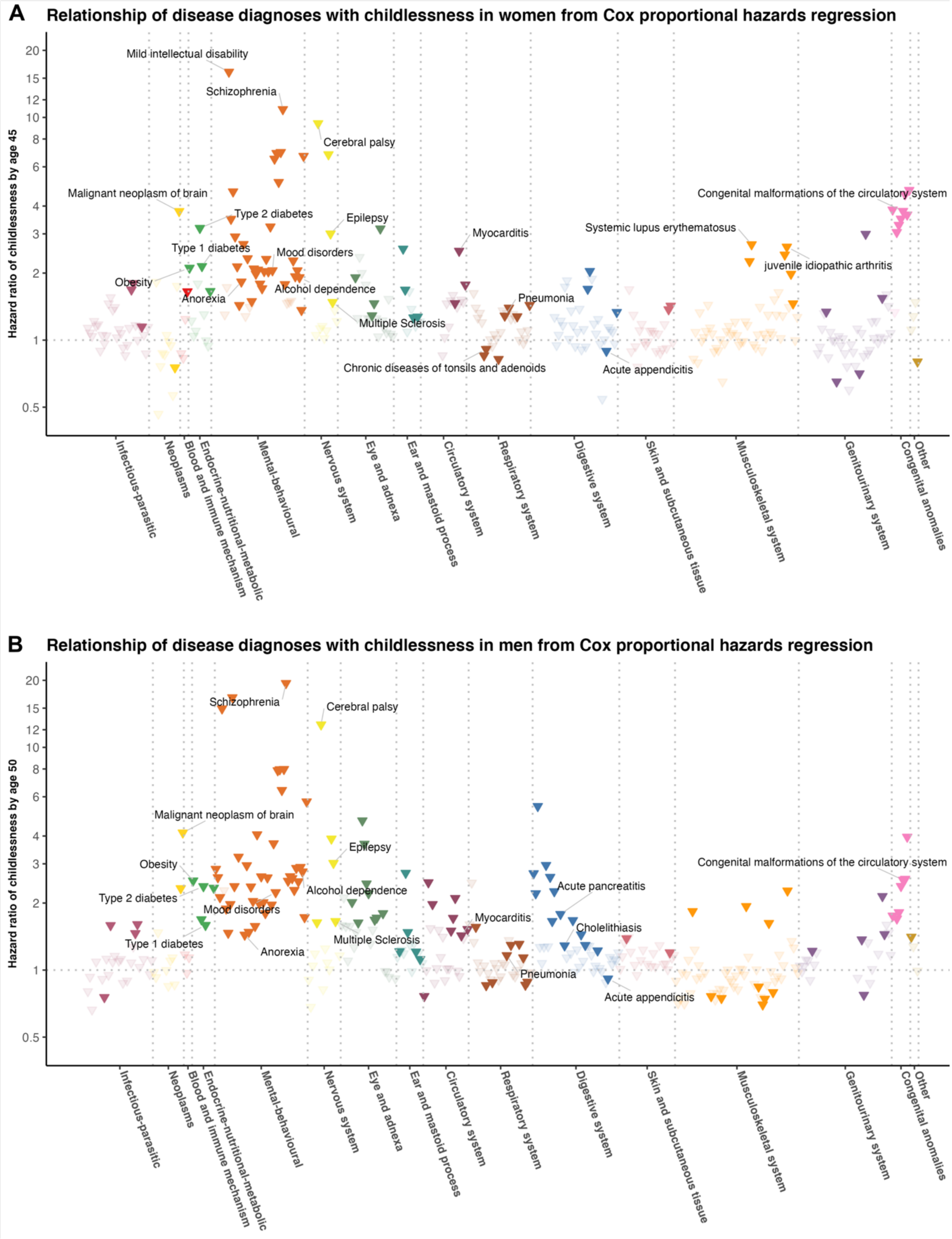

**Figure S15:** Relationship of disease diagnoses with childlessness by age 45 in women (Panel A) and 50 in men (Panel B) from conditional logistic regression (main analysis) versus stratified Cox proportional hazards regression, for 74 disease diagnoses that were significantly associated with childlessness from the main analysis.

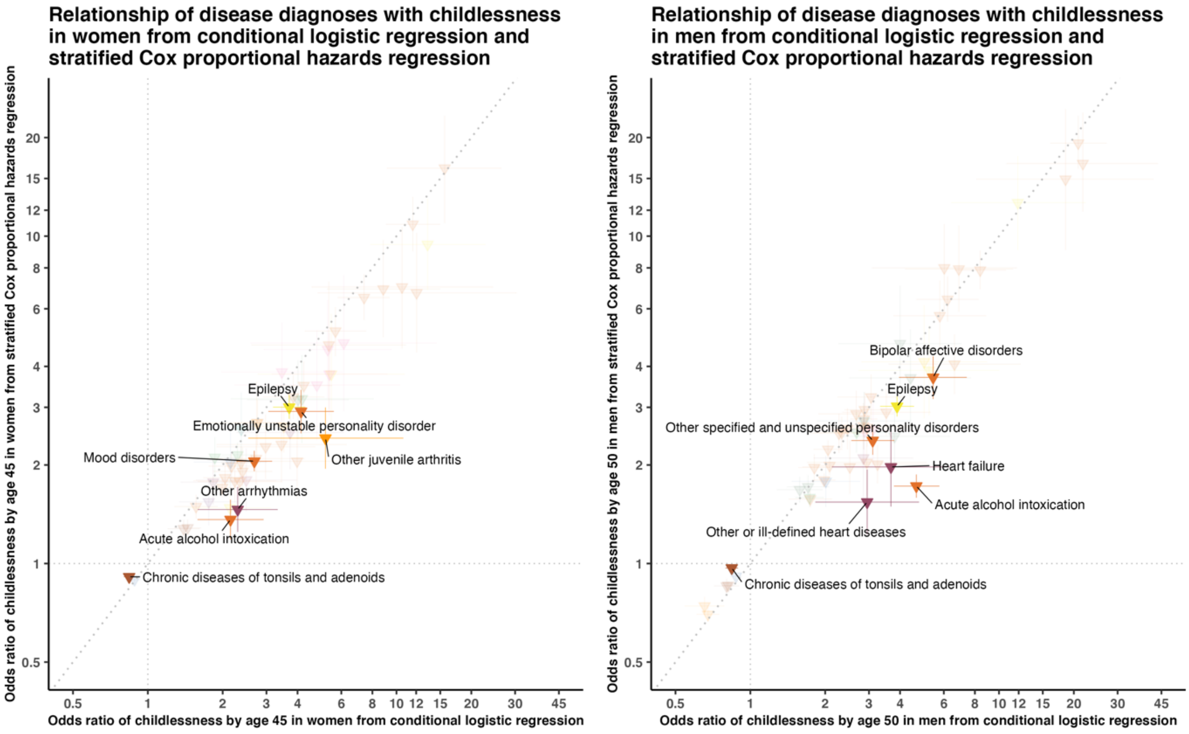

**Figure S16:** Relationship of disease diagnoses with childlessness by age 45 in women (Panel A) and 50 in men (Panel B) from main analysis versus sensitivity analysis adjusting for individuals' highest education level, for 74 disease diagnoses that were significantly associated with childlessness from the main analysis.

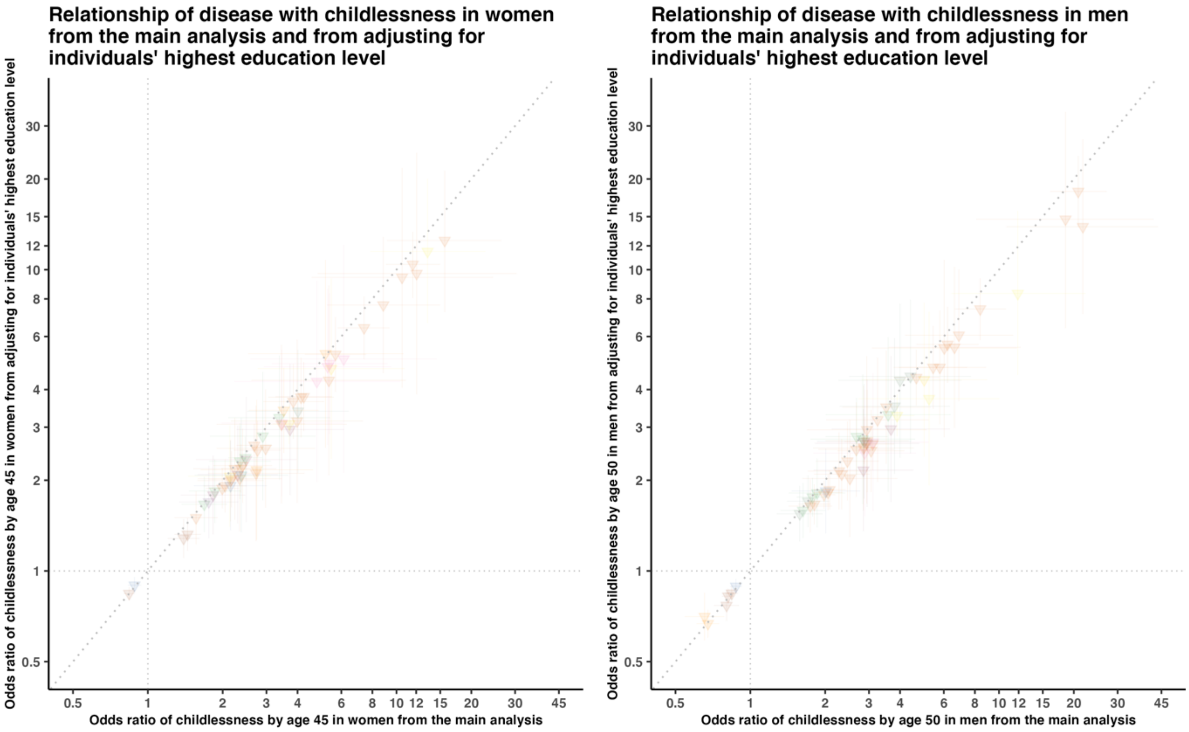

**Figure S17:** Relationship of disease diagnoses with childlessness by age 45 in women (Panel A) and 50 in men (Panel B) from population-based analyses with or without adjusting for individuals' highest education level, for 74 disease diagnoses that were significantly associated with childlessness from the main analysis.

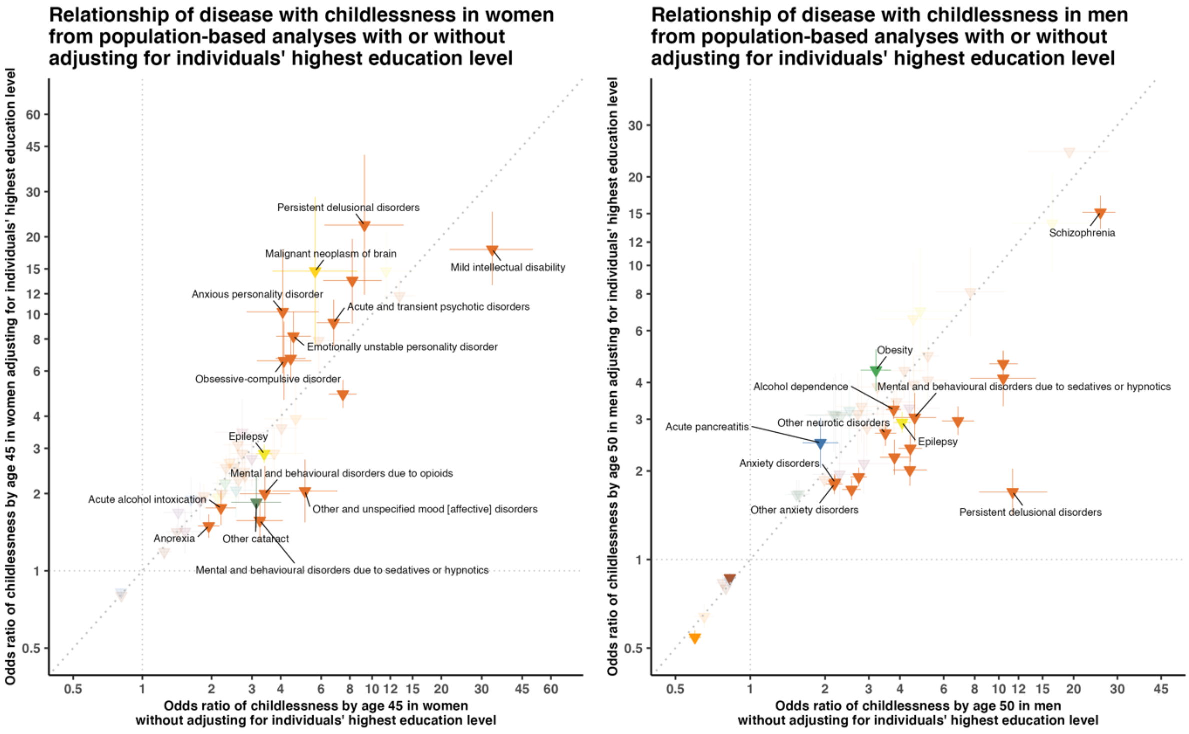

### Reference


1. Goldscheider, F.K. and Kaufman, G., 1996. Fertility and commitment: Bringing men back in. Population and Development Review, 22, pp.87-99.

2. Hellstrand, J., J. Nisén, and M. Myrskylä. 2020. All-time low period fertility in Finland: Demographic drivers, tempo effects, and cohort implications, Population Studies. doi:https://doi.org/10.1080/00324728.2020.1750677.

3. Andersson, L., 2021. Lifetime parenthood in the context of single-and multiple-partner fertility. Advances in Life Course Research, 47, p.100355.

4. Jalovaara, M., L. Andersson and A. Miettinen. (2022). Parity disparity: Educational differences in Nordic fertility across parities and number of reproductive partners, Population Studies, 76(1): 119-136. https://doi.org/10.1080/00324728.2021.1887506.

5. Rotkirch A, Miettinen A. Childlessness in Finland. In: Kreyenfeld M, Konietzka D, eds. *Childlessness in Europe: Contexts, causes, and consequences*. Cham: Springer, 2017: 139-158.

6. Jalovaara, M., G. Neyer, G. Andersson, J. Dahlberg, L. Dommermuth, P. Fallesen, and T. Lappegård. 2018. Education, gender, and cohort fertility in the Nordic countries, European Journal of Population 35(3): 563–586. doi:https://doi.org/10.1007/s10680-018-9492-2.

7. Holland, J. A., and E. Thomson. 2011. Stepfamily childbearing in Sweden: Quantum and tempo effects, 1950–99, Population Studies 65(1): 115–128. doi:https://doi.org/10.1080/00324728.2010.543693.

8. Wood, J., Neels, K. and Kil, T., 2014. The educational gradient of childlessness and cohort parity progression in 14 low fertility countries. Demographic research, 31, pp.1365-1416.

9. Schoen, R. 2006. Insights from parity status life tables for the 20th century U.S., Social Science Research 35(1): 29–39. doi:https://doi.org/10.1016/j.ssresearch.2004.06.002.
